# Individual Angiotensin Receptor Blockers and Risk of Dementia: A Target Trial Emulation Study

**DOI:** 10.64898/2026.10.13.26365306

**Authors:** Nicholas H Hampilos, Thomas R Campion, Evan Sholle, Chengxi Zang, Lidia Glodzik, Tracy A Butler, Lishomwa C Ndhlovu

**Author notes:** **Corresponding author**: Nicholas H Hampilos, M.D. Department of Radiology, Weill Cornell Medicine 413 E 69th St, 8^th^ Floor, New York, NY 10021, USA.

## Abstract

**INTRODUCTION:** Observational studies suggest that angiotensin receptor blockers (ARBs) are associated with lower dementia risk than angiotensin-converting enzyme inhibitors (ACEIs). Whether dementia risk differs across individual ARBs remains uncertain and was investigated in this study.

**METHODS:** This retrospective cohort study emulated target trials using electronic health record and claims data from 44 US health care organizations in the TriNetX Linked Research Network. ACEI/ARB-naïve adults aged 55–80 years initiating an ARB or lisinopril were eligible. Separate 1:1 propensity-score matching was performed for each comparison. Individual and pooled ARBs were compared with lisinopril (between-class), and non-losartan ARBs with losartan (within-class). The primary outcome was incident all-cause dementia; secondary outcomes included Alzheimer disease, vascular dementia, cerebral infarction, death, and dementia or death. Cox models estimated hazard ratios (HRs) and 95% CIs. Sensitivity analyses included lagged and treatment-persistence analyses, control outcomes, and E-values.

**RESULTS:** Of 22,025,599 individuals screened, 2,051,275 were eligible; matched cohorts ranged from 10,113–857,773, with median follow-up of 4.7–7.2 years. Pooled ARBs were associated with lower dementia risk than lisinopril (HR, 0.86; 95% CI, 0.85–0.87), with substantial variation across individual agents (HR range, 0.66–0.93). Within-class comparisons showed similar heterogeneity (HR range, 0.68–0.94) and reproduced between-class agent ordering by HR estimates. Considerable agent-level variation was also observed across secondary outcomes. Losartan, the most commonly prescribed ARB, had the least favorable dementia associations. Negative-control associations were near null within class, and sensitivity analyses supported robustness.

**DISCUSSION:** Individual ARBs were associated with differing dementia risks, suggesting clinically relevant within-class heterogeneity pending confirmation in randomized trials.

## 1. Background

Alzheimer disease and related dementias affect more than 50 million people worldwide, a number projected to exceed 130 million by 2050^1^. Disease-modifying therapies have recently emerged but remain costly, of limited availability, and modest in effect, and existing symptomatic treatments provide limited benefit^2^. Because novel drug development is slow and resource-intensive, there is growing interest in identifying and repurposing existing, approved interventions with known safety profiles for dementia prevention^3^. Large-scale clinical datasets with long-term follow-up may identify such candidates, as illustrated by a recent population study linking varicella-zoster vaccination to lower dementia risk^4^, which can then be evaluated in randomized trials^5^.

Among antihypertensive medications, angiotensin receptor blockers (ARBs) have been associated with lower dementia risk than other classes, including angiotensin-converting enzyme inhibitors (ACEIs)^6–10^. Although generally considered interchangeable for blood pressure control^11^, individual ARBs differ in half-life, receptor affinity and dissociation kinetics at the angiotensin II type 1 receptor, and central nervous system penetration, which may confer differential neuroprotective effects^12–14^. Whether these properties translate into clinically meaningful differences in dementia risk remains unclear, as prior studies have primarily evaluated class-level effects and lacked power to assess agent-specific outcomes^6–10^.

We conducted a multisite observational study to evaluate associations between individual ARBs and incident dementia among ACEI/ARB naïve adults aged 55 to 80 years. Using an active-comparator, propensity-score–matched design, we emulated a target trial comparing initiation of individual and pooled ARBs with lisinopril as the between-class reference and losartan as the within-class reference.

## 2 Methods

### 2.1 Study Design and Data Source

This retrospective cohort study emulated a target trial^15,16^ comparing individual and pooled ARBs with lisinopril (between-class) and losartan (within-class) for dementia risk (Table S1; Appendix S1). Data were obtained from the TriNetX Linked Research Network, a federated database providing access to de-identified electronic health records and insurance claims from 44 academic and community health care organizations in the United States^17^.

TriNetX data are standardized to common clinical terminologies and undergo routine platform quality control. Mortality was ascertained using the TriNetX “deceased” indicator, which integrates death information from health care organization records, claims data, and external mortality sources, with claims linkage enhancing capture of out-of-system deaths. Data were queried in June and July 2026.

This study used only de-identified data and did not constitute human subjects research; institutional review board review and informed consent were therefore not required. This study followed the Strengthening the Reporting of Observational Studies in Epidemiology (STROBE) reporting guidelines.

### 2.2 Target Trial Design

Eligible participants were ACEI/ARB-naïve adults 55 to 80 years initiating an ARB or lisinopril. Eligibility was based on treatment initiation rather than a recorded diagnosis of hypertension or heart failure because these medications are used for multiple indications and diagnosis capture may be incomplete in EHR data. Hypertension, heart failure, kidney disease, and other relevant conditions were included as baseline covariates. The index date (Day 0) was defined as the first recorded prescription, at which eligibility was assessed and treatment initiated. For each outcome, participants with a prior diagnosis of that outcome were excluded during cohort construction; additionally, TriNetX excluded patients with the outcome before the analysis window^18^. Participants were also required to have no prior exposure to an ARB or ACEI at any time before the index date (through Day −1), consistent with a new-user design (Table S1; Figure S1).

Treatment strategies comprised pooled and individual ARBs (candesartan, irbesartan, losartan, olmesartan, telmisartan, and valsartan) versus lisinopril in between-class comparisons and individual non-losartan ARBs versus losartan in within-class comparisons. Lisinopril and losartan were selected as fixed references because they are the most commonly prescribed ACEI and ARB, respectively, in the United States^19^.

For each pairwise comparison, a separate cohort and propensity-score model were constructed. Propensity scores were estimated using logistic regression including demographic characteristics, medical and psychiatric comorbidities, cardiovascular, antithrombotic and lipid-modifying medications, and proxy measures of health care utilization and socioeconomic status (Table S2). Baseline covariates were assessed from Day −365 to −1. One-to-one nearest-neighbor matching without replacement was performed. Covariate balance was assessed using standardized mean differences, with values below 0.1 indicating adequate balance. No imputation was performed; unknown demographic data were retained as separate categories.

### 2.3 Primary and Secondary Endpoints

The primary outcome was incident all-cause dementia, defined as the first recorded diagnosis of Alzheimer disease, vascular dementia, unspecified dementia, or dementia in other diseases (Table S3). Secondary outcomes included Alzheimer disease, vascular dementia, cerebral infarction, death, and a composite of all-cause dementia and death (Table S3).

### 2.4 Statistical Analyses

The primary estimand was the association of initial treatment with incident all-cause dementia under an intention-to-treat framework, expressed as the hazard ratio for each ARB or pooled ARBs versus lisinopril and each non-losartan ARB versus losartan. Participants were analyzed according to their index prescription regardless of subsequent treatment changes. The same framework was applied to secondary and control outcomes. Prespecified negative controls (carpal tunnel syndrome and hernia) assessed residual confounding (Table S3). Angioedema, which occurs more frequently with ACEIs than with ARBs^20^, was included as a positive-control outcome (Table S3). No a priori sample-size calculation was performed; cohort size was determined by prespecified eligibility criteria.

Cox proportional-hazards models were fitted within propensity-score–matched cohorts to estimate hazard ratios and 95% confidence intervals. Follow-up began on Day +1 to separate treatment initiation from outcome ascertainment and continued until the outcome, death (for nonmortality outcomes), or loss to follow-up, defined by the last recorded clinical encounter. Competing-risk analyses within the matched cohorts were not feasible in the aggregate TriNetX analytic workflow available for this study. Death was evaluated separately and in a composite outcome of dementia or death. The proportional hazards assumption was assessed using scaled Schoenfeld residuals, which, as previously described, were interpreted as qualitative indicators rather than by a strict significance threshold (Table S4-5)^18^. Ten-year absolute risk reductions (ARRs) and numbers needed to treat (NNTs) were calculated from Kaplan–Meier cumulative incidence. E-values were calculated for point estimates and the confidence-interval limit closest to the null to quantify the minimum strength of unmeasured confounding required to explain the observed associations^21^.

Sensitivity analyses in the irbesartan- and losartan-versus-lisinopril comparisons assessed treatment persistence and switching, dose effects, reverse causation, and differential follow-up. Per-protocol–like analyses required at least 2 recorded prescriptions within 12 months and no subsequent switching to another ACEI or ARB, providing a proxy for treatment persistence and switching^18^. 1- and 3-year lags were examined to evaluate reverse causation and protopathic bias. Analyses restricted to 10 years of follow-up evaluated the influence of differences in follow-up duration. Dose-stratified analyses compared low- and high-dose ARBs with corresponding lisinopril strata.

Analyses were primarily conducted within the TriNetX platform, with additional calculations performed as described in the Supplementary Appendix. Analyses were not adjusted for multiple comparisons, and secondary outcomes are hypothesis-generating. Results should be interpreted as estimates of association given the observational nature of the data.

## 3 Results

### 3.1 Cohort Characteristics and Follow-up

Among 22,025,599 individuals in the TriNetX Linked Research Network, 2,051,275 adults aged 55 to 80 years initiated an ARB or lisinopril and were potentially eligible. Of these, 956,771 initiated an ARB and 1,102,710 initiated lisinopril. Losartan was the most commonly prescribed ARB (583,461 patients) and candesartan the least common (10,113 patients) (Table S5).

Propensity-score matching yielded well-balanced cohorts, with all standardized mean differences below 0.1 (Table S6A-G; Table S7A-E). After matching, cohort sizes ranged from 10,113 to 857,773 between classes and from 10,113 to 202,904 for within-class comparisons (Table S8-9). Across matched cohorts, mean age ranged from 63.4 to 64.3 years, 51.1% to 54.0% were female (Table S6A-G; Table S7A-E). Median follow-up ranged from 4.7 to 7.2 years for between-class comparisons and 4.9 to 7.2 years for within-class comparisons (Table S10-11).

### 3.2 Individual ARBs and Dementia Risk

As a class, ARBs were associated with lower incident all-cause dementia than lisinopril (hazard ratio [HR], 0.86; 95% confidence interval [CI], 0.85-0.87) (**Figure 1**). Substantial heterogeneity was observed across individual agents, with HRs ranging from 0.66 (95% CI, 0.58-0.74) for candesartan to 0.93 (95% CI, 0.91-0.94) for losartan (**Figure 1**). Ten-year ARRs ranged from 0.72% to 3.07% (NNT of 33 to 138; Table S12). Kaplan-Meier curves illustrated this heterogeneity (Figure S2A-G), with representative comparisons shown in **Figure 2**. Irbesartan, among the most protective ARBs (HR 0.69, 95% CI, 0.66-0.73), demonstrated greater separation from lisinopril, with progressive widening over time (**Figure 2A**). In contrast, losartan displayed more modest but consistent separation (**Figure 2B**).

**Figure 1.**
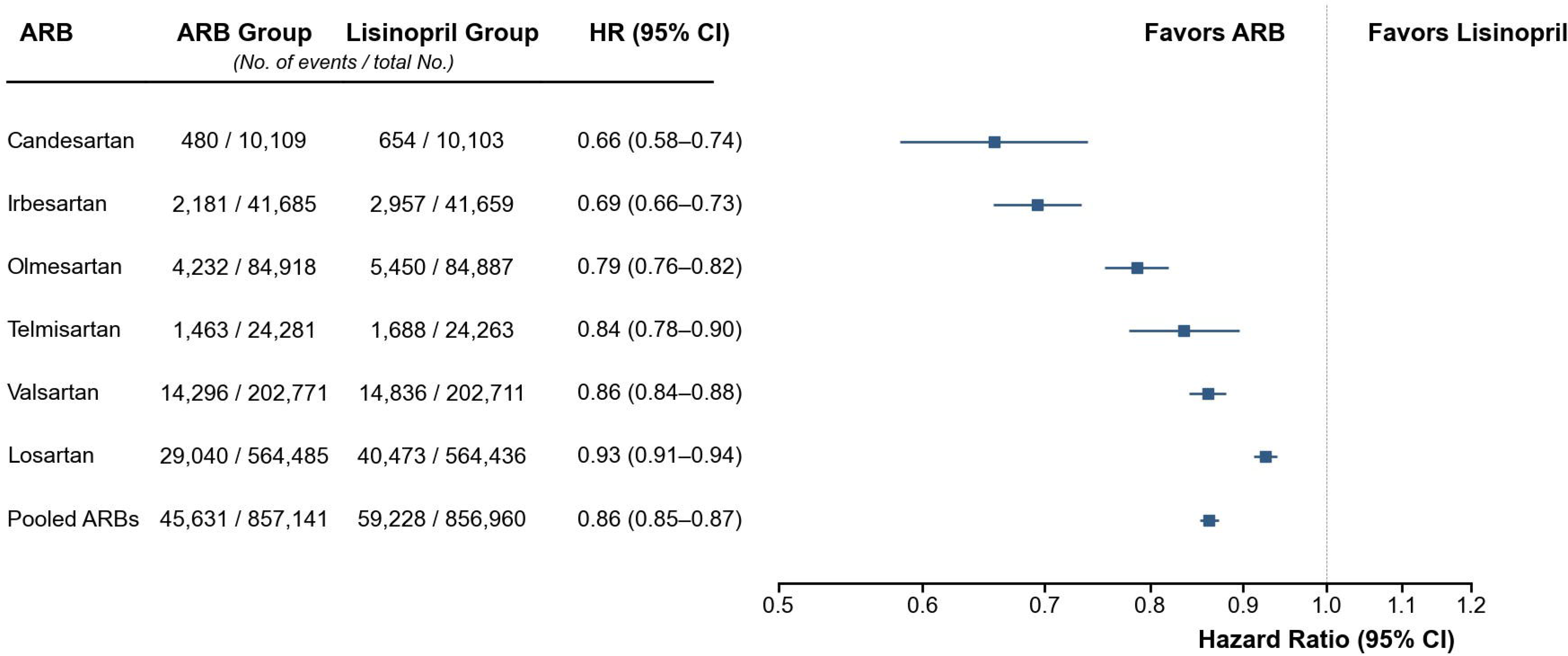
Primary Outcome in Between-Class Comparisons of ARBs vs Lisinopril. Forest plots show hazard ratios and 95% confidence intervals for the primary outcome (incident all-cause dementia) comparing individual and pooled angiotensin receptor blockers (ARBs) with lisinopril (reference). Hazard ratios are plotted on a logarithmic scale. Incident all-cause dementia codes included Alzheimer disease, vascular dementia, unspecified dementia, and dementia in other diseases, using International Classification of Diseases, 10th Revision (ICD-10) codes. Python/Matplotlib code used to render this figure was generated with assistance from ChatGPT (OpenAI), Claude (Anthropic), and/or Gemini (Google); the figure was verified against the source data.

**Figure 2.**
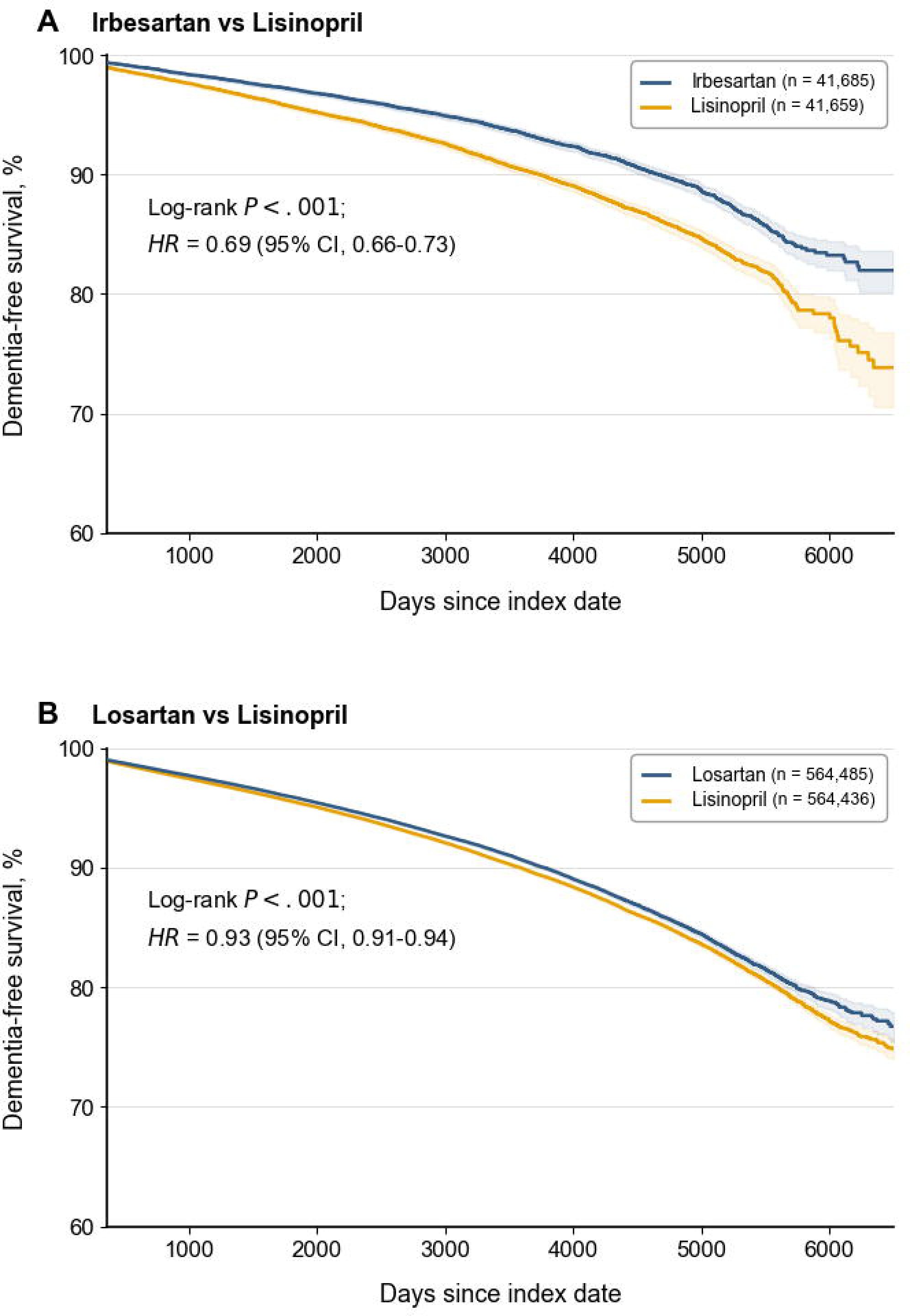
Dementia-free Survival in Select Between-Class Comparisons. Kaplan–Meier estimates of dementia-free survival for (A) irbesartan vs lisinopril (2,181/41,685 vs 2,957/41,659 events) and (B) losartan vs lisinopril (29,040/564,485 vs 40,473/564,436 events). Median observed follow-up (record length) was 2,415 days (6.61 years) for irbesartan and 2,429 days (6.65 years) for lisinopril in panel A, and 1,727 days (4.73 years) for losartan and 2,343 days (6.41 years) for lisinopril in panel B; this reflects the duration of available observation and is not censored at the outcome. Patients were followed from 1 day after the index date until the outcome (incident all-cause dementia), death, or loss to follow-up, defined by the last recorded clinical encounter. Hazard ratios (HR) with 95% confidence intervals and log-rank *P* values were computed over the entire follow-up period. For visual clarity, curves in this figure are truncated during the sparse terminal follow-up period; this visual truncation did not affect the hazard-ratio estimates. The full set of between-class Kaplan–Meier curves is provided in eFigure 2. Shaded areas indicate 95% confidence intervals, which widen at later time points as observations become sparse. Numbers at risk at individual follow-up time points are not shown because outcome-specific risk-set counts were not available from the aggregate TriNetX analytic output used for this study. Abbreviations: HR, hazard ratio.

### 3.3 Dementia Subtypes, Cerebral Infarction, and Mortality

Associations varied across dementia subtypes and mortality outcomes (**Figure 3**). Losartan, the most frequently prescribed ARB, had the smallest reductions across all secondary outcomes (**Figure 1**; **Figure 3**). Risk reductions were larger for vascular dementia (HR range, 0.52-0.84) than for Alzheimer disease (HR range, 0.79-0.98) and cerebral infarction (HR range, 0.80-0.97), with non-overlapping confidence intervals in 5 of 6 agents for each comparison (**Figure 3**). In exploratory analyses, agent-level estimates for vascular dementia and cerebral infarction were positively correlated (Spearman ρ=0.83; P=.04; eFigure 12).

**Figure 3.**
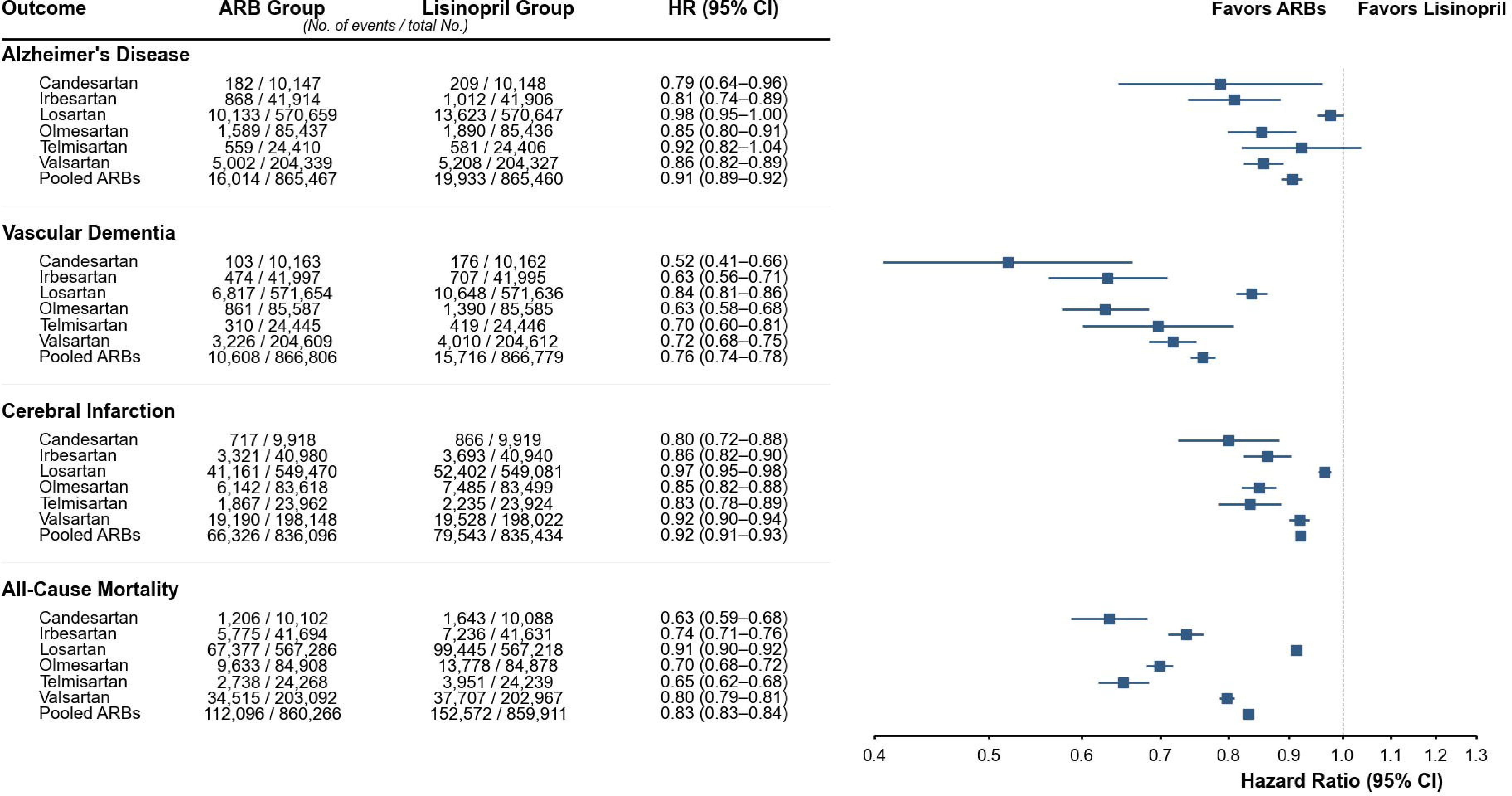
Secondary Outcomes in Between-Class Comparisons of ARBs vs Lisinopril. Forest plots show hazard ratios (HRs) and 95% confidence intervals for selected secondary outcomes comparing individual and pooled angiotensin receptor blockers (ARBs) with lisinopril (reference). Hazard ratios are plotted on a logarithmic scale. Secondary outcomes included Alzheimer disease, vascular dementia, cerebral infarction, all-cause mortality, and a composite outcome of dementia or death. Results for composite outcomes are presented in the Supplement. Outcomes were defined using International Classification of Diseases. Because analyses of secondary outcomes were not adjusted for multiple comparisons, these findings should be considered exploratory. Python/Matplotlib code used to render this figure was generated with assistance from ChatGPT (OpenAI), Claude (Anthropic), and/or Gemini (Google); the figure was verified against the source data.

All-cause mortality HRs ranged from 0.63 (95% CI, 0.59-0.68) for candesartan to 0.91 (95% CI, 0.90-0.92) for losartan (**Figure 3**). Ten-year ARRs ranged from 1.84% to 7.07% (NNT, 14 to 54; eTable 8). Agent-level mortality and dementia estimates were positively but non-significantly correlated (Spearman ρ=0.77; P=.07), with candesartan and losartan anchoring both extremes (Figure S3). Mortality curves showed minimal early separation and greater divergence over time (Figure S4A-G). Results for the composite outcome of dementia or death generally fell between the corresponding dementia and mortality estimates (Figure S5).

Within-class comparisons using losartan as the reference similarly demonstrated heterogeneity, with HRs for all-cause dementia ranging from 0.68 (95% CI, 0.60–0.77) for candesartan to 0.94 (95% CI, 0.91–0.96) for valsartan (**Figure 4**). All 5 non-losartan agents were associated with lower dementia risk than losartan, and the relative ordering of agent-level point estimates was identical to that observed in the between-class analyses, despite separate cohort construction and matching (**Figure 4**). As in between-class analyses, associations were generally larger for vascular dementia and mortality than Alzheimer disease and cerebral infarction, with losartan the least favorable agent throughout; within-class effects were attenuated relative to between-class estimates, and several Alzheimer disease comparisons were not statistically significant (Figure S6). Additional Kaplan–Meier curves, secondary analyses and ARR/NNT calculations for within-class comparisons are provided in the Supplemental Appendix (Figure S7-S9, Table S13).

**Figure 4.**
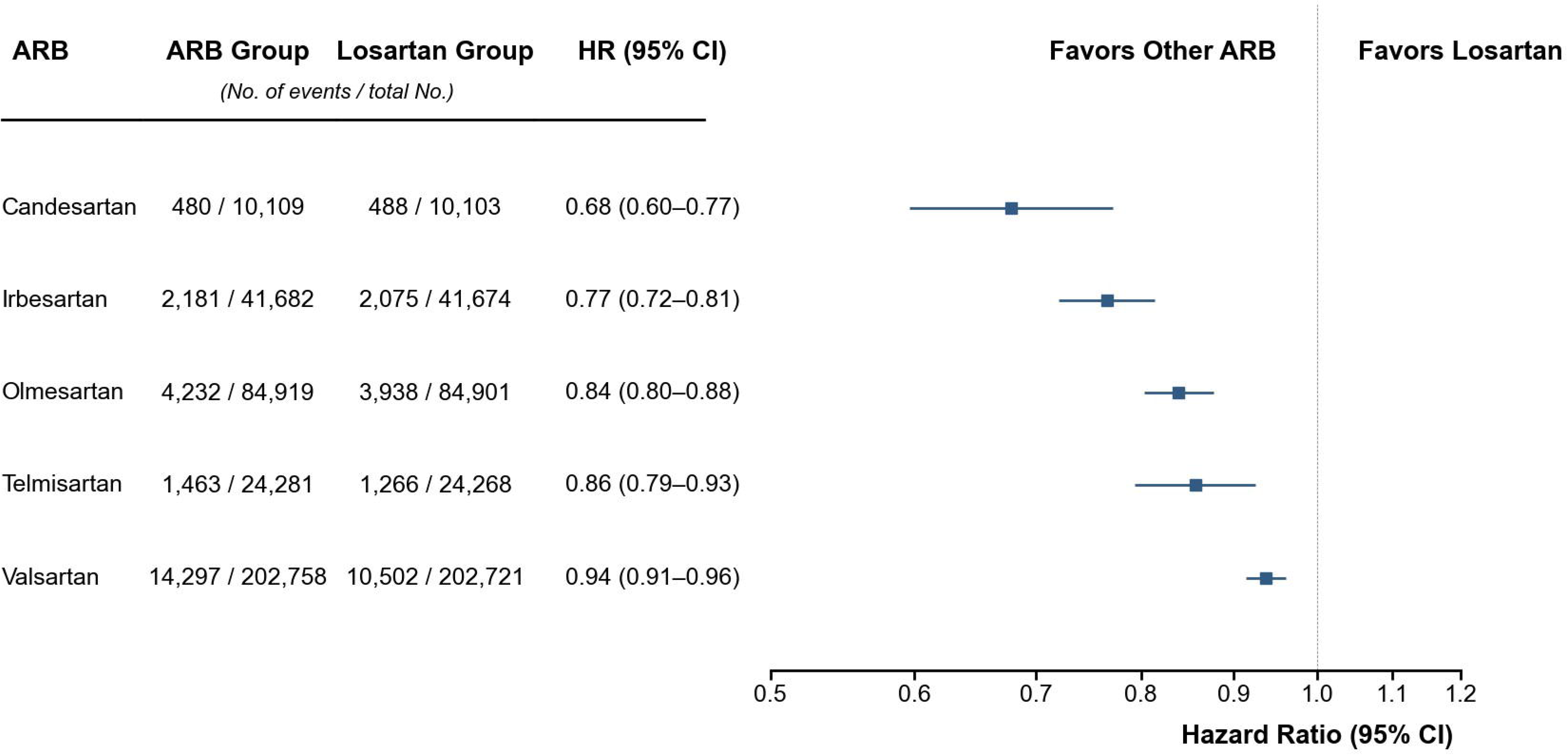
Primary Outcome in Within-Class Comparisons of other ARBs vs Losartan. Forest plots show hazard ratios and 95% confidence intervals for the primary outcome (incident all-cause dementia) comparing other angiotensin receptor blockers (ARBs) with losartan (reference). Hazard ratios are plotted on a logarithmic scale. Incident all-cause dementia codes included Alzheimer disease, vascular dementia, unspecified dementia, and dementia in other diseases, using International Classification of Diseases, 10th Revision (ICD-10) codes. Python/Matplotlib code used to render this figure was generated with assistance from ChatGPT (OpenAI), Claude (Anthropic), and/or Gemini (Google); the figure was verified against the source data.

### 3.4 Sensitivity and Bias Analyses

To evaluate the robustness of the primary analyses, we performed sensitivity analyses in two representative comparisons spanning the range of observed effects, irbesartan and losartan versus lisinopril. In per-protocol-like analyses, associations were stronger than intention-to-treat estimates for irbesartan (HR, 0.59 vs 0.69) and losartan (HR, 0.86 vs 0.93) (Table S14). Estimates remained protective across 1-day (main analysis), 1- and 3-year lags (irbesartan, 0.69 [0.66–0.73] to 0.73 [0.68–0.78]; losartan, 0.93 [0.91–0.94] to 0.95 [0.93–0.96]) (Table S15). Restricting follow-up to 10 years produced estimates similar to the primary analyses for both irbesartan (HR, 0.69; 95% CI, 0.65–0.73 vs 0.69; 95% CI, 0.66–0.73) and losartan (HR, 0.92; 95% CI, 0.90–0.93 vs 0.93; 95% CI, 0.91–0.94) (Table S16). In dose-stratified analyses, irbesartan was more strongly associated with lower dementia risk than losartan, with both compared with lisinopril (Table S17).

Negative control outcomes (carpal tunnel syndrome and hernia) were modestly elevated in between-class comparisons (HR range, 1.00–1.20, with 11/14 confidence intervals excluding 1; Figure S10), but attenuated to near-null within class (HR range, 0.91–1.03, with all confidence intervals crossing or reaching 1; Figure S11). E-value point estimates ranged from 1.37 to 2.42 across between-class comparisons and 1.33 to 2.31 across within-class comparisons, with corresponding confidence-interval-limit E-values of 1.32 to 2.07 and 1.24 to 1.92 (Table S18). ARBs were associated with a substantially lower risk of angioedema than lisinopril (HR range, 0.48-0.60; Figure S12), recovering a known drug-class effect.

## 4 Discussion

In this target-trial emulation study drawing from more than 2 million potentially eligible patients, dementia associations varied substantially across individual ARBs. Within-class estimates using losartan as the reference also varied across ARBs, broadly consistent with the between-class pattern. Associations were larger for vascular dementia than for Alzheimer disease or cerebral infarction, whereas mortality estimates did not uniformly parallel dementia. Losartan had the least favorable estimates across all outcomes. The large cohort size and long follow-up enabled informative agent-specific estimates, including for losartan and valsartan. Treating ARBs as interchangeable may therefore obscure clinically relevant differences.

Consistent with prior observational studies^6–10^, ARBs as a class were associated with modestly lower dementia risk than lisinopril (HR, 0.86), although pooled estimates varied substantially across studies. Prior agent-specific analyses were exploratory and imprecise; additionally, the aforementioned prior study cohort lacked losartan and included relatively few valsartan users^7^, which represent the 2 most commonly prescribed ARBs in the United States and globally. Our large sample and long follow-up enabled estimation across individual agents, revealing substantial heterogeneity in both between- and within-class analyses (HR range, 0.66– 0.93; 10-year ARRs, 0.7%-3.1%). Differences in prescribing composition may therefore contribute to variation in prior pooled class-level estimates. For example, the previously mentioned study reporting a stronger ARB-versus-ACE-inhibitor association (HR, 0.72) lacked losartan and included few valsartan events^7^. Underrepresentation of these less protective agents could yield a relatively more protective pooled estimate, although differences in ACEI comparators may also contribute.

The observed heterogeneity in dementia outcomes across ARBs is unlikely to be fully explained by blood pressure lowering. ARBs and lisinopril lower blood pressure comparably as classes, and inter-agent differences among ARBs are generally modest^11,13,22^. The larger association with vascular dementia is compatible with a microvascular contribution: experimental evidence indicates that ARBs may attenuate oxidative stress, improve endothelial function, and enhance nitric oxide bioavailability^23,24^, effects that could act on small-vessel pathology independently of blood pressure. Other pharmacologic differences, including AT1-receptor binding kinetics, central nervous system penetration, and non-AT1-receptor effects, may also contribute. For example, candesartan, the most protective agent in our analysis, has a longer half-life and produces more sustained, insurmountable AT1-receptor blockade than losartan, the least protective agent^12,13^. If sustained receptor blockade is important for central nervous system protection, these properties could plausibly contribute to their differing dementia associations, although this hypothesis requires direct investigation.

The timing of ARB initiation may also influence neuroprotective benefit, and prior randomized trials provide context. In older adults with mild cognitive impairment, candesartan improved executive function and memory versus lisinopril despite similar blood pressure control, consistent with a blood pressure–independent effect^22^. In contrast, the RADAR trial of losartan, the ARB with the least favorable estimates in our analysis, showed no benefit on brain atrophy in established Alzheimer disease, possibly reflecting late initiation^25^. Although these trials differed, their findings are compatible with agent-specific heterogeneity and a potential role for treatment timing, particularly before neurodegeneration is established.

ARBs were also associated with lower mortality than lisinopril, contrasting with ONTARGET^26^ and VALIANT^27^, which used ramipril and captopril as between-class comparators and found no significant mortality difference between ARB and ACEI regimens. The use of different ACEI comparators may partly explain this discordance. Prior observational studies comparing mortality risk between ARBs are also mixed. A recent target-trial emulation using the TriNetX global cohort found lower mortality with telmisartan than with pooled ARBs, losartan, and valsartan^28^. Conversely, a Korean national cohort found no adjusted mortality differences across seven ARBs despite substantial crude differences^29^. Differences in populations, comparators, exposure definitions, covariate timing, and analytic methods may contribute to this discordance. Residual confounding, including healthy-adherer and prescribing-channeling effects, remains plausible; near-null negative controls in our within-class comparisons argue against gross bias but cannot exclude it. The discrepancy in mortality associations therefore underscores the need for independent replication and prospective confirmation.

The potential clinical implications of antihypertensive selection as a modifiable determinant of long-term dementia risk are substantial. ARBs and ACEIs are used by millions worldwide^30,31^, so even modest individual risk reductions could yield meaningful population-level impact. If causal, preferential use of ARBs associated with lower dementia risk than losartan or lisinopril could prevent tens of thousands of dementia cases per million treated over 10 years (Appendix S2). Unlike anti-amyloid therapies, which act on established or prodromal disease at substantial cost, antihypertensive selection could represent a low-cost, population-scale strategy implemented before disease onset, complementing rather than replacing anti-amyloid treatment.

We performed complementary sensitivity and bias analyses supporting the primary findings. Negative-control associations attenuated from modest elevations between classes to near-null within class (HR range, 0.91-1.03), suggesting reduced systematic bias. Within-class analyses similarly demonstrated heterogeneity and a broadly consistent ordering of agent-level point estimates despite separate cohort construction and matching. Because ARB-vs-ARB comparisons are less susceptible to class-level prescribing channeling, this consistency argues against comparator-related confounding as the sole explanation. Since non-losartan ARBs were not directly compared and confidence intervals overlapped, this ordering should be interpreted descriptively. E-values ranged from 1.37 to 2.42 for point estimates and 1.32 to 2.07 for confidence-interval limits. Together, these analyses make confounding by indication alone less likely to explain the observed heterogeneity, although residual confounding cannot be excluded.

### Limitations

This study has several limitations. As an observational study, causal effects cannot be established, and residual confounding remains possible. Electronic health record data are subject to misclassification and incomplete capture; blood pressure and medication adherence were not directly observed, although randomized data indicate comparable blood pressure lowering across agents. Dementia was identified from diagnostic codes without adjudication, and differential cognitive workup could contribute to misclassification. However, losartan also had the least favorable mortality estimates, making differential cognitive workup unlikely to fully explain the pattern. Aggregate platform constraints precluded time-varying analyses and competing-risk analyses within matched cohorts. Generalizability may be limited in populations and health systems with different prescribing patterns.

## Conclusions

Individual ARBs were associated with differing dementia risk, suggesting clinically relevant heterogeneity within this drug class. Given their widespread use, safety profile, and low cost, certain ARBs may be pragmatic candidates for dementia prevention, particularly where access to newer disease-modifying therapies is limited. Ultimately, randomized trials are needed to determine whether these differences reflect causal effects and to inform optimal antihypertensive selection.

## AUTHOR CONTRIBUTIONS

N.H. Hampilos: study concept and design; acquisition, analysis, and interpretation of data; drafting and critical revision of the manuscript. T.R. Campion: analysis and interpretation of data; critical revision of the manuscript. E. Sholle: analysis and interpretation of data; critical revision of the manuscript. C. Zang: analysis and interpretation of data; critical revision of the manuscript. L. Glodzik: analysis and interpretation of data; critical revision of the manuscript. T.A. Butler: analysis and interpretation of data; critical revision of the manuscript. L.C. Ndhlovu: study concept and design; analysis and interpretation of data; critical revision of the manuscript. All authors approved the final version of the manuscript and agree to be accountable for the work.

## ACKNOWLEDGEMENTS

The authors thank the TriNetX Research Network and participating health care organizations for providing access to the data. During manuscript preparation, the corresponding author used ChatGPT (OpenAI; GPT-5-series models, 2026), Claude (Anthropic; Claude Opus 4.8 and earlier models, 2026), and Gemini (Google, 2026) for language editing and revision and to assist with generation of Python/Matplotlib code for study figure preparation. AI-generated code was used solely to render figures from study data and analytic outputs; AI tools were not used to perform study analyses, generate study results, or formulate scientific conclusions. All AI-assisted text, code, and resulting figures were reviewed and verified by the corresponding author, who takes responsibility for their use. All authors approved and take responsibility for the final manuscript.

## CONSENT STATEMENT

This study used only de-identified data and did not constitute human subjects research; institutional review board review and informed consent were therefore not required.

## DATA AVAILABILITY STATEMENT

Individual-level data from the TriNetX Linked Research Network are not publicly available and cannot be shared by the authors. Access is available to authorized users through institutional agreements with TriNetX. Aggregate results underlying the analyses are available from the corresponding author upon reasonable request.

