## Supplemental Appendix for "Individual Angiotensin Receptor Blockers and Risk of Dementia: A Target Trial Emulation Study"

**Table S1. Target Trial Specification and Corresponding Emulation Using Real-World Data**

| **Component** | **Target Trial Specification** | **Target Trial Emulation** |
| --- | --- | --- |
| **Eligibility Criteria** | Adults aged 55–80 eligible to an ARB or lisinopril, with no prior ACEI/ARB use and no prior outcome at baseline. | Adults aged 55–80 with a first recorded ARB or lisinopril prescription; excluded if any prior ACEI/ARB use before Day −1, or a prior diagnosis of the specific outcome under analysis (outcome-specific: each cohort excluded only prior diagnoses of the outcome being analyzed). |
| **Treatment Strategies** | Group 1: Initiate and continue an ARB. Group 2: Initiate and continue the reference agent (lisinopril for between-class comparisons; losartan for within-class comparisons). | Group 1: Initiation of an individual ARB (candesartan, irbesartan, losartan, olmesartan, telmisartan, valsartan) or pooled ARBs. Group 2: Initiation of the reference agent at the index date — lisinopril for between-class comparisons and losartan for within-class (ARB vs ARB) comparisons. |
| **Assignment** | Randomized assignment at baseline, with or without blinding. | Non-random assignment; emulated using 1:1 propensity-score matching on demographics, comorbidities, medications, and healthcare utilization. Each comparison (each ARB and pooled ARBs vs lisinopril; each ARB vs losartan) was matched within a separately constructed cohort. |
| **Time Zero (Baseline)** | Time of treatment assignment; eligibility and assignment aligned at baseline, with follow-up beginning immediately after assignment. | Index date = first prescription (Day 0); treatment assignment occurs at Day 0. Eligibility is assessed at baseline using data available prior to treatment initiation (≤ Day −1), including absence of prior ACEI or ARB use and prior outcome. Follow-up begins on the day after treatment initiation (Day +1). |
| **Exclusion Window** | Exclude prior ACEI or ARB exposure, and prior diagnosis of the outcome under analysis, before baseline. | Prior ACEI/ARB exposure and prior diagnosis of the analyzed outcome were assessed using all available data up to Day −1. |
| **Baseline Covariates** | Measured prior to treatment assignment. | Assessed during Day −365 to −1 (demographics, comorbidities, medications, healthcare utilization). |
| **Follow-up** | From treatment initiation to outcome, death, loss to follow-up, or administrative censoring. | From the day after treatment initiation (Day +1) to outcome, death, loss to follow-up, or administrative censoring at the end of the study period. |
| **Outcomes** | Time to incident dementia (primary) and related outcomes. | Time to first diagnosis of all-cause dementia (primary); secondary outcomes were Alzheimer disease, vascular dementia, cerebral infarction, death, and a composite of all-cause dementia and death. |
| **Causal Contrast** | Intention-to-treat and per-protocol effects comparing ARBs vs lisinopril, quantified using hazard ratios. | Primary: intention-to-treat, analyzing individuals according to their index treatment assignment in each pairwise comparison, regardless of subsequent treatment changes. Sensitivity analysis: per-protocol–like analyses in selected between-class comparisons required at least 2 recorded prescriptions of the index agent within 12 months and no subsequent exposure to another ARB or ACEI. |
| **Statistical Analysis** | Time-to-event analysis comparing treatment strategies, with adjustment for baseline confounders. | Cohorts matched 1:1 by propensity score (nearest-neighbor); balance assessed by standardized mean differences (threshold 0.1). Cox proportional-hazards models estimated hazard ratios and 95% confidence intervals. For non-mortality outcomes, death was treated as a censoring event. The proportional-hazards assumption was evaluated, and the log-rank test (which does not assume proportionality) was reported alongside. Kaplan–Meier curves were generated. Tests were 2-sided (α = 0.05). |

**Figure S1. Target trial emulation design and follow-up schematic**

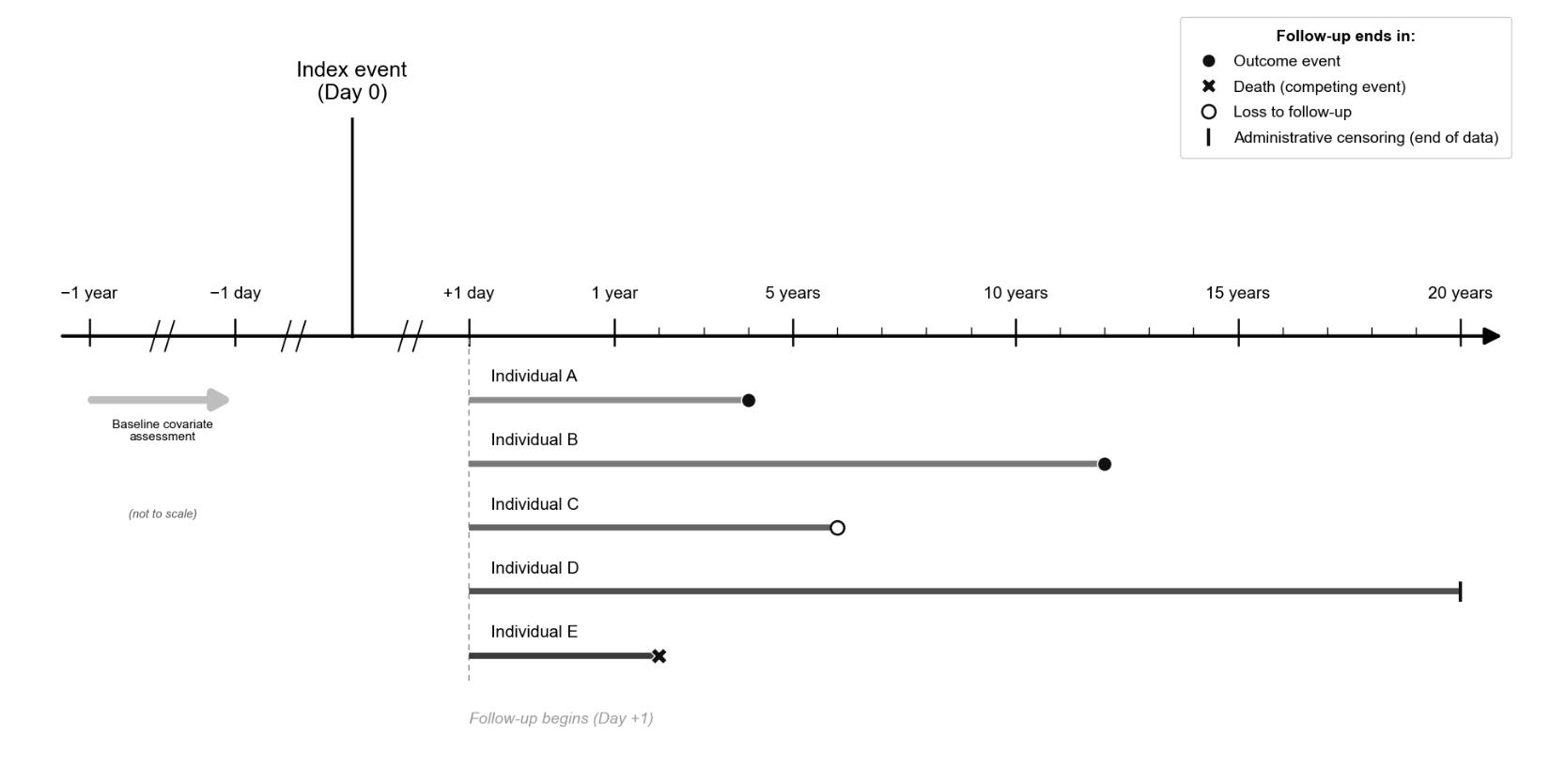

Schematic representation of the new-user, active-comparator design comparing initiation of individual and pooled ARBs with lisinopril or losartan. Baseline covariates were assessed during the 365 days before the index date (first qualifying prescription; Day 0). Follow-up began the day after treatment initiation (Day +1) and continued until the first of an incident outcome, death, loss to follow-up (last recorded encounter), or administrative censoring at the end of data availability. Five hypothetical individuals (A–E) illustrate variable follow-up durations and the four ways follow-up could end, as indicated in the legend. The baseline window and the ±1-day interval around the index date are not drawn to scale, as indicated by the axis breaks (//). Python/Matplotlib code used to render this figure was generated using ChatGPT (OpenAI; GPT-5-series, 2026), Claude (Anthropic; Claude Opus 4.8 and earlier models, 2026), and/or Gemini (Google; 2026); the figure was verified against the source data.

**Table S2.** **Covariate definitions and codes**

| **Category** | **Covariate** | **Code System** | **Codes** |
| --- | --- | --- | --- |
| **Demographics** | Age at index | — | — |
|  | Sex (female, male) | — | — |
|  | Race (White, Black or African American, Asian, American Indian or Alaska Native, Native Hawaiian or Other Pacific Islander, Other Race, Unknown Race) | — | — |
|  | Ethnicity (Hispanic or Latino, Not Hispanic or Latino, Unknown Ethnicity) | — | — |
| **Clinical Diagnoses** | Essential (primary) hypertension | ICD-10-CM | I10 |
|  | Acute myocardial infarction | ICD-10-CM | I21 |
|  | Chronic ischemic heart disease | ICD-10-CM | I25 |
|  | Atrial fibrillation and flutter | ICD-10-CM | I48 |
|  | Heart failure | ICD-10-CM | I50 |
|  | Cerebral infarction | ICD-10-CM | I63 |
|  | Other peripheral vascular diseases | ICD-10-CM | I73 |
|  | Overweight | ICD-10-CM | E66.3 |
|  | Obesity, unspecified | ICD-10-CM | E66.9 |
|  | Type 2 diabetes mellitus | ICD-10-CM | E11 |
|  | Disorders of lipoprotein metabolism | ICD-10-CM | E78 |
|  | Chronic kidney disease | ICD-10-CM | N18 |
|  | Major depressive disorder | ICD-10-CM | F33 |
|  | Other anxiety disorders | ICD-10-CM | F41 |
|  | Mental and behavioral disorders due to psychoactive substance use | ICD-10-CM | F10-F19 |
|  | Nicotine dependence | ICD-10-CM | F17 |
|  | Alcohol related disorders | ICD-10-CM | F10 |
| **Healthcare Utilization / Socioeconomic Variables** | Medicaid status proxy (State Medicaid Agency Codes) | Procedure | T |
|  | Office visit (established patient) | Procedure | 1013638 |
|  | Preventative visit (new patient) | Procedure | 1013627 |
|  | Emergency department services | Procedure | 1013711 |
|  | Problems related to housing and economic circumstances | ICD-10-CM | Z59 |
| **Medications** | HMG-CoA reductase inhibitors (statins) | ATC | C10AA |
|  | Platelet aggregation inhibitors (excluding heparin) | ATC | B01AC |
|  | Direct factor Xa inhibitors | ATC | B01AF |
|  | Vitamin K antagonists | ATC | B01AA |
|  | Calcium channel blockers | ATC | C08 |
|  | Beta Blocking Agents | ATC | C07 |

Covariates were defined using diagnosis, procedure, and medication codes recorded in the electronic health record prior to the index date. Demographic variables were obtained from structured electronic health record fields. Healthcare utilization variables were included as proxies for healthcare engagement and socioeconomic status (Medicaid, problems related to housing and economic circumstances). Medication classes were identified using Anatomical Therapeutic Chemical (ATC) classification codes and reflect prescriptions recorded prior to cohort entry. All covariates were assessed during the baseline period and used for propensity-score matching. Abbreviations: ICD-10-CM, International Classification of Diseases, Tenth Revision, Clinical Modification; ATC, Anatomical Therapeutic Chemical.

**Table S3. Outcome definitions and codes**

| **Outcome** | **Classification** | **Code System** | **Codes** |
| --- | --- | --- | --- |
| **All-cause dementia**  (Alzheimer's disease, vascular dementia, unspecified dementia, or dementia in other diseases) | Primary outcome | ICD-10-CM | G30, F01, F03, F02 |
| Alzheimer's disease | Secondary outcome | ICD-10-CM | G30 |
| Vascular dementia | Secondary outcome | ICD-10-CM | F01 |
| Unspecified dementia | N/A | ICD-10-CM | F03 |
| Dementia in other diseases | N/A | ICD-10-CM | F02 |
| All-cause mortality | Secondary outcome | Demographics | Deceased |
| Composite outcome  (All-cause dementia or all-cause mortality) | Secondary outcome | ICD-10-CM and Demographics | G30, F01, F03, F02 and Deceased |
| Cerebral infarction | Secondary outcome | ICD-10-CM | I63 |
| Carpal tunnel syndrome | Negative control outcome | ICD-10-CM | G56.0 |
| Hernia | Negative control outcome | ICD-10-CM | K40-K46 |
| Angioneurotic edema (Angioedema) | Positive control outcome | ICD-10-CM | T78.3 |

Dementia outcomes were defined using ICD-10-CM diagnosis codes. The composite outcome was defined as the first occurrence of any dementia subtype or all-cause mortality, whichever occurred first. Mortality status was obtained from structured demographic records. Negative control outcomes were selected a priori based on the absence of known associations with the renin–angiotensin–aldosterone system; angioedema was included as a positive control outcome for ARB-vs-lisinopril comparisons. Code definitions were applied uniformly across all analyses. Abbreviations: ARB, angiotensin receptor blocker; ICD-10-CM, International Classification of Diseases, Tenth Revision, Clinical Modification.

**Table S4. Proportional hazards assumption p values of incident all-cause dementia and mortality for between-class comparisons**

| **Outcome** | **Comparison** | **Proportionality p value** |
| --- | --- | --- |
| **All-cause dementia** | Candesartan vs. Lisinopril | 0.15 |
|  | Irbesartan vs. Lisinopril | 0.04 |
|  | Losartan vs. Lisinopril | 0.003 |
|  | Olmesartan vs. Lisinopril | < 0.001 |
|  | Telmisartan vs. Lisinopril | < 0.001 |
|  | Valsartan vs. Lisinopril | < 0.001 |
|  | Pooled ARB vs. Lisinopril | < 0.001 |
| **Death** | Candesartan vs. Lisinopril | 0.44 |
|  | Irbesartan vs. Lisinopril | 0.002 |
|  | Losartan vs. Lisinopril | < 0.001 |
|  | Olmesartan vs. Lisinopril | < 0.001 |
|  | Telmisartan vs. Lisinopril | 0.12 |
|  | Valsartan vs. Lisinopril | 0.60 |
|  | Pooled ARB vs. Lisinopril | 0.02 |

Proportional hazards assumptions were assessed for each comparison using tests of nonproportionality provided within the TriNetX platform. P values correspond to tests of nonproportionality, with smaller values indicating greater evidence against the proportional hazards assumption.

**Table S5. Proportional hazards assumption p values for incident all-cause dementia and mortality for within-class comparisons**

| **Outcome** | **Comparison** | **Proportionality p value** |
| --- | --- | --- |
| **All-cause dementia** | Candesartan vs. Losartan | 0.21 |
|  | Irbesartan vs. Losartan | 0.26 |
|  | Olmesartan vs. Losartan | <.001 |
|  | Telmisartan vs. Losartan | <.001 |
|  | Valsartan vs. Losartan | 0.001 |
| **All-cause mortality** | Candesartan vs. Losartan | 0.007 |
|  | Irbesartan vs. Losartan | 0.001 |
|  | Olmesartan vs. Losartan | <.001 |
|  | Telmisartan vs. Losartan | 0.08 |
|  | Valsartan vs. Losartan | 0.006 |

Proportional hazards assumptions were assessed for each comparison using tests of nonproportionality provided within the TriNetX platform. P values correspond to tests of nonproportionality, with smaller values indicating greater evidence against the proportional hazards assumption.

**Table S6A. Candesartan vs. Lisinopril — baseline characteristics**

| **Characteristic** | **Before PSM** | | | **After PSM** | | |
| --- | --- | --- | --- | --- | --- | --- |
|  | **Candesartan (N=10,113)** | **Lisinopril (N=1,083,068)** | **SMD** | **Candesartan (N=10,113)** | **Lisinopril (N=10,113)** | **SMD** |
| Age — Mean (SD) | 63.6 (6.1) | 63.1 (6.0) | 0.097 | 63.6 (6.1) | 63.6 (6.1) | 0.002 |
| **Sex — no. (%)** |  |  |  |  |  |  |
| Female | 5,464 (54.0%) | 504,922 (46.6%) | 0.149 | 5,464 (54.0%) | 5,358 (53.0%) | 0.021 |
| Male | 4,646 (45.9%) | 575,455 (53.1%) | 0.144 | 4,646 (45.9%) | 4,747 (46.9%) | 0.020 |
| Unknown Gender | 10 (0.1%) | 2,691 (0.2%) | 0.036 | 10 (0.1%) | 10 (0.1%) | < 0.001 |
| **Race — no. (%)** |  |  |  |  |  |  |
| American Indian or Alaska Native | 16 (0.2%) | 2,865 (0.3%) | 0.023 | 16 (0.2%) | 15 (0.1%) | 0.003 |
| Asian | 415 (4.1%) | 31,136 (2.9%) | 0.067 | 415 (4.1%) | 435 (4.3%) | 0.010 |
| Black or African American | 1,137 (11.2%) | 187,269 (17.3%) | 0.174 | 1,137 (11.2%) | 1,112 (11.0%) | 0.008 |
| Native Hawaiian or Other Pacific Islander | 27 (0.3%) | 4,403 (0.4%) | 0.024 | 27 (0.3%) | 17 (0.2%) | 0.021 |
| Other Race | 819 (8.1%) | 71,221 (6.6%) | 0.058 | 819 (8.1%) | 801 (7.9%) | 0.007 |
| Unknown Race | 1,015 (10.0%) | 71,420 (6.6%) | 0.125 | 1,015 (10.0%) | 1,049 (10.4%) | 0.011 |
| White | 6,684 (66.1%) | 714,754 (66.0%) | 0.002 | 6,684 (66.1%) | 6,684 (66.1%) | < 0.001 |
| **Ethnicity — no. (%)** |  |  |  |  |  |  |
| Hispanic or Latino | 901 (8.9%) | 100,347 (9.3%) | 0.012 | 901 (8.9%) | 848 (8.4%) | 0.019 |
| Not Hispanic or Latino | 7,839 (77.5%) | 818,331 (75.6%) | 0.046 | 7,839 (77.5%) | 7,929 (78.4%) | 0.021 |
| Unknown Ethnicity | 1,373 (13.6%) | 164,390 (15.2%) | 0.046 | 1,373 (13.6%) | 1,336 (13.2%) | 0.011 |
| **Comorbidities — no. (%)** |  |  |  |  |  |  |
| Essential (primary) hypertension | 3,791 (37.5%) | 490,406 (45.3%) | 0.159 | 3,791 (37.5%) | 3,727 (36.9%) | 0.013 |
| Disorders of lipoprotein metabolism | 3,043 (30.1%) | 369,384 (34.1%) | 0.086 | 3,043 (30.1%) | 2,988 (29.5%) | 0.012 |
| Type 2 diabetes mellitus | 1,393 (13.8%) | 243,983 (22.5%) | 0.229 | 1,393 (13.8%) | 1,377 (13.6%) | 0.005 |
| Chronic ischemic heart disease | 764 (7.6%) | 115,043 (10.6%) | 0.107 | 764 (7.6%) | 751 (7.4%) | 0.005 |
| Chronic kidney disease (CKD) | 375 (3.7%) | 47,525 (4.4%) | 0.035 | 375 (3.7%) | 312 (3.1%) | 0.034 |
| Atrial fibrillation and flutter | 309 (3.1%) | 43,648 (4.0%) | 0.053 | 309 (3.1%) | 308 (3.0%) | < 0.001 |
| Heart failure | 277 (2.7%) | 57,233 (5.3%) | 0.130 | 277 (2.7%) | 248 (2.5%) | 0.018 |
| Acute myocardial infarction | 68 (0.7%) | 30,215 (2.8%) | 0.163 | 68 (0.7%) | 66 (0.7%) | 0.002 |
| Cerebral infarction | 108 (1.1%) | 26,990 (2.5%) | 0.108 | 108 (1.1%) | 109 (1.1%) | < 0.001 |
| Other peripheral vascular diseases | 179 (1.8%) | 31,388 (2.9%) | 0.075 | 179 (1.8%) | 146 (1.4%) | 0.026 |
| Major depressive disorder | 258 (2.6%) | 30,219 (2.8%) | 0.015 | 258 (2.6%) | 233 (2.3%) | 0.016 |
| Other anxiety disorders | 739 (7.3%) | 87,889 (8.1%) | 0.030 | 739 (7.3%) | 711 (7.0%) | 0.011 |
| Mental and behavioral disorders due to psychoactive substance use | 373 (3.7%) | 126,185 (11.7%) | 0.303 | 373 (3.7%) | 383 (3.8%) | 0.005 |
| Alcohol related disorders | 57 (0.6%) | 29,861 (2.8%) | 0.172 | 57 (0.6%) | 69 (0.7%) | 0.015 |
| Nicotine dependence | 276 (2.7%) | 100,539 (9.3%) | 0.278 | 276 (2.7%) | 283 (2.8%) | 0.004 |
| Obesity, unspecified | 508 (5.0%) | 73,429 (6.8%) | 0.075 | 508 (5.0%) | 460 (4.5%) | 0.022 |
| Overweight | 133 (1.3%) | 14,430 (1.3%) | 0.002 | 133 (1.3%) | 124 (1.2%) | 0.008 |
| Problems related to housing and economic circumstances | 21 (0.2%) | 7,527 (0.7%) | 0.073 | 21 (0.2%) | 24 (0.2%) | 0.006 |
| **Medications — no. (%)** |  |  |  |  |  |  |
| HMG CoA reductase inhibitors | 2,085 (20.6%) | 245,062 (22.6%) | 0.049 | 2,085 (20.6%) | 2,105 (20.8%) | 0.005 |
| Beta blocking agents | 1,484 (14.7%) | 169,802 (15.7%) | 0.028 | 1,484 (14.7%) | 1,508 (14.9%) | 0.007 |
| Calcium channel blockers | 994 (9.8%) | 111,215 (10.3%) | 0.015 | 994 (9.8%) | 981 (9.7%) | 0.004 |
| Platelet aggregation inhibitors | 571 (5.6%) | 91,830 (8.5%) | 0.111 | 571 (5.6%) | 567 (5.6%) | 0.002 |
| Direct factor Xa inhibitors | 146 (1.4%) | 16,614 (1.5%) | 0.007 | 146 (1.4%) | 142 (1.4%) | 0.003 |
| Vitamin K antagonists | 117 (1.2%) | 16,263 (1.5%) | 0.030 | 117 (1.2%) | 119 (1.2%) | 0.002 |
| **Proxy measures — no. (%)** |  |  |  |  |  |  |
| Established Patient | 4,702 (46.5%) | 542,663 (50.1%) | 0.072 | 4,702 (46.5%) | 4,600 (45.5%) | 0.020 |
| New Patient | 2,259 (22.3%) | 271,662 (25.1%) | 0.065 | 2,259 (22.3%) | 2,156 (21.3%) | 0.025 |
| Emergency Department Services | 993 (9.8%) | 209,626 (19.4%) | 0.273 | 993 (9.8%) | 979 (9.7%) | 0.005 |
| State Medicaid Agency | 80 (0.8%) | 28,203 (2.6%) | 0.141 | 80 (0.8%) | 83 (0.8%) | 0.003 |

**Table S6B. Irbesartan vs. Lisinopril — baseline characteristics**

| **Characteristic** | **Before PSM** | | | **After PSM** | | |
| --- | --- | --- | --- | --- | --- | --- |
|  | **Irbesartan (N=41,709)** | **Lisinopril (N=1,083,077)** | **SMD** | **Irbesartan (N=41,709)** | **Lisinopril (N=41,709)** | **SMD** |
| Age — Mean (SD) | 64.3 (6.3) | 63.1 (6.0) | 0.208 | 64.3 (6.3) | 64.2 (6.3) | 0.015 |
| **Sex — no. (%)** |  |  |  |  |  |  |
| Female | 21,593 (51.8%) | 504,931 (46.6%) | 0.103 | 21,593 (51.8%) | 21,524 (51.6%) | 0.003 |
| Male | 19,952 (47.8%) | 575,455 (53.1%) | 0.106 | 19,952 (47.8%) | 20,032 (48.0%) | 0.004 |
| Unknown Gender | 164 (0.4%) | 2,691 (0.2%) | 0.026 | 164 (0.4%) | 153 (0.4%) | 0.004 |
| **Race — no. (%)** |  |  |  |  |  |  |
| American Indian or Alaska Native | 96 (0.2%) | 2,865 (0.3%) | 0.007 | 96 (0.2%) | 91 (0.2%) | 0.003 |
| Asian | 2,181 (5.2%) | 31,136 (2.9%) | 0.120 | 2,181 (5.2%) | 2,127 (5.1%) | 0.006 |
| Black or African American | 5,102 (12.2%) | 187,273 (17.3%) | 0.143 | 5,102 (12.2%) | 5,079 (12.2%) | 0.002 |
| Native Hawaiian or Other Pacific Islander | 103 (0.2%) | 4,403 (0.4%) | 0.028 | 103 (0.2%) | 96 (0.2%) | 0.003 |
| Other Race | 2,985 (7.2%) | 71,221 (6.6%) | 0.023 | 2,985 (7.2%) | 2,893 (6.9%) | 0.009 |
| Unknown Race | 3,682 (8.8%) | 71,420 (6.6%) | 0.084 | 3,682 (8.8%) | 3,752 (9.0%) | 0.006 |
| White | 27,560 (66.1%) | 714,759 (66.0%) | 0.002 | 27,560 (66.1%) | 27,671 (66.3%) | 0.006 |
| **Ethnicity — no. (%)** |  |  |  |  |  |  |
| Hispanic or Latino | 3,270 (7.8%) | 100,347 (9.3%) | 0.051 | 3,270 (7.8%) | 3,158 (7.6%) | 0.010 |
| Not Hispanic or Latino | 31,955 (76.6%) | 818,338 (75.6%) | 0.025 | 31,955 (76.6%) | 32,110 (77.0%) | 0.009 |
| Unknown Ethnicity | 6,484 (15.5%) | 164,392 (15.2%) | 0.010 | 6,484 (15.5%) | 6,441 (15.4%) | 0.003 |
| **Comorbidities — no. (%)** |  |  |  |  |  |  |
| Essential (primary) hypertension | 17,158 (41.1%) | 490,411 (45.3%) | 0.084 | 17,158 (41.1%) | 16,600 (39.8%) | 0.027 |
| Disorders of lipoprotein metabolism | 13,322 (31.9%) | 369,387 (34.1%) | 0.046 | 13,322 (31.9%) | 12,892 (30.9%) | 0.022 |
| Type 2 diabetes mellitus | 7,572 (18.2%) | 243,985 (22.5%) | 0.109 | 7,572 (18.2%) | 7,259 (17.4%) | 0.020 |
| Chronic ischemic heart disease | 3,354 (8.0%) | 115,044 (10.6%) | 0.089 | 3,354 (8.0%) | 3,081 (7.4%) | 0.025 |
| Chronic kidney disease (CKD) | 1,900 (4.6%) | 47,525 (4.4%) | 0.008 | 1,900 (4.6%) | 1,647 (3.9%) | 0.030 |
| Atrial fibrillation and flutter | 1,338 (3.2%) | 43,649 (4.0%) | 0.044 | 1,338 (3.2%) | 1,138 (2.7%) | 0.028 |
| Heart failure | 1,085 (2.6%) | 57,234 (5.3%) | 0.138 | 1,085 (2.6%) | 1,010 (2.4%) | 0.011 |
| Acute myocardial infarction | 360 (0.9%) | 30,215 (2.8%) | 0.144 | 360 (0.9%) | 367 (0.9%) | 0.002 |
| Cerebral infarction | 510 (1.2%) | 26,990 (2.5%) | 0.094 | 510 (1.2%) | 442 (1.1%) | 0.015 |
| Other peripheral vascular diseases | 885 (2.1%) | 31,388 (2.9%) | 0.050 | 885 (2.1%) | 753 (1.8%) | 0.023 |
| Major depressive disorder | 820 (2.0%) | 30,219 (2.8%) | 0.054 | 820 (2.0%) | 730 (1.8%) | 0.016 |
| Other anxiety disorders | 2,491 (6.0%) | 87,889 (8.1%) | 0.084 | 2,491 (6.0%) | 2,258 (5.4%) | 0.024 |
| Mental and behavioral disorders due to psychoactive substance use | 1,932 (4.6%) | 126,187 (11.7%) | 0.259 | 1,932 (4.6%) | 1,836 (4.4%) | 0.011 |
| Alcohol related disorders | 388 (0.9%) | 29,862 (2.8%) | 0.136 | 388 (0.9%) | 383 (0.9%) | 0.001 |
| Nicotine dependence | 1,460 (3.5%) | 100,541 (9.3%) | 0.238 | 1,460 (3.5%) | 1,435 (3.4%) | 0.003 |
| Obesity, unspecified | 2,357 (5.7%) | 73,429 (6.8%) | 0.047 | 2,357 (5.7%) | 2,121 (5.1%) | 0.025 |
| Overweight | 540 (1.3%) | 14,430 (1.3%) | 0.003 | 540 (1.3%) | 461 (1.1%) | 0.017 |
| Problems related to housing and economic circumstances | 69 (0.2%) | 7,527 (0.7%) | 0.081 | 69 (0.2%) | 77 (0.2%) | 0.005 |
| **Medications — no. (%)** |  |  |  |  |  |  |
| HMG CoA reductase inhibitors | 7,934 (19.0%) | 245,062 (22.6%) | 0.089 | 7,934 (19.0%) | 7,761 (18.6%) | 0.011 |
| Beta blocking agents | 5,628 (13.5%) | 169,804 (15.7%) | 0.062 | 5,628 (13.5%) | 5,396 (12.9%) | 0.016 |
| Calcium channel blockers | 4,691 (11.2%) | 111,215 (10.3%) | 0.032 | 4,691 (11.2%) | 4,424 (10.6%) | 0.021 |
| Platelet aggregation inhibitors | 2,156 (5.2%) | 91,831 (8.5%) | 0.132 | 2,156 (5.2%) | 2,104 (5.0%) | 0.006 |
| Direct factor Xa inhibitors | 593 (1.4%) | 16,614 (1.5%) | 0.009 | 593 (1.4%) | 479 (1.1%) | 0.024 |
| Vitamin K antagonists | 456 (1.1%) | 16,263 (1.5%) | 0.036 | 456 (1.1%) | 405 (1.0%) | 0.012 |
| **Proxy measures — no. (%)** |  |  |  |  |  |  |
| Established Patient | 19,315 (46.3%) | 542,666 (50.1%) | 0.076 | 19,315 (46.3%) | 18,677 (44.8%) | 0.031 |
| New Patient | 8,819 (21.1%) | 271,664 (25.1%) | 0.094 | 8,819 (21.1%) | 8,478 (20.3%) | 0.020 |
| Emergency Department Services | 4,299 (10.3%) | 209,626 (19.4%) | 0.257 | 4,299 (10.3%) | 4,093 (9.8%) | 0.016 |
| State Medicaid Agency | 445 (1.1%) | 28,203 (2.6%) | 0.115 | 445 (1.1%) | 409 (1.0%) | 0.009 |

**Table S6C. Losartan vs. Lisinopril — baseline characteristics**

| **Characteristic** | **Before PSM** | | | **After PSM** | | |
| --- | --- | --- | --- | --- | --- | --- |
|  | **Losartan (N=583,461)** | **Lisinopril (N=1,083,068)** | **SMD** | **Losartan (N=564,984)** | **Lisinopril (N=564,984)** | **SMD** |
| Age — Mean (SD) | 64.1 (6.4) | 63.1 (6.0) | 0.166 | 63.9 (6.3) | 63.9 (6.3) | 0.006 |
| **Sex — no. (%)** |  |  |  |  |  |  |
| Female | 312,128 (53.5%) | 504,922 (46.6%) | 0.138 | 299,435 (53.0%) | 300,838 (53.2%) | 0.005 |
| Male | 270,748 (46.4%) | 575,455 (53.1%) | 0.135 | 264,964 (46.9%) | 263,518 (46.6%) | 0.005 |
| Unknown Gender | 585 (0.1%) | 2,691 (0.2%) | 0.036 | 585 (0.1%) | 628 (0.1%) | 0.002 |
| **Race — no. (%)** |  |  |  |  |  |  |
| American Indian or Alaska Native | 1,626 (0.3%) | 2,865 (0.3%) | 0.003 | 1,619 (0.3%) | 1,601 (0.3%) | < 0.001 |
| Asian | 46,373 (7.9%) | 31,136 (2.9%) | 0.226 | 30,409 (5.4%) | 30,663 (5.4%) | 0.002 |
| Black or African American | 99,656 (17.1%) | 187,269 (17.3%) | 0.006 | 99,142 (17.5%) | 96,727 (17.1%) | 0.011 |
| Native Hawaiian or Other Pacific Islander | 2,505 (0.4%) | 4,403 (0.4%) | 0.004 | 2,488 (0.4%) | 2,386 (0.4%) | 0.003 |
| Other Race | 49,155 (8.4%) | 71,221 (6.6%) | 0.070 | 48,203 (8.5%) | 47,906 (8.5%) | 0.002 |
| Unknown Race | 42,695 (7.3%) | 71,420 (6.6%) | 0.028 | 42,342 (7.5%) | 42,546 (7.5%) | 0.001 |
| White | 341,451 (58.5%) | 714,754 (66.0%) | 0.155 | 340,781 (60.3%) | 343,155 (60.7%) | 0.009 |
| **Ethnicity — no. (%)** |  |  |  |  |  |  |
| Hispanic or Latino | 58,774 (10.1%) | 100,347 (9.3%) | 0.027 | 57,980 (10.3%) | 58,109 (10.3%) | < 0.001 |
| Not Hispanic or Latino | 444,917 (76.3%) | 818,331 (75.6%) | 0.016 | 428,063 (75.8%) | 425,803 (75.4%) | 0.009 |
| Unknown Ethnicity | 79,770 (13.7%) | 164,390 (15.2%) | 0.043 | 78,941 (14.0%) | 81,072 (14.3%) | 0.011 |
| **Comorbidities — no. (%)** |  |  |  |  |  |  |
| Essential (primary) hypertension | 275,406 (47.2%) | 490,406 (45.3%) | 0.039 | 265,091 (46.9%) | 262,123 (46.4%) | 0.011 |
| Disorders of lipoprotein metabolism | 213,152 (36.5%) | 369,384 (34.1%) | 0.051 | 204,008 (36.1%) | 200,666 (35.5%) | 0.012 |
| Type 2 diabetes mellitus | 122,348 (21.0%) | 243,983 (22.5%) | 0.038 | 118,593 (21.0%) | 116,194 (20.6%) | 0.010 |
| Chronic ischemic heart disease | 56,695 (9.7%) | 115,043 (10.6%) | 0.030 | 55,181 (9.8%) | 52,750 (9.3%) | 0.015 |
| Chronic kidney disease (CKD) | 32,454 (5.6%) | 47,525 (4.4%) | 0.054 | 30,498 (5.4%) | 29,579 (5.2%) | 0.007 |
| Atrial fibrillation and flutter | 23,354 (4.0%) | 43,648 (4.0%) | 0.001 | 22,505 (4.0%) | 21,410 (3.8%) | 0.010 |
| Heart failure | 26,726 (4.6%) | 57,233 (5.3%) | 0.033 | 26,175 (4.6%) | 24,674 (4.4%) | 0.013 |
| Acute myocardial infarction | 10,533 (1.8%) | 30,215 (2.8%) | 0.066 | 10,474 (1.9%) | 9,819 (1.7%) | 0.009 |
| Cerebral infarction | 11,789 (2.0%) | 26,990 (2.5%) | 0.032 | 11,569 (2.0%) | 10,677 (1.9%) | 0.011 |
| Other peripheral vascular diseases | 17,749 (3.0%) | 31,388 (2.9%) | 0.008 | 17,004 (3.0%) | 15,724 (2.8%) | 0.014 |
| Major depressive disorder | 17,275 (3.0%) | 30,219 (2.8%) | 0.010 | 16,832 (3.0%) | 15,954 (2.8%) | 0.009 |
| Other anxiety disorders | 49,557 (8.5%) | 87,889 (8.1%) | 0.014 | 48,518 (8.6%) | 47,029 (8.3%) | 0.009 |
| Mental and behavioral disorders due to psychoactive substance use | 52,838 (9.1%) | 126,185 (11.7%) | 0.085 | 52,364 (9.3%) | 49,733 (8.8%) | 0.016 |
| Alcohol related disorders | 11,689 (2.0%) | 29,861 (2.8%) | 0.049 | 11,642 (2.1%) | 10,773 (1.9%) | 0.011 |
| Nicotine dependence | 41,669 (7.1%) | 100,539 (9.3%) | 0.078 | 41,287 (7.3%) | 39,437 (7.0%) | 0.013 |
| Obesity, unspecified | 43,351 (7.4%) | 73,429 (6.8%) | 0.025 | 42,306 (7.5%) | 40,877 (7.2%) | 0.010 |
| Overweight | 10,361 (1.8%) | 14,430 (1.3%) | 0.036 | 9,599 (1.7%) | 9,149 (1.6%) | 0.006 |
| Problems related to housing and economic circumstances | 3,740 (0.6%) | 7,527 (0.7%) | 0.007 | 3,643 (0.6%) | 3,342 (0.6%) | 0.007 |
| **Medications — no. (%)** |  |  |  |  |  |  |
| HMG CoA reductase inhibitors | 136,788 (23.4%) | 245,062 (22.6%) | 0.019 | 131,514 (23.3%) | 129,359 (22.9%) | 0.009 |
| Beta blocking agents | 99,907 (17.1%) | 169,802 (15.7%) | 0.039 | 95,878 (17.0%) | 94,613 (16.7%) | 0.006 |
| Calcium channel blockers | 85,033 (14.6%) | 111,215 (10.3%) | 0.131 | 79,239 (14.0%) | 78,576 (13.9%) | 0.003 |
| Platelet aggregation inhibitors | 47,181 (8.1%) | 91,830 (8.5%) | 0.014 | 45,490 (8.1%) | 43,309 (7.7%) | 0.014 |
| Direct factor Xa inhibitors | 12,597 (2.2%) | 16,614 (1.5%) | 0.046 | 11,677 (2.1%) | 11,208 (2.0%) | 0.006 |
| Vitamin K antagonists | 6,073 (1.0%) | 16,263 (1.5%) | 0.041 | 6,034 (1.1%) | 5,851 (1.0%) | 0.003 |
| **Proxy measures — no. (%)** |  |  |  |  |  |  |
| Established Patient | 300,145 (51.4%) | 542,663 (50.1%) | 0.027 | 289,829 (51.3%) | 288,388 (51.0%) | 0.005 |
| New Patient | 160,801 (27.6%) | 271,662 (25.1%) | 0.056 | 154,211 (27.3%) | 152,625 (27.0%) | 0.006 |
| Emergency Department Services | 100,706 (17.3%) | 209,626 (19.4%) | 0.054 | 98,814 (17.5%) | 96,734 (17.1%) | 0.010 |
| State Medicaid Agency | 15,319 (2.6%) | 28,203 (2.6%) | 0.001 | 14,649 (2.6%) | 13,618 (2.4%) | 0.012 |

**Table S6D. Olmesartan vs. Lisinopril — baseline characteristics**

| **Characteristic** | **Before PSM** | | | **After PSM** | | |
| --- | --- | --- | --- | --- | --- | --- |
|  | **Olmesartan (N=84,958)** | **Lisinopril (N=1,083,068)** | **SMD** | **Olmesartan (N=84,957)** | **Lisinopril (N=84,957)** | **SMD** |
| Age — Mean (SD) | 63.5 (6.2) | 63.1 (6.0) | 0.078 | 63.5 (6.2) | 63.4 (6.1) | 0.022 |
| **Sex — no. (%)** |  |  |  |  |  |  |
| Female | 43,737 (51.5%) | 504,922 (46.6%) | 0.097 | 43,736 (51.5%) | 43,449 (51.1%) | 0.007 |
| Male | 41,157 (48.4%) | 575,455 (53.1%) | 0.094 | 41,157 (48.4%) | 41,419 (48.8%) | 0.006 |
| Unknown Gender | 64 (0.1%) | 2,691 (0.2%) | 0.043 | 64 (0.1%) | 89 (0.1%) | 0.010 |
| **Race — no. (%)** |  |  |  |  |  |  |
| American Indian or Alaska Native | 200 (0.2%) | 2,865 (0.3%) | 0.006 | 200 (0.2%) | 205 (0.2%) | 0.001 |
| Asian | 5,164 (6.1%) | 31,136 (2.9%) | 0.155 | 5,163 (6.1%) | 5,101 (6.0%) | 0.003 |
| Black or African American | 13,715 (16.1%) | 187,269 (17.3%) | 0.031 | 13,715 (16.1%) | 13,542 (15.9%) | 0.006 |
| Native Hawaiian or Other Pacific Islander | 167 (0.2%) | 4,403 (0.4%) | 0.038 | 167 (0.2%) | 182 (0.2%) | 0.004 |
| Other Race | 6,235 (7.3%) | 71,221 (6.6%) | 0.030 | 6,235 (7.3%) | 6,086 (7.2%) | 0.007 |
| Unknown Race | 7,640 (9.0%) | 71,420 (6.6%) | 0.090 | 7,640 (9.0%) | 7,781 (9.2%) | 0.006 |
| White | 51,837 (61.0%) | 714,754 (66.0%) | 0.104 | 51,837 (61.0%) | 52,060 (61.3%) | 0.005 |
| **Ethnicity — no. (%)** |  |  |  |  |  |  |
| Hispanic or Latino | 7,967 (9.4%) | 100,347 (9.3%) | 0.004 | 7,967 (9.4%) | 7,581 (8.9%) | 0.016 |
| Not Hispanic or Latino | 63,091 (74.3%) | 818,331 (75.6%) | 0.030 | 63,090 (74.3%) | 63,474 (74.7%) | 0.010 |
| Unknown Ethnicity | 13,900 (16.4%) | 164,390 (15.2%) | 0.032 | 13,900 (16.4%) | 13,902 (16.4%) | < 0.001 |
| **Comorbidities — no. (%)** |  |  |  |  |  |  |
| Essential (primary) hypertension | 35,788 (42.1%) | 490,406 (45.3%) | 0.064 | 35,787 (42.1%) | 34,816 (41.0%) | 0.023 |
| Disorders of lipoprotein metabolism | 26,498 (31.2%) | 369,384 (34.1%) | 0.062 | 26,497 (31.2%) | 25,418 (29.9%) | 0.028 |
| Type 2 diabetes mellitus | 12,944 (15.2%) | 243,983 (22.5%) | 0.187 | 12,944 (15.2%) | 12,403 (14.6%) | 0.018 |
| Chronic ischemic heart disease | 5,461 (6.4%) | 115,043 (10.6%) | 0.151 | 5,461 (6.4%) | 4,979 (5.9%) | 0.024 |
| Chronic kidney disease (CKD) | 3,044 (3.6%) | 47,525 (4.4%) | 0.041 | 3,043 (3.6%) | 2,516 (3.0%) | 0.035 |
| Atrial fibrillation and flutter | 1,998 (2.4%) | 43,648 (4.0%) | 0.096 | 1,998 (2.4%) | 1,725 (2.0%) | 0.022 |
| Heart failure | 1,485 (1.7%) | 57,233 (5.3%) | 0.193 | 1,485 (1.7%) | 1,342 (1.6%) | 0.013 |
| Acute myocardial infarction | 418 (0.5%) | 30,215 (2.8%) | 0.182 | 418 (0.5%) | 476 (0.6%) | 0.009 |
| Cerebral infarction | 899 (1.1%) | 26,990 (2.5%) | 0.109 | 899 (1.1%) | 866 (1.0%) | 0.004 |
| Other peripheral vascular diseases | 1,686 (2.0%) | 31,388 (2.9%) | 0.059 | 1,686 (2.0%) | 1,404 (1.7%) | 0.025 |
| Major depressive disorder | 1,675 (2.0%) | 30,219 (2.8%) | 0.054 | 1,675 (2.0%) | 1,465 (1.7%) | 0.018 |
| Other anxiety disorders | 5,358 (6.3%) | 87,889 (8.1%) | 0.070 | 5,358 (6.3%) | 4,802 (5.7%) | 0.028 |
| Mental and behavioral disorders due to psychoactive substance use | 4,426 (5.2%) | 126,185 (11.7%) | 0.233 | 4,426 (5.2%) | 4,108 (4.8%) | 0.017 |
| Alcohol related disorders | 895 (1.1%) | 29,861 (2.8%) | 0.125 | 895 (1.1%) | 749 (0.9%) | 0.018 |
| Nicotine dependence | 3,360 (4.0%) | 100,539 (9.3%) | 0.216 | 3,360 (4.0%) | 3,201 (3.8%) | 0.010 |
| Obesity, unspecified | 4,759 (5.6%) | 73,429 (6.8%) | 0.049 | 4,759 (5.6%) | 4,390 (5.2%) | 0.019 |
| Overweight | 1,199 (1.4%) | 14,430 (1.3%) | 0.007 | 1,198 (1.4%) | 1,004 (1.2%) | 0.020 |
| Problems related to housing and economic circumstances | 195 (0.2%) | 7,527 (0.7%) | 0.069 | 195 (0.2%) | 177 (0.2%) | 0.005 |
| **Medications — no. (%)** |  |  |  |  |  |  |
| HMG CoA reductase inhibitors | 16,849 (19.8%) | 245,062 (22.6%) | 0.068 | 16,849 (19.8%) | 16,321 (19.2%) | 0.016 |
| Beta blocking agents | 11,048 (13.0%) | 169,802 (15.7%) | 0.076 | 11,048 (13.0%) | 10,481 (12.3%) | 0.020 |
| Calcium channel blockers | 9,975 (11.7%) | 111,215 (10.3%) | 0.047 | 9,974 (11.7%) | 9,379 (11.0%) | 0.022 |
| Platelet aggregation inhibitors | 4,260 (5.0%) | 91,830 (8.5%) | 0.138 | 4,260 (5.0%) | 4,013 (4.7%) | 0.014 |
| Direct factor Xa inhibitors | 1,107 (1.3%) | 16,614 (1.5%) | 0.020 | 1,107 (1.3%) | 894 (1.1%) | 0.023 |
| Vitamin K antagonists | 561 (0.7%) | 16,263 (1.5%) | 0.081 | 561 (0.7%) | 533 (0.6%) | 0.004 |
| **Proxy measures — no. (%)** |  |  |  |  |  |  |
| Established Patient | 39,865 (46.9%) | 542,663 (50.1%) | 0.064 | 39,864 (46.9%) | 38,874 (45.8%) | 0.023 |
| New Patient | 19,043 (22.4%) | 271,662 (25.1%) | 0.063 | 19,042 (22.4%) | 18,144 (21.4%) | 0.026 |
| Emergency Department Services | 9,235 (10.9%) | 209,626 (19.4%) | 0.239 | 9,235 (10.9%) | 9,066 (10.7%) | 0.006 |
| State Medicaid Agency | 1,201 (1.4%) | 28,203 (2.6%) | 0.085 | 1,201 (1.4%) | 1,089 (1.3%) | 0.011 |

**Table S6E. Telmisartan vs. Lisinopril — baseline characteristics**

| **Characteristic** | **Before PSM** | | | **After PSM** | | |
| --- | --- | --- | --- | --- | --- | --- |
|  | **Telmisartan (N=24,295)** | **Lisinopril (N=1,083,068)** | **SMD** | **Telmisartan (N=24,294)** | **Lisinopril (N=24,294)** | **SMD** |
| Age — Mean (SD) | 64.1 (6.2) | 63.1 (6.0) | 0.173 | 64.1 (6.2) | 64.1 (6.2) | 0.008 |
| **Sex — no. (%)** |  |  |  |  |  |  |
| Female | 13,074 (53.8%) | 504,922 (46.6%) | 0.144 | 13,073 (53.8%) | 13,074 (53.8%) | < 0.001 |
| Male | 11,191 (46.1%) | 575,455 (53.1%) | 0.142 | 11,191 (46.1%) | 11,177 (46.0%) | 0.001 |
| Unknown Gender | 30 (0.1%) | 2,691 (0.2%) | 0.029 | 30 (0.1%) | 43 (0.2%) | 0.014 |
| **Race — no. (%)** |  |  |  |  |  |  |
| American Indian or Alaska Native | 91 (0.4%) | 2,865 (0.3%) | 0.019 | 91 (0.4%) | 81 (0.3%) | 0.007 |
| Asian | 2,126 (8.8%) | 31,136 (2.9%) | 0.253 | 2,125 (8.7%) | 2,139 (8.8%) | 0.002 |
| Black or African American | 3,539 (14.6%) | 187,269 (17.3%) | 0.074 | 3,539 (14.6%) | 3,580 (14.7%) | 0.005 |
| Native Hawaiian or Other Pacific Islander | 58 (0.2%) | 4,403 (0.4%) | 0.030 | 58 (0.2%) | 50 (0.2%) | 0.007 |
| Other Race | 2,219 (9.1%) | 71,221 (6.6%) | 0.095 | 2,219 (9.1%) | 2,198 (9.0%) | 0.003 |
| Unknown Race | 2,497 (10.3%) | 71,420 (6.6%) | 0.133 | 2,497 (10.3%) | 2,459 (10.1%) | 0.005 |
| White | 13,765 (56.7%) | 714,754 (66.0%) | 0.193 | 13,765 (56.7%) | 13,787 (56.8%) | 0.002 |
| **Ethnicity — no. (%)** |  |  |  |  |  |  |
| Hispanic or Latino | 3,049 (12.5%) | 100,347 (9.3%) | 0.106 | 3,049 (12.6%) | 2,911 (12.0%) | 0.017 |
| Not Hispanic or Latino | 17,742 (73.0%) | 818,331 (75.6%) | 0.058 | 17,741 (73.0%) | 17,931 (73.8%) | 0.018 |
| Unknown Ethnicity | 3,504 (14.4%) | 164,390 (15.2%) | 0.021 | 3,504 (14.4%) | 3,452 (14.2%) | 0.006 |
| **Comorbidities — no. (%)** |  |  |  |  |  |  |
| Essential (primary) hypertension | 10,081 (41.5%) | 490,406 (45.3%) | 0.076 | 10,080 (41.5%) | 9,769 (40.2%) | 0.026 |
| Disorders of lipoprotein metabolism | 7,669 (31.6%) | 369,384 (34.1%) | 0.054 | 7,668 (31.6%) | 7,338 (30.2%) | 0.029 |
| Type 2 diabetes mellitus | 4,168 (17.2%) | 243,983 (22.5%) | 0.135 | 4,168 (17.2%) | 3,946 (16.2%) | 0.025 |
| Chronic ischemic heart disease | 1,685 (6.9%) | 115,043 (10.6%) | 0.131 | 1,685 (6.9%) | 1,540 (6.3%) | 0.024 |
| Chronic kidney disease (CKD) | 962 (4.0%) | 47,525 (4.4%) | 0.021 | 961 (4.0%) | 860 (3.5%) | 0.022 |
| Atrial fibrillation and flutter | 558 (2.3%) | 43,648 (4.0%) | 0.099 | 557 (2.3%) | 506 (2.1%) | 0.014 |
| Heart failure | 433 (1.8%) | 57,233 (5.3%) | 0.191 | 433 (1.8%) | 398 (1.6%) | 0.011 |
| Acute myocardial infarction | 147 (0.6%) | 30,215 (2.8%) | 0.170 | 147 (0.6%) | 159 (0.7%) | 0.006 |
| Cerebral infarction | 219 (0.9%) | 26,990 (2.5%) | 0.123 | 219 (0.9%) | 220 (0.9%) | < 0.001 |
| Other peripheral vascular diseases | 516 (2.1%) | 31,388 (2.9%) | 0.049 | 516 (2.1%) | 484 (2.0%) | 0.009 |
| Major depressive disorder | 492 (2.0%) | 30,219 (2.8%) | 0.050 | 492 (2.0%) | 422 (1.7%) | 0.021 |
| Other anxiety disorders | 1,351 (5.6%) | 87,889 (8.1%) | 0.101 | 1,351 (5.6%) | 1,151 (4.7%) | 0.037 |
| Mental and behavioral disorders due to psychoactive substance use | 984 (4.1%) | 126,185 (11.7%) | 0.285 | 984 (4.1%) | 947 (3.9%) | 0.008 |
| Alcohol related disorders | 201 (0.8%) | 29,861 (2.8%) | 0.146 | 201 (0.8%) | 179 (0.7%) | 0.010 |
| Nicotine dependence | 736 (3.0%) | 100,539 (9.3%) | 0.262 | 736 (3.0%) | 739 (3.0%) | < 0.001 |
| Obesity, unspecified | 1,303 (5.4%) | 73,429 (6.8%) | 0.059 | 1,303 (5.4%) | 1,186 (4.9%) | 0.022 |
| Overweight | 374 (1.5%) | 14,430 (1.3%) | 0.017 | 374 (1.5%) | 311 (1.3%) | 0.022 |
| Problems related to housing and economic circumstances | 45 (0.2%) | 7,527 (0.7%) | 0.077 | 45 (0.2%) | 51 (0.2%) | 0.006 |
| **Medications — no. (%)** |  |  |  |  |  |  |
| HMG CoA reductase inhibitors | 5,025 (20.7%) | 245,062 (22.6%) | 0.047 | 5,025 (20.7%) | 4,902 (20.2%) | 0.013 |
| Beta blocking agents | 3,185 (13.1%) | 169,802 (15.7%) | 0.073 | 3,184 (13.1%) | 3,020 (12.4%) | 0.020 |
| Calcium channel blockers | 2,900 (11.9%) | 111,215 (10.3%) | 0.053 | 2,899 (11.9%) | 2,765 (11.4%) | 0.017 |
| Platelet aggregation inhibitors | 1,253 (5.2%) | 91,830 (8.5%) | 0.132 | 1,253 (5.2%) | 1,219 (5.0%) | 0.006 |
| Direct factor Xa inhibitors | 294 (1.2%) | 16,614 (1.5%) | 0.028 | 293 (1.2%) | 252 (1.0%) | 0.016 |
| Vitamin K antagonists | 163 (0.7%) | 16,263 (1.5%) | 0.080 | 163 (0.7%) | 152 (0.6%) | 0.006 |
| **Proxy measures — no. (%)** |  |  |  |  |  |  |
| Established Patient | 10,959 (45.1%) | 542,663 (50.1%) | 0.100 | 10,959 (45.1%) | 10,563 (43.5%) | 0.033 |
| New Patient | 5,432 (22.4%) | 271,662 (25.1%) | 0.064 | 5,432 (22.4%) | 5,049 (20.8%) | 0.038 |
| Emergency Department Services | 2,369 (9.8%) | 209,626 (19.4%) | 0.275 | 2,369 (9.8%) | 2,276 (9.4%) | 0.013 |
| State Medicaid Agency | 231 (1.0%) | 28,203 (2.6%) | 0.125 | 231 (1.0%) | 208 (0.9%) | 0.010 |

**Table S6F. Valsartan vs. Lisinopril — baseline characteristics**

| **Characteristic** | **Before PSM** | | | **After PSM** | | |
| --- | --- | --- | --- | --- | --- | --- |
|  | **Valsartan (N=202,930)** | **Lisinopril (N=1,083,068)** | **SMD** | **Valsartan (N=202,917)** | **Lisinopril (N=202,917)** | **SMD** |
| Age — Mean (SD) | 64.2 (6.3) | 63.1 (6.0) | 0.193 | 64.2 (6.3) | 64.2 (6.3) | 0.005 |
| **Sex — no. (%)** |  |  |  |  |  |  |
| Female | 105,117 (51.8%) | 504,922 (46.6%) | 0.104 | 105,107 (51.8%) | 105,959 (52.2%) | 0.008 |
| Male | 97,406 (48.0%) | 575,455 (53.1%) | 0.103 | 97,403 (48.0%) | 96,583 (47.6%) | 0.008 |
| Unknown Gender | 407 (0.2%) | 2,691 (0.2%) | 0.010 | 407 (0.2%) | 375 (0.2%) | 0.004 |
| **Race — no. (%)** |  |  |  |  |  |  |
| American Indian or Alaska Native | 501 (0.2%) | 2,865 (0.3%) | 0.003 | 501 (0.2%) | 470 (0.2%) | 0.003 |
| Asian | 9,935 (4.9%) | 31,136 (2.9%) | 0.105 | 9,927 (4.9%) | 10,191 (5.0%) | 0.006 |
| Black or African American | 36,668 (18.1%) | 187,269 (17.3%) | 0.020 | 36,668 (18.1%) | 36,312 (17.9%) | 0.005 |
| Native Hawaiian or Other Pacific Islander | 418 (0.2%) | 4,403 (0.4%) | 0.036 | 418 (0.2%) | 410 (0.2%) | < 0.001 |
| Other Race | 16,383 (8.1%) | 71,221 (6.6%) | 0.057 | 16,382 (8.1%) | 16,312 (8.0%) | 0.001 |
| Unknown Race | 16,322 (8.0%) | 71,420 (6.6%) | 0.056 | 16,320 (8.0%) | 16,487 (8.1%) | 0.003 |
| White | 122,703 (60.5%) | 714,754 (66.0%) | 0.115 | 122,701 (60.5%) | 122,735 (60.5%) | < 0.001 |
| **Ethnicity — no. (%)** |  |  |  |  |  |  |
| Hispanic or Latino | 17,471 (8.6%) | 100,347 (9.3%) | 0.023 | 17,471 (8.6%) | 16,665 (8.2%) | 0.014 |
| Not Hispanic or Latino | 154,183 (76.0%) | 818,331 (75.6%) | 0.010 | 154,170 (76.0%) | 154,620 (76.2%) | 0.005 |
| Unknown Ethnicity | 31,276 (15.4%) | 164,390 (15.2%) | 0.007 | 31,276 (15.4%) | 31,632 (15.6%) | 0.005 |
| **Comorbidities — no. (%)** |  |  |  |  |  |  |
| Essential (primary) hypertension | 85,291 (42.0%) | 490,406 (45.3%) | 0.066 | 85,285 (42.0%) | 84,266 (41.5%) | 0.010 |
| Disorders of lipoprotein metabolism | 64,880 (32.0%) | 369,384 (34.1%) | 0.045 | 64,871 (32.0%) | 63,084 (31.1%) | 0.019 |
| Type 2 diabetes mellitus | 37,910 (18.7%) | 243,983 (22.5%) | 0.095 | 37,907 (18.7%) | 36,625 (18.0%) | 0.016 |
| Chronic ischemic heart disease | 23,544 (11.6%) | 115,043 (10.6%) | 0.031 | 23,534 (11.6%) | 21,131 (10.4%) | 0.038 |
| Chronic kidney disease (CKD) | 10,498 (5.2%) | 47,525 (4.4%) | 0.037 | 10,494 (5.2%) | 9,315 (4.6%) | 0.027 |
| Atrial fibrillation and flutter | 10,517 (5.2%) | 43,648 (4.0%) | 0.055 | 10,506 (5.2%) | 8,982 (4.4%) | 0.035 |
| Heart failure | 16,411 (8.1%) | 57,233 (5.3%) | 0.112 | 16,398 (8.1%) | 14,070 (6.9%) | 0.044 |
| Acute myocardial infarction | 5,120 (2.5%) | 30,215 (2.8%) | 0.017 | 5,119 (2.5%) | 4,374 (2.2%) | 0.024 |
| Cerebral infarction | 3,507 (1.7%) | 26,990 (2.5%) | 0.053 | 3,507 (1.7%) | 3,002 (1.5%) | 0.020 |
| Other peripheral vascular diseases | 5,797 (2.9%) | 31,388 (2.9%) | 0.002 | 5,796 (2.9%) | 4,992 (2.5%) | 0.025 |
| Major depressive disorder | 4,033 (2.0%) | 30,219 (2.8%) | 0.053 | 4,033 (2.0%) | 3,604 (1.8%) | 0.016 |
| Other anxiety disorders | 12,896 (6.4%) | 87,889 (8.1%) | 0.068 | 12,894 (6.4%) | 11,709 (5.8%) | 0.024 |
| Mental and behavioral disorders due to psychoactive substance use | 14,261 (7.0%) | 126,185 (11.7%) | 0.159 | 14,261 (7.0%) | 12,671 (6.2%) | 0.031 |
| Alcohol related disorders | 3,222 (1.6%) | 29,861 (2.8%) | 0.080 | 3,222 (1.6%) | 2,820 (1.4%) | 0.016 |
| Nicotine dependence | 11,239 (5.5%) | 100,539 (9.3%) | 0.143 | 11,239 (5.5%) | 10,045 (5.0%) | 0.026 |
| Obesity, unspecified | 12,596 (6.2%) | 73,429 (6.8%) | 0.023 | 12,592 (6.2%) | 11,604 (5.7%) | 0.021 |
| Overweight | 2,368 (1.2%) | 14,430 (1.3%) | 0.015 | 2,368 (1.2%) | 2,096 (1.0%) | 0.013 |
| Problems related to housing and economic circumstances | 832 (0.4%) | 7,527 (0.7%) | 0.038 | 832 (0.4%) | 697 (0.3%) | 0.011 |
| **Medications — no. (%)** |  |  |  |  |  |  |
| HMG CoA reductase inhibitors | 43,040 (21.2%) | 245,062 (22.6%) | 0.034 | 43,033 (21.2%) | 41,424 (20.4%) | 0.020 |
| Beta blocking agents | 35,365 (17.4%) | 169,802 (15.7%) | 0.047 | 35,354 (17.4%) | 33,072 (16.3%) | 0.030 |
| Calcium channel blockers | 24,637 (12.1%) | 111,215 (10.3%) | 0.059 | 24,628 (12.1%) | 23,416 (11.5%) | 0.018 |
| Platelet aggregation inhibitors | 17,037 (8.4%) | 91,830 (8.5%) | 0.003 | 17,031 (8.4%) | 15,470 (7.6%) | 0.028 |
| Direct factor Xa inhibitors | 5,217 (2.6%) | 16,614 (1.5%) | 0.073 | 5,207 (2.6%) | 4,272 (2.1%) | 0.031 |
| Vitamin K antagonists | 2,884 (1.4%) | 16,263 (1.5%) | 0.007 | 2,884 (1.4%) | 2,537 (1.3%) | 0.015 |
| **Proxy measures — no. (%)** |  |  |  |  |  |  |
| Established Patient | 93,064 (45.9%) | 542,663 (50.1%) | 0.085 | 93,057 (45.9%) | 91,543 (45.1%) | 0.015 |
| New Patient | 42,914 (21.1%) | 271,662 (25.1%) | 0.093 | 42,908 (21.1%) | 40,838 (20.1%) | 0.025 |
| Emergency Department Services | 28,472 (14.0%) | 209,626 (19.4%) | 0.143 | 28,470 (14.0%) | 26,294 (13.0%) | 0.031 |
| State Medicaid Agency | 3,195 (1.6%) | 28,203 (2.6%) | 0.072 | 3,195 (1.6%) | 2,781 (1.4%) | 0.017 |

**Table S6G. Pooled ARBs vs. Lisinopril — baseline characteristics**

| **Characteristic** | **Before PSM** | | | **After PSM** | | |
| --- | --- | --- | --- | --- | --- | --- |
|  | **Pooled ARBs (N=942,275)** | **Lisinopril (N=1,083,068)** | **SMD** | **Pooled ARBs (N=857,773)** | **Lisinopril (N=857,773)** | **SMD** |
| Age — Mean (SD) | 64.1 (6.3) | 63.1 (6.0) | 0.165 | 63.7 (6.2) | 63.6 (6.2) | 0.004 |
| **Sex — no. (%)** |  |  |  |  |  |  |
| Female | 498,276 (52.9%) | 504,922 (46.6%) | 0.125 | 438,786 (51.2%) | 438,698 (51.1%) | < 0.001 |
| Male | 442,748 (47.0%) | 575,455 (53.1%) | 0.123 | 417,738 (48.7%) | 417,823 (48.7%) | < 0.001 |
| Unknown Gender | 1,251 (0.1%) | 2,691 (0.2%) | 0.027 | 1,249 (0.1%) | 1,252 (0.1%) | < 0.001 |
| **Race — no. (%)** |  |  |  |  |  |  |
| American Indian or Alaska Native | 2,510 (0.3%) | 2,865 (0.3%) | < 0.001 | 2,363 (0.3%) | 2,248 (0.3%) | 0.003 |
| Asian | 65,932 (7.0%) | 31,136 (2.9%) | 0.191 | 30,601 (3.6%) | 31,048 (3.6%) | 0.003 |
| Black or African American | 158,821 (16.9%) | 187,269 (17.3%) | 0.012 | 149,499 (17.4%) | 146,356 (17.1%) | 0.010 |
| Native Hawaiian or Other Pacific Islander | 3,267 (0.3%) | 4,403 (0.4%) | 0.010 | 3,169 (0.4%) | 3,033 (0.4%) | 0.003 |
| Other Race | 77,594 (8.2%) | 71,221 (6.6%) | 0.063 | 65,747 (7.7%) | 65,107 (7.6%) | 0.003 |
| Unknown Race | 73,694 (7.8%) | 71,420 (6.6%) | 0.047 | 64,797 (7.6%) | 63,460 (7.4%) | 0.006 |
| White | 560,457 (59.5%) | 714,754 (66.0%) | 0.135 | 541,597 (63.1%) | 546,521 (63.7%) | 0.012 |
| **Ethnicity — no. (%)** |  |  |  |  |  |  |
| Hispanic or Latino | 91,097 (9.7%) | 100,347 (9.3%) | 0.014 | 83,033 (9.7%) | 82,781 (9.7%) | < 0.001 |
| Not Hispanic or Latino | 716,647 (76.1%) | 818,331 (75.6%) | 0.012 | 647,424 (75.5%) | 644,476 (75.1%) | 0.008 |
| Unknown Ethnicity | 134,531 (14.3%) | 164,390 (15.2%) | 0.025 | 127,316 (14.8%) | 130,516 (15.2%) | 0.010 |
| **Comorbidities — no. (%)** |  |  |  |  |  |  |
| Essential (primary) hypertension | 425,520 (45.2%) | 490,406 (45.3%) | 0.002 | 381,962 (44.5%) | 381,139 (44.4%) | 0.002 |
| Disorders of lipoprotein metabolism | 327,181 (34.7%) | 369,384 (34.1%) | 0.013 | 291,272 (34.0%) | 289,740 (33.8%) | 0.004 |
| Type 2 diabetes mellitus | 185,510 (19.7%) | 243,983 (22.5%) | 0.070 | 172,208 (20.1%) | 169,595 (19.8%) | 0.008 |
| Chronic ischemic heart disease | 90,977 (9.7%) | 115,043 (10.6%) | 0.032 | 82,929 (9.7%) | 80,954 (9.4%) | 0.008 |
| Chronic kidney disease (CKD) | 48,950 (5.2%) | 47,525 (4.4%) | 0.038 | 40,801 (4.8%) | 39,970 (4.7%) | 0.005 |
| Atrial fibrillation and flutter | 37,836 (4.0%) | 43,648 (4.0%) | < 0.001 | 33,276 (3.9%) | 32,463 (3.8%) | 0.005 |
| Heart failure | 46,069 (4.9%) | 57,233 (5.3%) | 0.018 | 41,769 (4.9%) | 40,472 (4.7%) | 0.007 |
| Acute myocardial infarction | 16,494 (1.8%) | 30,215 (2.8%) | 0.070 | 16,104 (1.9%) | 15,526 (1.8%) | 0.005 |
| Cerebral infarction | 16,944 (1.8%) | 26,990 (2.5%) | 0.048 | 16,199 (1.9%) | 15,729 (1.8%) | 0.004 |
| Other peripheral vascular diseases | 26,690 (2.8%) | 31,388 (2.9%) | 0.004 | 23,619 (2.8%) | 22,728 (2.6%) | 0.006 |
| Major depressive disorder | 24,479 (2.6%) | 30,219 (2.8%) | 0.012 | 22,521 (2.6%) | 21,884 (2.6%) | 0.005 |
| Other anxiety disorders | 72,072 (7.6%) | 87,889 (8.1%) | 0.017 | 66,335 (7.7%) | 65,023 (7.6%) | 0.006 |
| Mental and behavioral disorders due to psychoactive substance use | 74,494 (7.9%) | 126,185 (11.7%) | 0.126 | 72,858 (8.5%) | 70,557 (8.2%) | 0.010 |
| Alcohol related disorders | 16,377 (1.7%) | 29,861 (2.8%) | 0.069 | 16,172 (1.9%) | 15,461 (1.8%) | 0.006 |
| Nicotine dependence | 58,494 (6.2%) | 100,539 (9.3%) | 0.115 | 57,236 (6.7%) | 55,576 (6.5%) | 0.008 |
| Obesity, unspecified | 64,540 (6.8%) | 73,429 (6.8%) | 0.003 | 58,387 (6.8%) | 57,306 (6.7%) | 0.005 |
| Overweight | 14,917 (1.6%) | 14,430 (1.3%) | 0.021 | 12,422 (1.4%) | 12,147 (1.4%) | 0.003 |
| Problems related to housing and economic circumstances | 4,883 (0.5%) | 7,527 (0.7%) | 0.023 | 4,596 (0.5%) | 4,367 (0.5%) | 0.004 |
| **Medications — no. (%)** |  |  |  |  |  |  |
| HMG CoA reductase inhibitors | 211,160 (22.4%) | 245,062 (22.6%) | 0.005 | 189,041 (22.0%) | 187,743 (21.9%) | 0.004 |
| Beta blocking agents | 156,092 (16.6%) | 169,802 (15.7%) | 0.024 | 136,480 (15.9%) | 135,079 (15.7%) | 0.004 |
| Calcium channel blockers | 127,895 (13.6%) | 111,215 (10.3%) | 0.102 | 101,117 (11.8%) | 99,559 (11.6%) | 0.006 |
| Platelet aggregation inhibitors | 72,068 (7.6%) | 91,830 (8.5%) | 0.031 | 65,394 (7.6%) | 63,923 (7.5%) | 0.006 |
| Direct factor Xa inhibitors | 19,874 (2.1%) | 16,614 (1.5%) | 0.043 | 15,181 (1.8%) | 14,895 (1.7%) | 0.003 |
| Vitamin K antagonists | 10,223 (1.1%) | 16,263 (1.5%) | 0.037 | 9,931 (1.2%) | 9,678 (1.1%) | 0.003 |
| **Proxy measures — no. (%)** |  |  |  |  |  |  |
| Established Patient | 466,306 (49.5%) | 542,663 (50.1%) | 0.012 | 422,941 (49.3%) | 422,010 (49.2%) | 0.002 |
| New Patient | 238,213 (25.3%) | 271,662 (25.1%) | 0.005 | 213,743 (24.9%) | 211,521 (24.7%) | 0.006 |
| Emergency Department Services | 145,257 (15.4%) | 209,626 (19.4%) | 0.104 | 138,059 (16.1%) | 135,737 (15.8%) | 0.007 |
| State Medicaid Agency | 20,412 (2.2%) | 28,203 (2.6%) | 0.029 | 18,754 (2.2%) | 17,991 (2.1%) | 0.006 |

**Table S7A. Candesartan vs. Losartan — Baseline Characteristics**

| **Characteristic** | **Before PSM** | | | **After PSM** | | |
| --- | --- | --- | --- | --- | --- | --- |
|  | **Candesartan (N=10,113)** | **Losartan (N=583,461)** | **SMD** | **Candesartan (N=10,113)** | **Losartan (N=10,113)** | **SMD** |
| Age — Mean (SD) | 63.6 (6.1) | 64.1 (6.4) | 0.071 | 63.6 (6.1) | 63.6 (6.2) | 0.004 |
| **Sex — no. (%)** |  |  |  |  |  |  |
| Female | 5,464 (54.0%) | 312,128 (53.5%) | 0.011 | 5,464 (54.0%) | 5,443 (53.8%) | 0.004 |
| Male | 4,646 (45.9%) | 270,748 (46.4%) | 0.009 | 4,646 (45.9%) | 4,667 (46.1%) | 0.004 |
| Unknown Gender | 10 (0.1%) | 585 (0.1%) | < 0.001 | 10 (0.1%) | 10 (0.1%) | < 0.001 |
| **Race — no. (%)** |  |  |  |  |  |  |
| American Indian or Alaska Native | 16 (0.2%) | 1,626 (0.3%) | 0.026 | 16 (0.2%) | 22 (0.2%) | 0.014 |
| Asian | 415 (4.1%) | 46,373 (7.9%) | 0.162 | 415 (4.1%) | 426 (4.2%) | 0.005 |
| Black or African American | 1,137 (11.2%) | 99,656 (17.1%) | 0.168 | 1,137 (11.2%) | 1,134 (11.2%) | < 0.001 |
| Native Hawaiian or Other Pacific Islander | 27 (0.3%) | 2,505 (0.4%) | 0.028 | 27 (0.3%) | 31 (0.3%) | 0.007 |
| Other Race | 819 (8.1%) | 49,155 (8.4%) | 0.012 | 819 (8.1%) | 777 (7.7%) | 0.015 |
| Unknown Race | 1,015 (10.0%) | 42,695 (7.3%) | 0.097 | 1,015 (10.0%) | 987 (9.8%) | 0.009 |
| White | 6,684 (66.1%) | 341,451 (58.5%) | 0.157 | 6,684 (66.1%) | 6,736 (66.6%) | 0.011 |
| **Ethnicity — no. (%)** |  |  |  |  |  |  |
| Hispanic or Latino | 901 (8.9%) | 58,774 (10.1%) | 0.040 | 901 (8.9%) | 814 (8.0%) | 0.031 |
| Not Hispanic or Latino | 7,839 (77.5%) | 444,917 (76.3%) | 0.030 | 7,839 (77.5%) | 7,981 (78.9%) | 0.034 |
| Unknown Ethnicity | 1,373 (13.6%) | 79,770 (13.7%) | 0.003 | 1,373 (13.6%) | 1,318 (13.0%) | 0.016 |
| **Comorbidities — no. (%)** |  |  |  |  |  |  |
| Essential (primary) hypertension | 3,791 (37.5%) | 275,406 (47.2%) | 0.198 | 3,791 (37.5%) | 3,781 (37.4%) | 0.002 |
| Disorders of lipoprotein metabolism | 3,043 (30.1%) | 213,152 (36.5%) | 0.137 | 3,043 (30.1%) | 2,994 (29.6%) | 0.011 |
| Type 2 diabetes mellitus | 1,393 (13.8%) | 122,348 (21.0%) | 0.191 | 1,393 (13.8%) | 1,380 (13.6%) | 0.004 |
| Chronic ischemic heart disease | 764 (7.6%) | 56,695 (9.7%) | 0.077 | 764 (7.6%) | 688 (6.8%) | 0.029 |
| Chronic kidney disease (CKD) | 375 (3.7%) | 32,454 (5.6%) | 0.088 | 375 (3.7%) | 333 (3.3%) | 0.023 |
| Atrial fibrillation and flutter | 309 (3.1%) | 23,354 (4.0%) | 0.051 | 309 (3.1%) | 261 (2.6%) | 0.029 |
| Heart failure | 277 (2.7%) | 26,726 (4.6%) | 0.098 | 277 (2.7%) | 236 (2.3%) | 0.026 |
| Acute myocardial infarction | 68 (0.7%) | 10,533 (1.8%) | 0.103 | 68 (0.7%) | 66 (0.7%) | 0.002 |
| Cerebral infarction | 108 (1.1%) | 11,789 (2.0%) | 0.077 | 108 (1.1%) | 111 (1.1%) | 0.003 |
| Other peripheral vascular diseases | 179 (1.8%) | 17,749 (3.0%) | 0.083 | 179 (1.8%) | 165 (1.6%) | 0.011 |
| Major depressive disorder | 258 (2.6%) | 17,275 (3.0%) | 0.025 | 258 (2.6%) | 233 (2.3%) | 0.016 |
| Other anxiety disorders | 739 (7.3%) | 49,557 (8.5%) | 0.044 | 739 (7.3%) | 741 (7.3%) | < 0.001 |
| Mental and behavioral disorders due to psychoactive substance use | 373 (3.7%) | 52,838 (9.1%) | 0.221 | 373 (3.7%) | 369 (3.6%) | 0.002 |
| Alcohol related disorders | 57 (0.6%) | 11,689 (2.0%) | 0.128 | 57 (0.6%) | 63 (0.6%) | 0.008 |
| Nicotine dependence | 276 (2.7%) | 41,669 (7.1%) | 0.205 | 276 (2.7%) | 278 (2.7%) | 0.001 |
| Obesity, unspecified | 508 (5.0%) | 43,351 (7.4%) | 0.100 | 508 (5.0%) | 502 (5.0%) | 0.003 |
| Overweight | 133 (1.3%) | 10,361 (1.8%) | 0.037 | 133 (1.3%) | 120 (1.2%) | 0.012 |
| Problems related to housing and economic circumstances | 21 (0.2%) | 3,740 (0.6%) | 0.067 | 21 (0.2%) | 26 (0.3%) | 0.010 |
| **Medications — no. (%)** |  |  |  |  |  |  |
| HMG CoA reductase inhibitors | 2,085 (20.6%) | 136,788 (23.4%) | 0.068 | 2,085 (20.6%) | 2,068 (20.4%) | 0.004 |
| BETA BLOCKING AGENTS | 1,484 (14.7%) | 99,907 (17.1%) | 0.067 | 1,484 (14.7%) | 1,489 (14.7%) | 0.001 |
| CALCIUM CHANNEL BLOCKERS | 994 (9.8%) | 85,033 (14.6%) | 0.145 | 994 (9.8%) | 1,004 (9.9%) | 0.003 |
| Platelet aggregation inhibitors | 571 (5.6%) | 47,181 (8.1%) | 0.097 | 571 (5.6%) | 551 (5.4%) | 0.009 |
| Direct factor Xa inhibitors | 146 (1.4%) | 12,597 (2.2%) | 0.054 | 146 (1.4%) | 128 (1.3%) | 0.015 |
| Vitamin K antagonists | 117 (1.2%) | 6,073 (1.0%) | 0.011 | 117 (1.2%) | 104 (1.0%) | 0.012 |
| **Proxy measures — no. (%)** |  |  |  |  |  |  |
| Established Patient | 4,702 (46.5%) | 300,145 (51.4%) | 0.099 | 4,702 (46.5%) | 4,697 (46.4%) | < 0.001 |
| New Patient | 2,259 (22.3%) | 160,801 (27.6%) | 0.121 | 2,259 (22.3%) | 2,242 (22.2%) | 0.004 |
| Emergency Department Services | 993 (9.8%) | 100,706 (17.3%) | 0.219 | 993 (9.8%) | 1,028 (10.2%) | 0.012 |
| State Medicaid Agency | 80 (0.8%) | 15,319 (2.6%) | 0.142 | 80 (0.8%) | 76 (0.8%) | 0.005 |

**Table S7B. Irbesartan vs. Losartan — Baseline Characteristics**

| **Characteristic** | **Before PSM** | | | **After PSM** | | |
| --- | --- | --- | --- | --- | --- | --- |
|  | **Irbesartan (N=41,709)** | **Losartan (N=583,461)** | **SMD** | **Irbesartan (N=41,706)** | **Losartan (N=41,706)** | **SMD** |
| Age — Mean (SD) | 64.3 (6.3) | 64.1 (6.4) | 0.039 | 64.3 (6.3) | 64.2 (6.3) | 0.016 |
| **Sex — no. (%)** |  |  |  |  |  |  |
| Female | 21,593 (51.8%) | 312,128 (53.5%) | 0.035 | 21,593 (51.8%) | 21,517 (51.6%) | 0.004 |
| Male | 19,952 (47.8%) | 270,748 (46.4%) | 0.029 | 19,952 (47.8%) | 20,029 (48.0%) | 0.004 |
| Unknown Gender | 164 (0.4%) | 585 (0.1%) | 0.059 | 161 (0.4%) | 160 (0.4%) | < 0.001 |
| **Race — no. (%)** |  |  |  |  |  |  |
| American Indian or Alaska Native | 96 (0.2%) | 1,626 (0.3%) | 0.010 | 96 (0.2%) | 98 (0.2%) | < 0.001 |
| Asian | 2,181 (5.2%) | 46,373 (7.9%) | 0.110 | 2,181 (5.2%) | 2,236 (5.4%) | 0.006 |
| Black or African American | 5,102 (12.2%) | 99,656 (17.1%) | 0.137 | 5,102 (12.2%) | 5,278 (12.7%) | 0.013 |
| Native Hawaiian or Other Pacific Islander | 103 (0.2%) | 2,505 (0.4%) | 0.031 | 103 (0.2%) | 112 (0.3%) | 0.004 |
| Other Race | 2,985 (7.2%) | 49,155 (8.4%) | 0.047 | 2,985 (7.2%) | 2,917 (7.0%) | 0.006 |
| Unknown Race | 3,682 (8.8%) | 42,695 (7.3%) | 0.055 | 3,681 (8.8%) | 3,583 (8.6%) | 0.008 |
| White | 27,560 (66.1%) | 341,451 (58.5%) | 0.156 | 27,558 (66.1%) | 27,482 (65.9%) | 0.004 |
| **Ethnicity — no. (%)** |  |  |  |  |  |  |
| Hispanic or Latino | 3,270 (7.8%) | 58,774 (10.1%) | 0.078 | 3,270 (7.8%) | 3,224 (7.7%) | 0.004 |
| Not Hispanic or Latino | 31,955 (76.6%) | 444,917 (76.3%) | 0.008 | 31,954 (76.6%) | 32,266 (77.4%) | 0.018 |
| Unknown Ethnicity | 6,484 (15.5%) | 79,770 (13.7%) | 0.053 | 6,482 (15.5%) | 6,216 (14.9%) | 0.018 |
| **Comorbidities — no. (%)** |  |  |  |  |  |  |
| Essential (primary) hypertension | 17,158 (41.1%) | 275,406 (47.2%) | 0.122 | 17,156 (41.1%) | 16,771 (40.2%) | 0.019 |
| Disorders of lipoprotein metabolism | 13,322 (31.9%) | 213,152 (36.5%) | 0.097 | 13,320 (31.9%) | 12,931 (31.0%) | 0.020 |
| Type 2 diabetes mellitus | 7,572 (18.2%) | 122,348 (21.0%) | 0.071 | 7,571 (18.2%) | 7,157 (17.2%) | 0.026 |
| Chronic ischemic heart disease | 3,354 (8.0%) | 56,695 (9.7%) | 0.059 | 3,353 (8.0%) | 2,972 (7.1%) | 0.035 |
| Chronic kidney disease (CKD) | 1,900 (4.6%) | 32,454 (5.6%) | 0.046 | 1,900 (4.6%) | 1,605 (3.8%) | 0.035 |
| Atrial fibrillation and flutter | 1,338 (3.2%) | 23,354 (4.0%) | 0.043 | 1,337 (3.2%) | 1,124 (2.7%) | 0.030 |
| Heart failure | 1,085 (2.6%) | 26,726 (4.6%) | 0.107 | 1,085 (2.6%) | 982 (2.4%) | 0.016 |
| Acute myocardial infarction | 360 (0.9%) | 10,533 (1.8%) | 0.082 | 360 (0.9%) | 381 (0.9%) | 0.005 |
| Cerebral infarction | 510 (1.2%) | 11,789 (2.0%) | 0.063 | 510 (1.2%) | 434 (1.0%) | 0.017 |
| Other peripheral vascular diseases | 885 (2.1%) | 17,749 (3.0%) | 0.058 | 885 (2.1%) | 744 (1.8%) | 0.024 |
| Major depressive disorder | 820 (2.0%) | 17,275 (3.0%) | 0.064 | 820 (2.0%) | 746 (1.8%) | 0.013 |
| Other anxiety disorders | 2,491 (6.0%) | 49,557 (8.5%) | 0.097 | 2,491 (6.0%) | 2,362 (5.7%) | 0.013 |
| Mental and behavioral disorders due to psychoactive substance use | 1,932 (4.6%) | 52,838 (9.1%) | 0.176 | 1,930 (4.6%) | 1,932 (4.6%) | < 0.001 |
| Alcohol related disorders | 388 (0.9%) | 11,689 (2.0%) | 0.089 | 387 (0.9%) | 405 (1.0%) | 0.004 |
| Nicotine dependence | 1,460 (3.5%) | 41,669 (7.1%) | 0.163 | 1,459 (3.5%) | 1,484 (3.6%) | 0.003 |
| Obesity, unspecified | 2,357 (5.7%) | 43,351 (7.4%) | 0.072 | 2,357 (5.7%) | 2,187 (5.2%) | 0.018 |
| Overweight | 540 (1.3%) | 10,361 (1.8%) | 0.039 | 540 (1.3%) | 454 (1.1%) | 0.019 |
| Problems related to housing and economic circumstances | 69 (0.2%) | 3,740 (0.6%) | 0.075 | 69 (0.2%) | 78 (0.2%) | 0.005 |
| **Medications — no. (%)** |  |  |  |  |  |  |
| HMG CoA reductase inhibitors | 7,934 (19.0%) | 136,788 (23.4%) | 0.108 | 7,933 (19.0%) | 7,873 (18.9%) | 0.004 |
| BETA BLOCKING AGENTS | 5,628 (13.5%) | 99,907 (17.1%) | 0.101 | 5,627 (13.5%) | 5,295 (12.7%) | 0.024 |
| CALCIUM CHANNEL BLOCKERS | 4,691 (11.2%) | 85,033 (14.6%) | 0.099 | 4,690 (11.2%) | 4,495 (10.8%) | 0.015 |
| Platelet aggregation inhibitors | 2,156 (5.2%) | 47,181 (8.1%) | 0.117 | 2,156 (5.2%) | 1,983 (4.8%) | 0.019 |
| Direct factor Xa inhibitors | 593 (1.4%) | 12,597 (2.2%) | 0.056 | 593 (1.4%) | 544 (1.3%) | 0.010 |
| Vitamin K antagonists | 456 (1.1%) | 6,073 (1.0%) | 0.005 | 455 (1.1%) | 374 (0.9%) | 0.020 |
| **Proxy measures — no. (%)** |  |  |  |  |  |  |
| Established Patient | 19,315 (46.3%) | 300,145 (51.4%) | 0.103 | 19,313 (46.3%) | 18,832 (45.2%) | 0.023 |
| New Patient | 8,819 (21.1%) | 160,801 (27.6%) | 0.150 | 8,818 (21.1%) | 8,582 (20.6%) | 0.014 |
| Emergency Department Services | 4,299 (10.3%) | 100,706 (17.3%) | 0.203 | 4,298 (10.3%) | 4,243 (10.2%) | 0.004 |
| State Medicaid Agency | 445 (1.1%) | 15,319 (2.6%) | 0.116 | 445 (1.1%) | 461 (1.1%) | 0.004 |

**Table S7C. Olmesartan vs. Losartan — Baseline Characteristics**

| **Characteristic** | **Before PSM** | | | **After PSM** | | |
| --- | --- | --- | --- | --- | --- | --- |
|  | **Olmesartan (N=84,958)** | **Losartan (N=583,461)** | **SMD** | **Olmesartan (N=84,958)** | **Losartan (N=84,958)** | **SMD** |
| Age — Mean (SD) | 63.5 (6.2) | 64.1 (6.4) | 0.087 | 63.5 (6.2) | 63.5 (6.2) | 0.007 |
| **Sex — no. (%)** |  |  |  |  |  |  |
| Female | 43,737 (51.5%) | 312,128 (53.5%) | 0.040 | 43,737 (51.5%) | 43,611 (51.3%) | 0.003 |
| Male | 41,157 (48.4%) | 270,748 (46.4%) | 0.041 | 41,157 (48.4%) | 41,271 (48.6%) | 0.003 |
| Unknown Gender | 64 (0.1%) | 585 (0.1%) | 0.008 | 64 (0.1%) | 76 (0.1%) | 0.005 |
| **Race — no. (%)** |  |  |  |  |  |  |
| American Indian or Alaska Native | 200 (0.2%) | 1,626 (0.3%) | 0.009 | 200 (0.2%) | 176 (0.2%) | 0.006 |
| Asian | 5,164 (6.1%) | 46,373 (7.9%) | 0.073 | 5,164 (6.1%) | 5,296 (6.2%) | 0.006 |
| Black or African American | 13,715 (16.1%) | 99,656 (17.1%) | 0.025 | 13,715 (16.1%) | 13,845 (16.3%) | 0.004 |
| Native Hawaiian or Other Pacific Islander | 167 (0.2%) | 2,505 (0.4%) | 0.042 | 167 (0.2%) | 191 (0.2%) | 0.006 |
| Other Race | 6,235 (7.3%) | 49,155 (8.4%) | 0.040 | 6,235 (7.3%) | 6,197 (7.3%) | 0.002 |
| Unknown Race | 7,640 (9.0%) | 42,695 (7.3%) | 0.061 | 7,640 (9.0%) | 7,290 (8.6%) | 0.015 |
| White | 51,837 (61.0%) | 341,451 (58.5%) | 0.051 | 51,837 (61.0%) | 51,963 (61.2%) | 0.003 |
| **Ethnicity — no. (%)** |  |  |  |  |  |  |
| Hispanic or Latino | 7,967 (9.4%) | 58,774 (10.1%) | 0.023 | 7,967 (9.4%) | 7,582 (8.9%) | 0.016 |
| Not Hispanic or Latino | 63,091 (74.3%) | 444,917 (76.3%) | 0.046 | 63,091 (74.3%) | 64,037 (75.4%) | 0.026 |
| Unknown Ethnicity | 13,900 (16.4%) | 79,770 (13.7%) | 0.075 | 13,900 (16.4%) | 13,339 (15.7%) | 0.018 |
| **Comorbidities — no. (%)** |  |  |  |  |  |  |
| Essential (primary) hypertension | 35,788 (42.1%) | 275,406 (47.2%) | 0.102 | 35,788 (42.1%) | 35,064 (41.3%) | 0.017 |
| Disorders of lipoprotein metabolism | 26,498 (31.2%) | 213,152 (36.5%) | 0.113 | 26,498 (31.2%) | 25,772 (30.3%) | 0.019 |
| Type 2 diabetes mellitus | 12,944 (15.2%) | 122,348 (21.0%) | 0.149 | 12,944 (15.2%) | 12,838 (15.1%) | 0.003 |
| Chronic ischemic heart disease | 5,461 (6.4%) | 56,695 (9.7%) | 0.121 | 5,461 (6.4%) | 5,244 (6.2%) | 0.011 |
| Chronic kidney disease (CKD) | 3,044 (3.6%) | 32,454 (5.6%) | 0.095 | 3,044 (3.6%) | 2,797 (3.3%) | 0.016 |
| Atrial fibrillation and flutter | 1,998 (2.4%) | 23,354 (4.0%) | 0.094 | 1,998 (2.4%) | 1,924 (2.3%) | 0.006 |
| Heart failure | 1,485 (1.7%) | 26,726 (4.6%) | 0.162 | 1,485 (1.7%) | 1,646 (1.9%) | 0.014 |
| Acute myocardial infarction | 418 (0.5%) | 10,533 (1.8%) | 0.123 | 418 (0.5%) | 495 (0.6%) | 0.012 |
| Cerebral infarction | 899 (1.1%) | 11,789 (2.0%) | 0.078 | 899 (1.1%) | 858 (1.0%) | 0.005 |
| Other peripheral vascular diseases | 1,686 (2.0%) | 17,749 (3.0%) | 0.068 | 1,686 (2.0%) | 1,509 (1.8%) | 0.015 |
| Major depressive disorder | 1,675 (2.0%) | 17,275 (3.0%) | 0.064 | 1,675 (2.0%) | 1,512 (1.8%) | 0.014 |
| Other anxiety disorders | 5,358 (6.3%) | 49,557 (8.5%) | 0.084 | 5,358 (6.3%) | 4,979 (5.9%) | 0.019 |
| Mental and behavioral disorders due to psychoactive substance use | 4,426 (5.2%) | 52,838 (9.1%) | 0.150 | 4,426 (5.2%) | 4,361 (5.1%) | 0.003 |
| Alcohol related disorders | 895 (1.1%) | 11,689 (2.0%) | 0.077 | 895 (1.1%) | 885 (1.0%) | 0.001 |
| Nicotine dependence | 3,360 (4.0%) | 41,669 (7.1%) | 0.140 | 3,360 (4.0%) | 3,413 (4.0%) | 0.003 |
| Obesity, unspecified | 4,759 (5.6%) | 43,351 (7.4%) | 0.074 | 4,759 (5.6%) | 4,423 (5.2%) | 0.017 |
| Overweight | 1,199 (1.4%) | 10,361 (1.8%) | 0.029 | 1,199 (1.4%) | 971 (1.1%) | 0.024 |
| Problems related to housing and economic circumstances | 195 (0.2%) | 3,740 (0.6%) | 0.063 | 195 (0.2%) | 214 (0.3%) | 0.005 |
| **Medications — no. (%)** |  |  |  |  |  |  |
| HMG CoA reductase inhibitors | 16,849 (19.8%) | 136,788 (23.4%) | 0.088 | 16,849 (19.8%) | 16,444 (19.4%) | 0.012 |
| BETA BLOCKING AGENTS | 11,048 (13.0%) | 99,907 (17.1%) | 0.115 | 11,048 (13.0%) | 10,791 (12.7%) | 0.009 |
| CALCIUM CHANNEL BLOCKERS | 9,975 (11.7%) | 85,033 (14.6%) | 0.084 | 9,975 (11.7%) | 9,380 (11.0%) | 0.022 |
| Platelet aggregation inhibitors | 4,260 (5.0%) | 47,181 (8.1%) | 0.124 | 4,260 (5.0%) | 4,138 (4.9%) | 0.007 |
| Direct factor Xa inhibitors | 1,107 (1.3%) | 12,597 (2.2%) | 0.066 | 1,107 (1.3%) | 1,005 (1.2%) | 0.011 |
| Vitamin K antagonists | 561 (0.7%) | 6,073 (1.0%) | 0.041 | 561 (0.7%) | 499 (0.6%) | 0.009 |
| **Proxy measures — no. (%)** |  |  |  |  |  |  |
| Established Patient | 39,865 (46.9%) | 300,145 (51.4%) | 0.090 | 39,865 (46.9%) | 38,925 (45.8%) | 0.022 |
| New Patient | 19,043 (22.4%) | 160,801 (27.6%) | 0.119 | 19,043 (22.4%) | 18,485 (21.8%) | 0.016 |
| Emergency Department Services | 9,235 (10.9%) | 100,706 (17.3%) | 0.185 | 9,235 (10.9%) | 9,411 (11.1%) | 0.007 |
| State Medicaid Agency | 1,201 (1.4%) | 15,319 (2.6%) | 0.086 | 1,201 (1.4%) | 1,162 (1.4%) | 0.004 |

**Table S7D. Telmisartan vs. Losartan — Baseline Characteristics**

| **Characteristic** | **Before PSM** | | | **After PSM** | | |
| --- | --- | --- | --- | --- | --- | --- |
|  | **Telmisartan (N=24,295)** | **Losartan (N=583,461)** | **SMD** | **Telmisartan (N=24,294)** | **Losartan (N=24,294)** | **SMD** |
| Age — Mean (SD) | 64.1 (6.2) | 64.1 (6.4) | 0.005 | 64.1 (6.2) | 64.0 (6.2) | 0.017 |
| **Sex — no. (%)** |  |  |  |  |  |  |
| Female | 13,074 (53.8%) | 312,128 (53.5%) | 0.006 | 13,074 (53.8%) | 13,086 (53.9%) | < 0.001 |
| Male | 11,191 (46.1%) | 270,748 (46.4%) | 0.007 | 11,191 (46.1%) | 11,174 (46.0%) | 0.001 |
| Unknown Gender | 30 (0.1%) | 585 (0.1%) | 0.007 | 29 (0.1%) | 34 (0.1%) | 0.006 |
| **Race — no. (%)** |  |  |  |  |  |  |
| American Indian or Alaska Native | 91 (0.4%) | 1,626 (0.3%) | 0.017 | 91 (0.4%) | 84 (0.3%) | 0.005 |
| Asian | 2,126 (8.8%) | 46,373 (7.9%) | 0.029 | 2,126 (8.8%) | 2,137 (8.8%) | 0.002 |
| Black or African American | 3,539 (14.6%) | 99,656 (17.1%) | 0.069 | 3,539 (14.6%) | 3,607 (14.8%) | 0.008 |
| Native Hawaiian or Other Pacific Islander | 58 (0.2%) | 2,505 (0.4%) | 0.033 | 58 (0.2%) | 65 (0.3%) | 0.006 |
| Other Race | 2,219 (9.1%) | 49,155 (8.4%) | 0.025 | 2,219 (9.1%) | 2,211 (9.1%) | 0.001 |
| Unknown Race | 2,497 (10.3%) | 42,695 (7.3%) | 0.105 | 2,496 (10.3%) | 2,435 (10.0%) | 0.008 |
| White | 13,765 (56.7%) | 341,451 (58.5%) | 0.038 | 13,765 (56.7%) | 13,755 (56.6%) | < 0.001 |
| **Ethnicity — no. (%)** |  |  |  |  |  |  |
| Hispanic or Latino | 3,049 (12.5%) | 58,774 (10.1%) | 0.078 | 3,048 (12.5%) | 2,837 (11.7%) | 0.027 |
| Not Hispanic or Latino | 17,742 (73.0%) | 444,917 (76.3%) | 0.074 | 17,742 (73.0%) | 18,102 (74.5%) | 0.034 |
| Unknown Ethnicity | 3,504 (14.4%) | 79,770 (13.7%) | 0.022 | 3,504 (14.4%) | 3,355 (13.8%) | 0.018 |
| **Comorbidities — no. (%)** |  |  |  |  |  |  |
| Essential (primary) hypertension | 10,081 (41.5%) | 275,406 (47.2%) | 0.115 | 10,081 (41.5%) | 9,817 (40.4%) | 0.022 |
| Disorders of lipoprotein metabolism | 7,669 (31.6%) | 213,152 (36.5%) | 0.105 | 7,669 (31.6%) | 7,435 (30.6%) | 0.021 |
| Type 2 diabetes mellitus | 4,168 (17.2%) | 122,348 (21.0%) | 0.097 | 4,168 (17.2%) | 3,945 (16.2%) | 0.025 |
| Chronic ischemic heart disease | 1,685 (6.9%) | 56,695 (9.7%) | 0.101 | 1,685 (6.9%) | 1,496 (6.2%) | 0.031 |
| Chronic kidney disease (CKD) | 962 (4.0%) | 32,454 (5.6%) | 0.075 | 962 (4.0%) | 828 (3.4%) | 0.029 |
| Atrial fibrillation and flutter | 558 (2.3%) | 23,354 (4.0%) | 0.098 | 558 (2.3%) | 480 (2.0%) | 0.022 |
| Heart failure | 433 (1.8%) | 26,726 (4.6%) | 0.160 | 433 (1.8%) | 460 (1.9%) | 0.008 |
| Acute myocardial infarction | 147 (0.6%) | 10,533 (1.8%) | 0.110 | 147 (0.6%) | 170 (0.7%) | 0.012 |
| Cerebral infarction | 219 (0.9%) | 11,789 (2.0%) | 0.093 | 219 (0.9%) | 191 (0.8%) | 0.013 |
| Other peripheral vascular diseases | 516 (2.1%) | 17,749 (3.0%) | 0.058 | 516 (2.1%) | 431 (1.8%) | 0.025 |
| Major depressive disorder | 492 (2.0%) | 17,275 (3.0%) | 0.060 | 492 (2.0%) | 483 (2.0%) | 0.003 |
| Other anxiety disorders | 1,351 (5.6%) | 49,557 (8.5%) | 0.115 | 1,351 (5.6%) | 1,269 (5.2%) | 0.015 |
| Mental and behavioral disorders due to psychoactive substance use | 984 (4.1%) | 52,838 (9.1%) | 0.203 | 984 (4.1%) | 990 (4.1%) | 0.001 |
| Alcohol related disorders | 201 (0.8%) | 11,689 (2.0%) | 0.100 | 201 (0.8%) | 170 (0.7%) | 0.015 |
| Nicotine dependence | 736 (3.0%) | 41,669 (7.1%) | 0.188 | 736 (3.0%) | 761 (3.1%) | 0.006 |
| Obesity, unspecified | 1,303 (5.4%) | 43,351 (7.4%) | 0.085 | 1,303 (5.4%) | 1,210 (5.0%) | 0.017 |
| Overweight | 374 (1.5%) | 10,361 (1.8%) | 0.019 | 374 (1.5%) | 317 (1.3%) | 0.020 |
| Problems related to housing and economic circumstances | 45 (0.2%) | 3,740 (0.6%) | 0.071 | 45 (0.2%) | 50 (0.2%) | 0.005 |
| **Medications — no. (%)** |  |  |  |  |  |  |
| HMG CoA reductase inhibitors | 5,025 (20.7%) | 136,788 (23.4%) | 0.067 | 5,025 (20.7%) | 4,831 (19.9%) | 0.020 |
| BETA BLOCKING AGENTS | 3,185 (13.1%) | 99,907 (17.1%) | 0.112 | 3,185 (13.1%) | 3,066 (12.6%) | 0.015 |
| CALCIUM CHANNEL BLOCKERS | 2,900 (11.9%) | 85,033 (14.6%) | 0.078 | 2,900 (11.9%) | 2,736 (11.3%) | 0.021 |
| Platelet aggregation inhibitors | 1,253 (5.2%) | 47,181 (8.1%) | 0.118 | 1,253 (5.2%) | 1,159 (4.8%) | 0.018 |
| Direct factor Xa inhibitors | 294 (1.2%) | 12,597 (2.2%) | 0.074 | 294 (1.2%) | 262 (1.1%) | 0.012 |
| Vitamin K antagonists | 163 (0.7%) | 6,073 (1.0%) | 0.040 | 163 (0.7%) | 156 (0.6%) | 0.004 |
| **Proxy measures — no. (%)** |  |  |  |  |  |  |
| Established Patient | 10,959 (45.1%) | 300,145 (51.4%) | 0.127 | 10,959 (45.1%) | 10,716 (44.1%) | 0.020 |
| New Patient | 5,432 (22.4%) | 160,801 (27.6%) | 0.120 | 5,432 (22.4%) | 5,244 (21.6%) | 0.019 |
| Emergency Department Services | 2,369 (9.8%) | 100,706 (17.3%) | 0.221 | 2,369 (9.8%) | 2,363 (9.7%) | < 0.001 |
| State Medicaid Agency | 231 (1.0%) | 15,319 (2.6%) | 0.127 | 231 (1.0%) | 235 (1.0%) | 0.002 |

**Table S7E. Valsartan vs. Losartan — Baseline Characteristics**

| **Characteristic** | **Before PSM** | | | **After PSM** | | |
| --- | --- | --- | --- | --- | --- | --- |
|  | **Valsartan (N=202,930)** | **Losartan (N=583,461)** | **SMD** | **Valsartan (N=202,904)** | **Losartan (N=202,904)** | **SMD** |
| Age — Mean (SD) | 64.2 (6.3) | 64.1 (6.4) | 0.025 | 64.2 (6.3) | 64.2 (6.3) | 0.006 |
| **Sex — no. (%)** |  |  |  |  |  |  |
| Female | 105,117 (51.8%) | 312,128 (53.5%) | 0.034 | 105,115 (51.8%) | 105,829 (52.2%) | 0.007 |
| Male | 97,406 (48.0%) | 270,748 (46.4%) | 0.032 | 97,389 (48.0%) | 96,697 (47.7%) | 0.007 |
| Unknown Gender | 407 (0.2%) | 585 (0.1%) | 0.026 | 400 (0.2%) | 378 (0.2%) | 0.002 |
| **Race — no. (%)** |  |  |  |  |  |  |
| American Indian or Alaska Native | 501 (0.2%) | 1,626 (0.3%) | 0.006 | 501 (0.2%) | 458 (0.2%) | 0.004 |
| Asian | 9,935 (4.9%) | 46,373 (7.9%) | 0.125 | 9,935 (4.9%) | 10,403 (5.1%) | 0.011 |
| Black or African American | 36,668 (18.1%) | 99,656 (17.1%) | 0.026 | 36,663 (18.1%) | 36,988 (18.2%) | 0.004 |
| Native Hawaiian or Other Pacific Islander | 418 (0.2%) | 2,505 (0.4%) | 0.040 | 418 (0.2%) | 463 (0.2%) | 0.005 |
| Other Race | 16,383 (8.1%) | 49,155 (8.4%) | 0.013 | 16,383 (8.1%) | 16,137 (8.0%) | 0.004 |
| Unknown Race | 16,322 (8.0%) | 42,695 (7.3%) | 0.027 | 16,317 (8.0%) | 16,086 (7.9%) | 0.004 |
| White | 122,703 (60.5%) | 341,451 (58.5%) | 0.040 | 122,687 (60.5%) | 122,369 (60.3%) | 0.003 |
| **Ethnicity — no. (%)** |  |  |  |  |  |  |
| Hispanic or Latino | 17,471 (8.6%) | 58,774 (10.1%) | 0.050 | 17,471 (8.6%) | 16,563 (8.2%) | 0.016 |
| Not Hispanic or Latino | 154,183 (76.0%) | 444,917 (76.3%) | 0.006 | 154,169 (76.0%) | 155,534 (76.7%) | 0.016 |
| Unknown Ethnicity | 31,276 (15.4%) | 79,770 (13.7%) | 0.049 | 31,264 (15.4%) | 30,807 (15.2%) | 0.006 |
| **Comorbidities — no. (%)** |  |  |  |  |  |  |
| Essential (primary) hypertension | 85,291 (42.0%) | 275,406 (47.2%) | 0.104 | 85,279 (42.0%) | 85,181 (42.0%) | < 0.001 |
| Disorders of lipoprotein metabolism | 64,880 (32.0%) | 213,152 (36.5%) | 0.096 | 64,872 (32.0%) | 63,697 (31.4%) | 0.012 |
| Type 2 diabetes mellitus | 37,910 (18.7%) | 122,348 (21.0%) | 0.057 | 37,906 (18.7%) | 36,961 (18.2%) | 0.012 |
| Chronic ischemic heart disease | 23,544 (11.6%) | 56,695 (9.7%) | 0.061 | 23,522 (11.6%) | 21,439 (10.6%) | 0.033 |
| Chronic kidney disease (CKD) | 10,498 (5.2%) | 32,454 (5.6%) | 0.017 | 10,495 (5.2%) | 9,622 (4.7%) | 0.020 |
| Atrial fibrillation and flutter | 10,517 (5.2%) | 23,354 (4.0%) | 0.056 | 10,502 (5.2%) | 9,318 (4.6%) | 0.027 |
| Heart failure | 16,411 (8.1%) | 26,726 (4.6%) | 0.144 | 16,385 (8.1%) | 15,335 (7.6%) | 0.019 |
| Acute myocardial infarction | 5,120 (2.5%) | 10,533 (1.8%) | 0.049 | 5,111 (2.5%) | 4,642 (2.3%) | 0.015 |
| Cerebral infarction | 3,507 (1.7%) | 11,789 (2.0%) | 0.022 | 3,505 (1.7%) | 3,110 (1.5%) | 0.015 |
| Other peripheral vascular diseases | 5,797 (2.9%) | 17,749 (3.0%) | 0.011 | 5,795 (2.9%) | 5,136 (2.5%) | 0.020 |
| Major depressive disorder | 4,033 (2.0%) | 17,275 (3.0%) | 0.063 | 4,033 (2.0%) | 3,701 (1.8%) | 0.012 |
| Other anxiety disorders | 12,896 (6.4%) | 49,557 (8.5%) | 0.082 | 12,895 (6.4%) | 12,064 (5.9%) | 0.017 |
| Mental and behavioral disorders due to psychoactive substance use | 14,261 (7.0%) | 52,838 (9.1%) | 0.075 | 14,260 (7.0%) | 12,938 (6.4%) | 0.026 |
| Alcohol related disorders | 3,222 (1.6%) | 11,689 (2.0%) | 0.031 | 3,222 (1.6%) | 2,928 (1.4%) | 0.012 |
| Nicotine dependence | 11,239 (5.5%) | 41,669 (7.1%) | 0.066 | 11,238 (5.5%) | 10,184 (5.0%) | 0.023 |
| Obesity, unspecified | 12,596 (6.2%) | 43,351 (7.4%) | 0.049 | 12,596 (6.2%) | 11,805 (5.8%) | 0.016 |
| Overweight | 2,368 (1.2%) | 10,361 (1.8%) | 0.051 | 2,368 (1.2%) | 2,101 (1.0%) | 0.013 |
| Problems related to housing and economic circumstances | 832 (0.4%) | 3,740 (0.6%) | 0.032 | 832 (0.4%) | 743 (0.4%) | 0.007 |
| **Medications — no. (%)** |  |  |  |  |  |  |
| HMG CoA reductase inhibitors | 43,040 (21.2%) | 136,788 (23.4%) | 0.054 | 43,027 (21.2%) | 41,445 (20.4%) | 0.019 |
| BETA BLOCKING AGENTS | 35,365 (17.4%) | 99,907 (17.1%) | 0.008 | 35,347 (17.4%) | 33,143 (16.3%) | 0.029 |
| CALCIUM CHANNEL BLOCKERS | 24,637 (12.1%) | 85,033 (14.6%) | 0.072 | 24,635 (12.1%) | 23,669 (11.7%) | 0.015 |
| Platelet aggregation inhibitors | 17,037 (8.4%) | 47,181 (8.1%) | 0.011 | 17,023 (8.4%) | 15,351 (7.6%) | 0.030 |
| Direct factor Xa inhibitors | 5,217 (2.6%) | 12,597 (2.2%) | 0.027 | 5,215 (2.6%) | 4,547 (2.2%) | 0.021 |
| Vitamin K antagonists | 2,884 (1.4%) | 6,073 (1.0%) | 0.034 | 2,874 (1.4%) | 2,634 (1.3%) | 0.010 |
| **Proxy measures — no. (%)** |  |  |  |  |  |  |
| Established Patient | 93,064 (45.9%) | 300,145 (51.4%) | 0.112 | 93,055 (45.9%) | 92,190 (45.4%) | 0.009 |
| New Patient | 42,914 (21.1%) | 160,801 (27.6%) | 0.150 | 42,914 (21.1%) | 41,723 (20.6%) | 0.014 |
| Emergency Department Services | 28,472 (14.0%) | 100,706 (17.3%) | 0.089 | 28,467 (14.0%) | 26,939 (13.3%) | 0.022 |
| State Medicaid Agency | 3,195 (1.6%) | 15,319 (2.6%) | 0.073 | 3,195 (1.6%) | 2,909 (1.4%) | 0.012 |

**Table S8. Cohort sizes before and after matching using PSM for between-class comparisons**

| **Comparison** | **No. patients (pre matching)** | **No. patients (post matching)** | **ARB cohort retention rate (%)** |
| --- | --- | --- | --- |
| **Candesartan vs. Lisinopril** |  |  |  |
| Candesartan | 10,113 | 10,113 | 100.0 |
| Lisinopril | 1,083,068 | 10,113 |  |
| **Irbesartan vs. Lisinopril** |  |  |  |
| Irbesartan | 41,709 | 41,709 | 100.0 |
| Lisinopril | 1,083,077 | 41,709 |  |
| **Losartan vs. Lisinopril** |  |  |  |
| Losartan | 583,461 | 564,984 | 96.8 |
| Lisinopril | 1,083,068 | 564,984 |  |
| **Olmesartan vs. Lisinopril** |  |  |  |
| Olmesartan | 84,958 | 84,957 | >99.9 |
| Lisinopril | 1,083,068 | 84,957 |  |
| **Telmisartan vs. Lisinopril** |  |  |  |
| Telmisartan | 24,295 | 24,294 | >99.9 |
| Lisinopril | 1,083,068 | 24,294 |  |
| **Valsartan vs. Lisinopril** |  |  |  |
| Valsartan | 202,930 | 202,917 | >99.9 |
| Lisinopril | 1,083,068 | 202,917 |  |
| **Pooled ARB vs. Lisinopril** |  |  |  |
| Pooled ARB | 942,275 | 857,773 | 91 |
| Lisinopril | 1,083,068 | 857,773 |  |

Matching was performed separately for each pairwise comparison using predefined baseline covariates. Post-matching cohort sizes reflect the number of matched patients included in the analysis. The retention rate represents the proportion of the ARB cohort retained after matching relative to its pre-matching size. Abbreviations: PSM, propensity-score matching.

**Table S9.** **Cohort sizes before and after matching using PSM for within-class comparisons**

| **Comparison** | **No. Patients (Pre Matching)** | **No. Patients (Post Matching)** | **Other ARB Cohort Retention Rate (%)** |
| --- | --- | --- | --- |
| **Candesartan vs. Losartan** |  |  |  |
| Candesartan | 10,113 | 10,113 | 100 |
| Losartan | 583,461 | 10,113 |  |
| **Irbesartan vs. Losartan** |  |  |  |
| Irbesartan | 41,709 | 41,706 | >99.9 |
| Losartan | 583,461 | 41,706 |  |
| **Olmesartan vs. Losartan** |  |  |  |
| Olmesartan | 84,958 | 84,958 | 100 |
| Losartan | 583,461 | 84,958 |  |
| **Telmisartan vs. Losartan** |  |  |  |
| Telmisartan | 24,295 | 24,294 | >99.9 |
| Losartan | 583,461 | 24,294 |  |
| **Valsartan vs. Losartan** |  |  |  |
| Valsartan | 202,930 | 202,904 | >99.9 |
| Losartan | 583,461 | 202,904 |  |

Matching was performed separately for each pairwise comparison using predefined baseline covariates. Post-matching cohort sizes reflect the number of matched patients included in the analysis. The retention rate represents the proportion of the non-losartan ARB cohort retained after matching relative to its pre-matching size. Abbreviations: PSM, propensity-score matching.

**Table S10.** **Median follow-up and interquartile range by treatment group for between-class comparisons**

| **Comparison** | **Median (Days)** | **Median (Years)** | **IQR (Days)** | **IQR (Years)** |
| --- | --- | --- | --- | --- |
| **Candesartan vs. Lisinopril** |  |  |  |  |
| Candesartan | 2330 | 6.38 | 2822 | 7.73 |
| Lisinopril | 2422 | 6.63 | 2420 | 6.63 |
| **Irbesartan vs. Lisinopril** |  |  |  |  |
| Irbesartan | 2415 | 6.61 | 2603 | 7.13 |
| Lisinopril | 2429 | 6.65 | 2403 | 6.58 |
| **Losartan vs. Lisinopril** |  |  |  |  |
| Losartan | 1727 | 4.73 | 2250 | 6.16 |
| Lisinopril | 2343 | 6.41 | 2356 | 6.45 |
| **Olmesartan vs. Lisinopril** |  |  |  |  |
| Olmesartan | 2114 | 5.79 | 2841 | 7.78 |
| Lisinopril | 2434 | 6.66 | 2387 | 6.54 |
| **Telmisartan vs. Lisinopril** |  |  |  |  |
| Telmisartan | 2100 | 5.75 | 2963 | 8.11 |
| Lisinopril | 2395 | 6.56 | 2410 | 6.6 |
| **Valsartan vs. Lisinopril** |  |  |  |  |
| Valsartan | 2611 | 7.15 | 2945 | 8.06 |
| Lisinopril | 2397 | 6.56 | 2407 | 6.59 |
| **Pooled ARB vs. Lisinopril** |  |  |  |  |
| Pooled ARB | 1967 | 5.39 | 2521 | 6.9 |
| Lisinopril | 2380 | 6.52 | 2371 | 6.49 |

Median follow-up time and interquartile range by treatment group for between-class comparisons. Follow-up time reflects the observed record length for each patient, defined as the interval from the index date to the patient's last recorded fact date (or the end of the analysis time window, whichever came first), per the TriNetX Follow-up Time metric. This metric characterizes the duration of available observation for each cohort and is independent of outcome or death dates; accordingly, it is not censored at incident dementia or death and may exceed the time to those events. The maximum possible value is 7,300 days (20 years), as patients with index dates more than 20 years prior are excluded. Values are medians with interquartile ranges (IQRs), assessed in the matched cohorts for each comparison. Abbreviations: IQR, interquartile range.

**Table S11. Median follow-up and interquartile range by treatment group for within-class comparisons**

| **Comparison** | **Median (Days)** | **Median (Years)** | **IQR (Days)** | **IQR (Years)** |
| --- | --- | --- | --- | --- |
| **Candesartan vs. Losartan** |  |  |  |  |
| Candesartan | 2330 | 6.38 | 2822 | 7.73 |
| Losartan | 1868 | 5.11 | 2306 | 6.31 |
| **Irbesartan vs. Losartan** |  |  |  |  |
| Irbesartan | 2415 | 6.61 | 2603 | 7.13 |
| Losartan | 1832 | 5.02 | 2326 | 6.37 |
| **Olmesartan vs. Losartan** |  |  |  |  |
| Olmesartan | 2114 | 5.79 | 2841 | 7.78 |
| Losartan | 1844 | 5.05 | 2306 | 6.31 |
| **Telmisartan vs. Losartan** |  |  |  |  |
| Telmisartan | 2100 | 5.75 | 2963 | 8.11 |
| Losartan | 1839 | 5.03 | 2325 | 6.37 |
| **Valsartan vs. Losartan** |  |  |  |  |
| Valsartan | 2611 | 7.15 | 2945 | 8.06 |
| Losartan | 1785 | 4.89 | 2312 | 6.33 |

Median follow-up time and interquartile range by treatment group for within-class comparisons. Follow-up time reflects the observed record length for each patient, defined as the interval from the index date to the patient's last recorded fact date (or the end of the analysis time window, whichever came first), per the TriNetX Follow-up Time metric. This metric characterizes the duration of available observation for each cohort and is independent of outcome or death dates; accordingly, it is not censored at incident dementia or death and may exceed the time to those events. The maximum possible value is 7,300 days (20 years), as patients with index dates more than 20 years prior are excluded. Values are medians with interquartile ranges (IQRs), assessed in the matched cohorts for each comparison. Abbreviations: IQR, interquartile range.

| **Comparison** | **HR** | **ARR (%)** | **NNT** |
| --- | --- | --- | --- |
| **All-cause dementia** |  |  |  |
| Candesartan vs Lisinopril | 0.63 | 3.067 | 33 |
| Irbesartan vs Lisinopril | 0.69 | 2.887 | 35 |
| Olmesartan vs Lisinopril | 0.74 | 2.139 | 47 |
| Telmisartan vs Lisinopril | 0.77 | 1.89 | 53 |
| Valsartan vs Lisinopril | 0.83 | 1.591 | 63 |
| Losartan vs Lisinopril | 0.92 | 0.723 | 138 |
| Pooled ARBs vs Lisinopril | 0.85 | 1.357 | 74 |
| **All-cause mortality** |  |  |  |
| Candesartan vs Lisinopril | 0.61 | 7.074 | 14 |
| Irbesartan vs Lisinopril | 0.69 | 6.341 | 16 |
| Telmisartan vs Lisinopril | 0.65 | 6.356 | 16 |
| Olmesartan vs Lisinopril | 0.69 | 5.828 | 17 |
| Valsartan vs Lisinopril | 0.78 | 4.931 | 20 |
| Losartan vs Lisinopril | 0.92 | 1.843 | 54 |
| Pooled ARBs vs Lisinopril | 0.83 | 3.692 | 27 |

**Table S12. Absolute risk reduction (ARR) and number needed to treat (NNT) at 10 years for all-cause dementia and death for between-class comparisons**

Absolute risk reduction (ARR) and number needed to treat (NNT) were estimated at 10 years using Kaplan–Meier cumulative incidence estimates from the matched cohorts. ARR was calculated as the difference in absolute risk between treatment groups, and NNT as the reciprocal of ARR. ARR values are expressed as percentages, and NNT values were rounded to the nearest integer. Hazard ratios (HRs) correspond to analyses restricted to the same 10-year follow-up horizon, rounded to the 2^nd^ decimal place, and therefore may differ from the primary full-follow-up estimates. Abbreviations: ARR, absolute risk reduction; NNT, number needed to treat; HR, hazard ratio.

**Figure S2.** **Kaplan–Meier curves for dementia-free survival in between-class comparisons.**

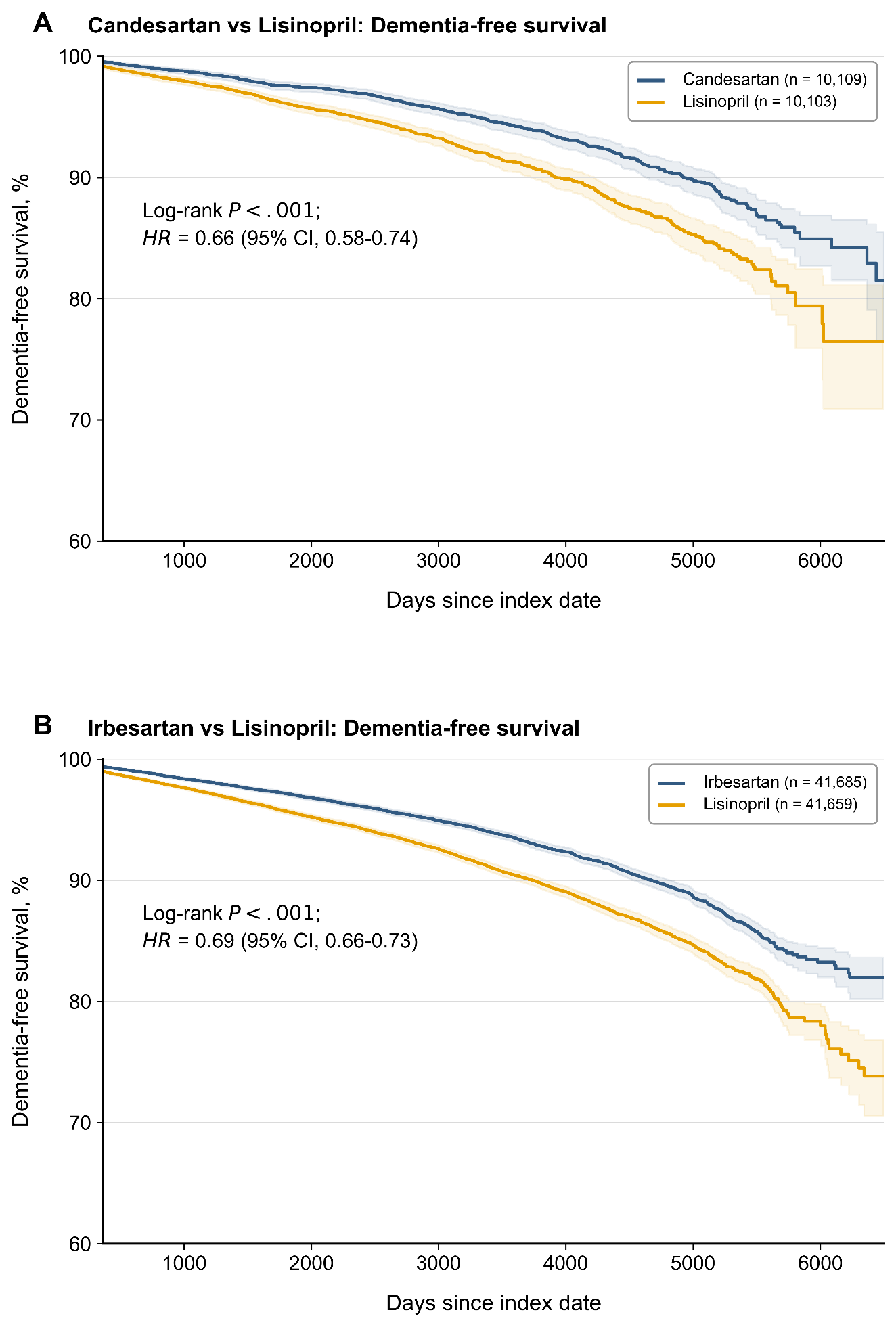

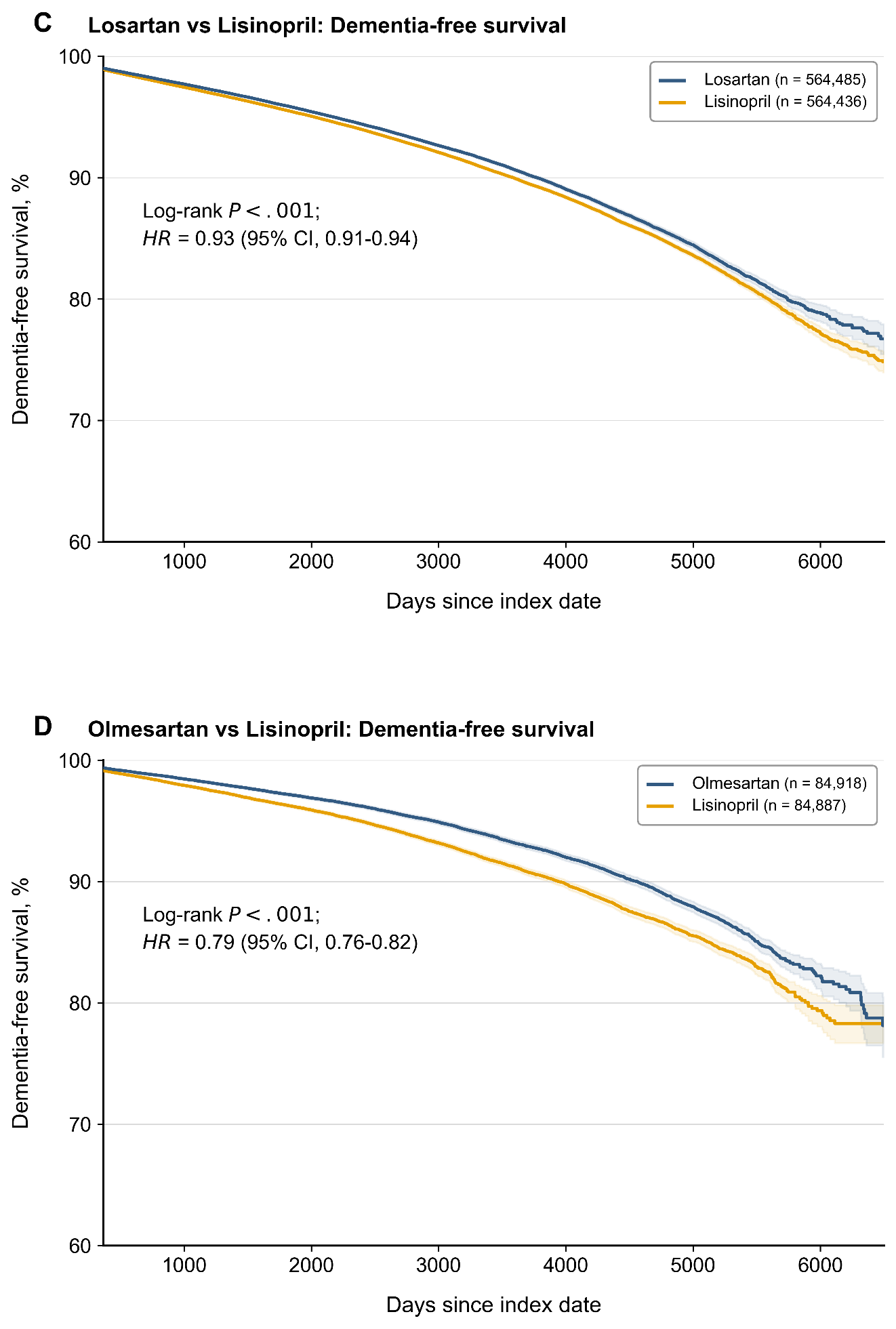

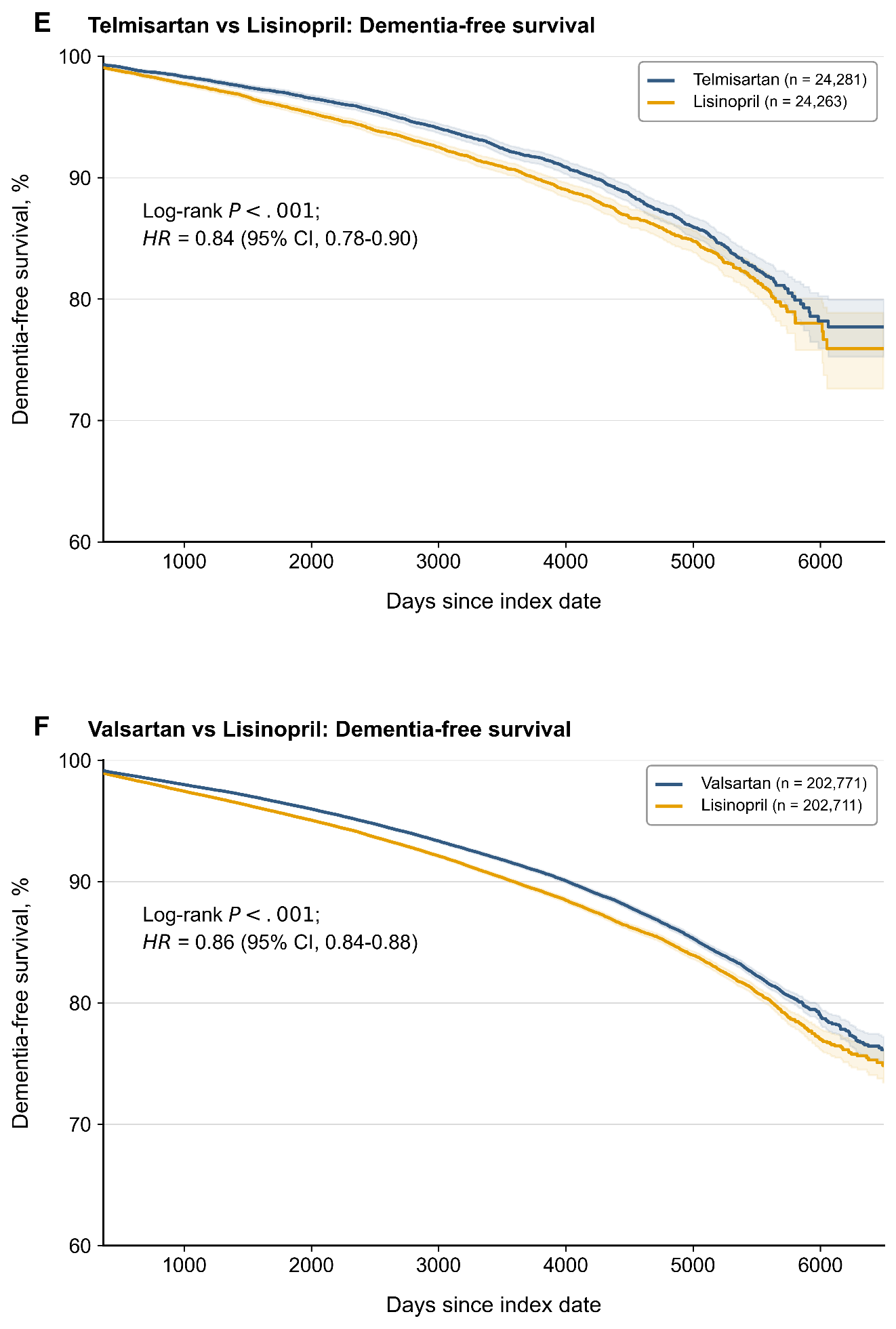

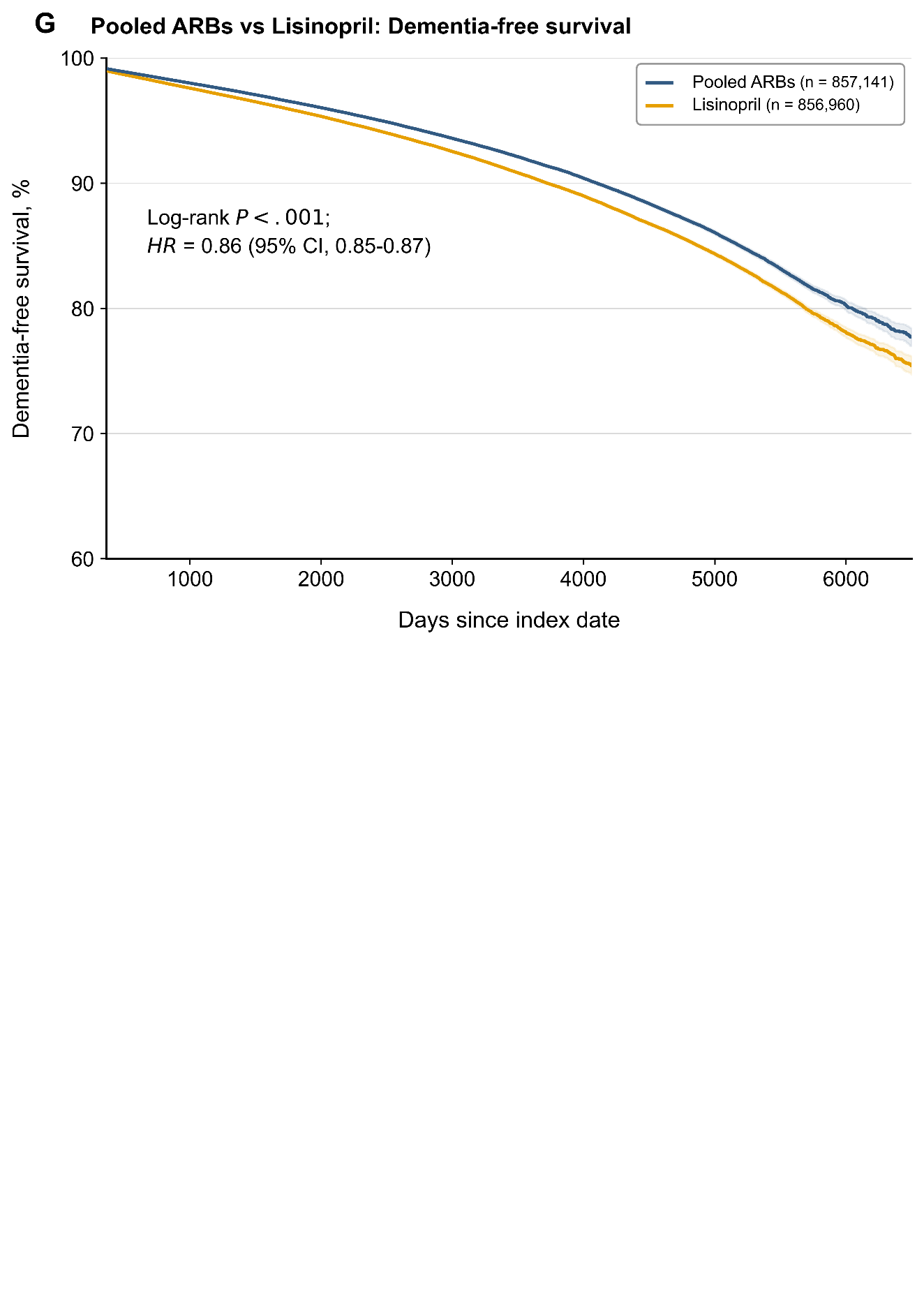

Kaplan–Meier curves show dementia-free survival for propensity-score–matched comparisons of individual and pooled angiotensin receptor blockers (ARBs) with lisinopril (reference). Patients were followed from 1 day after the index date until incident dementia; those who died, were lost to follow-up, or reached the end of the observation window were censored. Hazard ratios (HRs) with 95% confidence intervals and log-rank P values were computed over the entire follow-up period. For visual clarity, curves are truncated during the sparse terminal follow-up period; this visual truncation did not affect the hazard-ratio estimates. Shaded areas indicate 95% confidence intervals, which widen at later time points as observations become sparse. Numbers at risk at individual follow-up time points are not shown because outcome-specific risk-set counts were not available from the aggregate TriNetX analytic output used for this study. Abbreviation: HR, hazard ratio. Python/Matplotlib code used to render this figure was generated using ChatGPT (OpenAI; GPT-5-series, 2026), Claude (Anthropic; Claude Opus 4.8 and earlier models, 2026), and/or Gemini (Google; 2026); the figure was verified against the source data.

**Figure S3. Exploratory correlations of agent-level hazard ratios across outcomes in between-class comparisons**

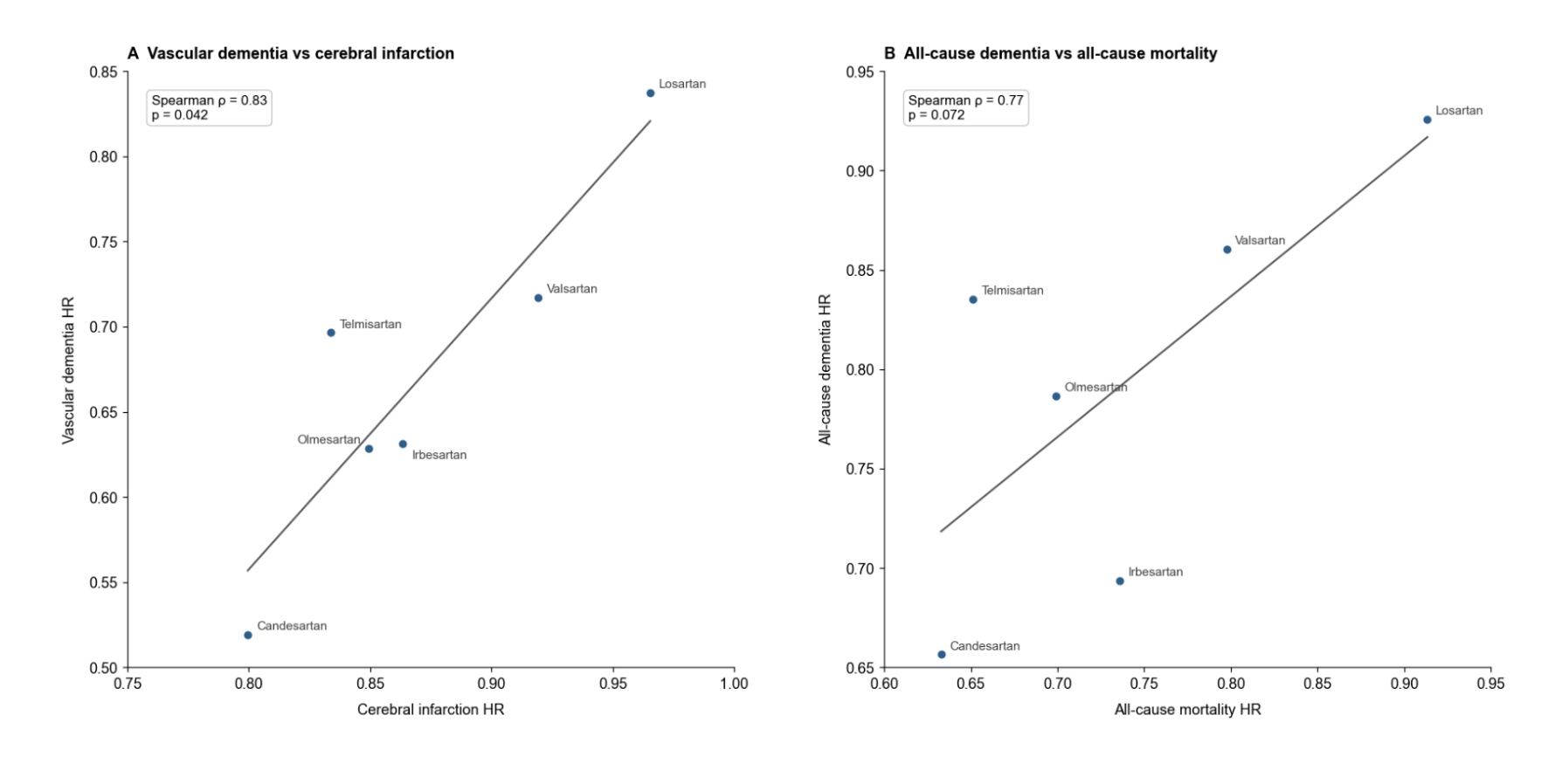

Panel A compares agent-level hazard ratios for vascular dementia and cerebral infarction. Panel B compares agent-level hazard ratios for all-cause dementia and all-cause mortality. Each point represents one individual angiotensin receptor blocker (ARB) from between-class comparisons vs lisinopril. Lines indicate fitted linear trends for visualization only. Given the small number of agents (n = 6), these analyses were descriptive and intended to assess concordance in the direction and magnitude of agent-level associations rather than formal statistical inference. Agent-level estimates for vascular dementia and cerebral infarction were positively correlated (Spearman ρ = 0.83; P = .04), and agent-level estimates for all-cause dementia and mortality showed a positive trend (Spearman ρ = 0.77; P = .07). Abbreviations: HR, hazard ratio. Python/Matplotlib code used to render this figure was generated using ChatGPT (OpenAI; GPT-5-series, 2026), Claude (Anthropic; Claude Opus 4.8 and earlier models, 2026), and/or Gemini (Google; 2026); the figure was verified against the source data.

**Figure S4.** **Kaplan–Meier curves for mortality-free survival in between-class comparisons.**

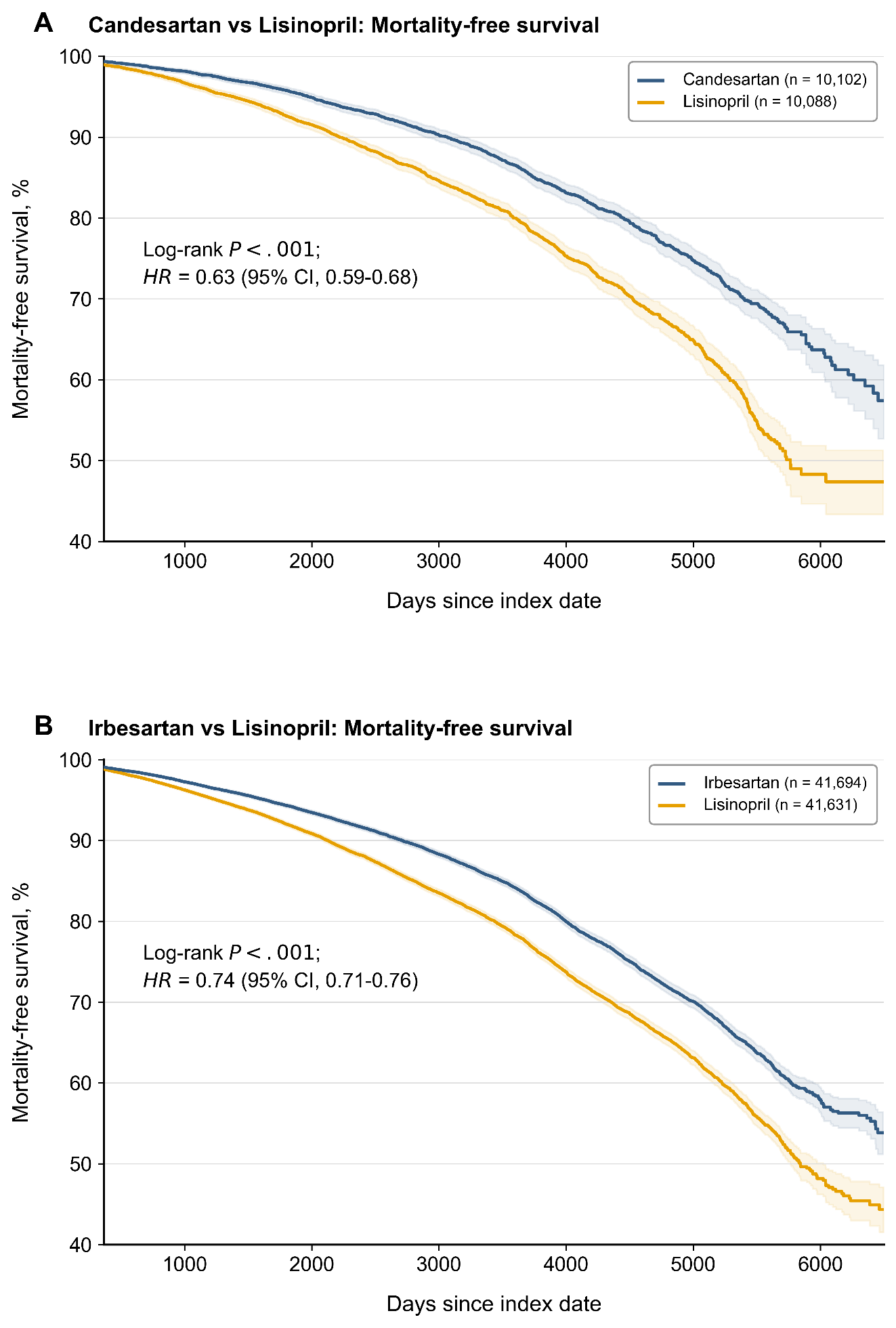

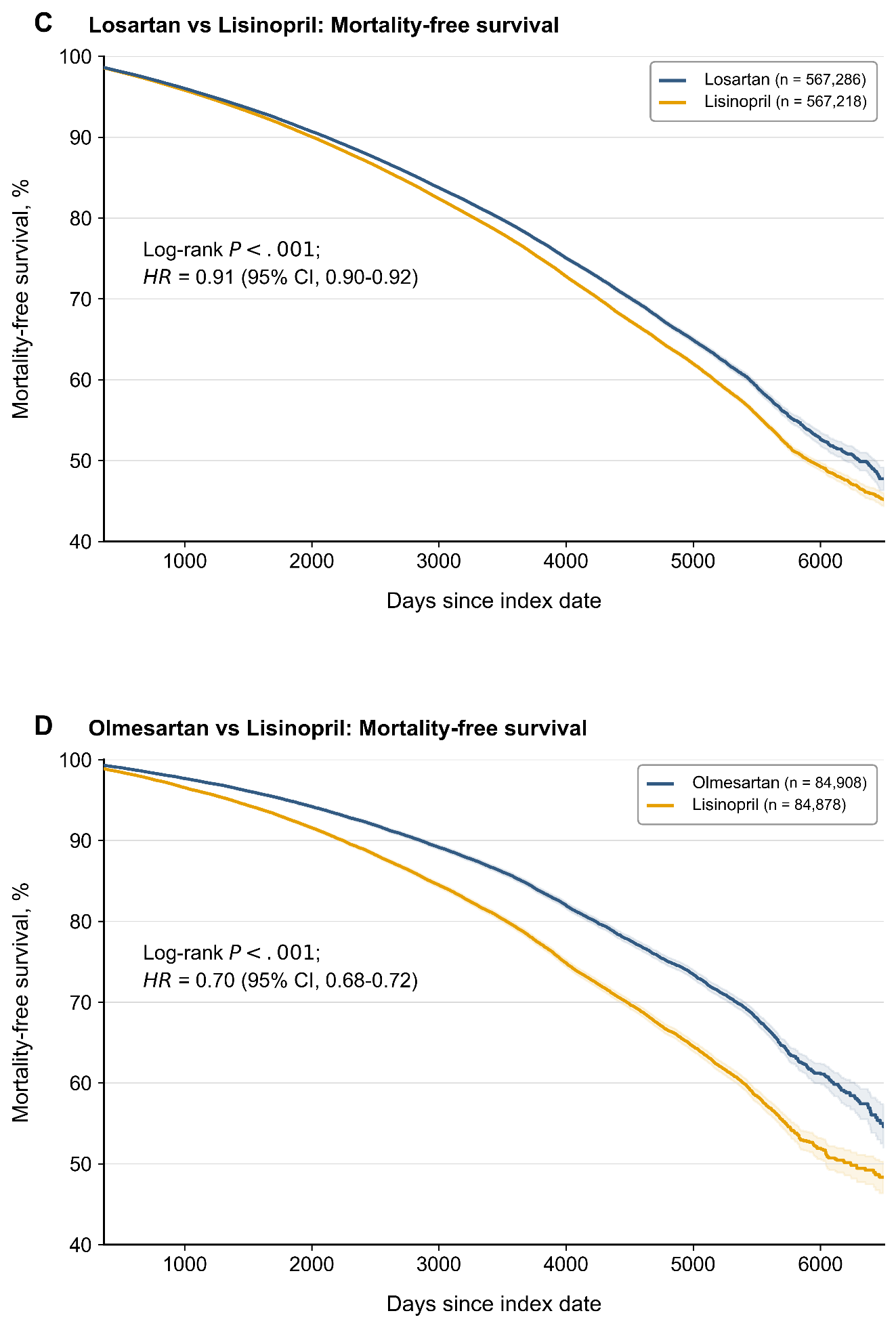

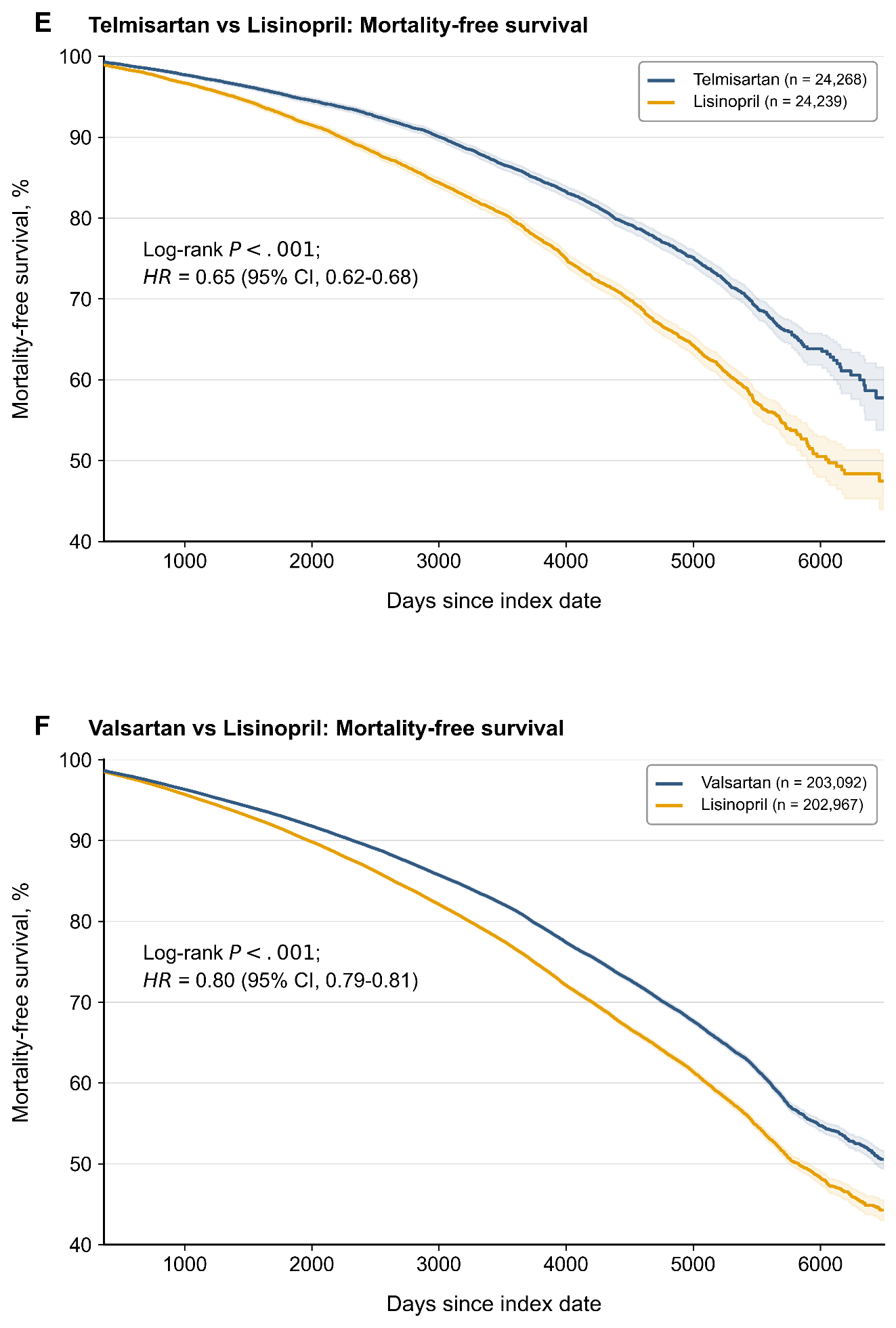

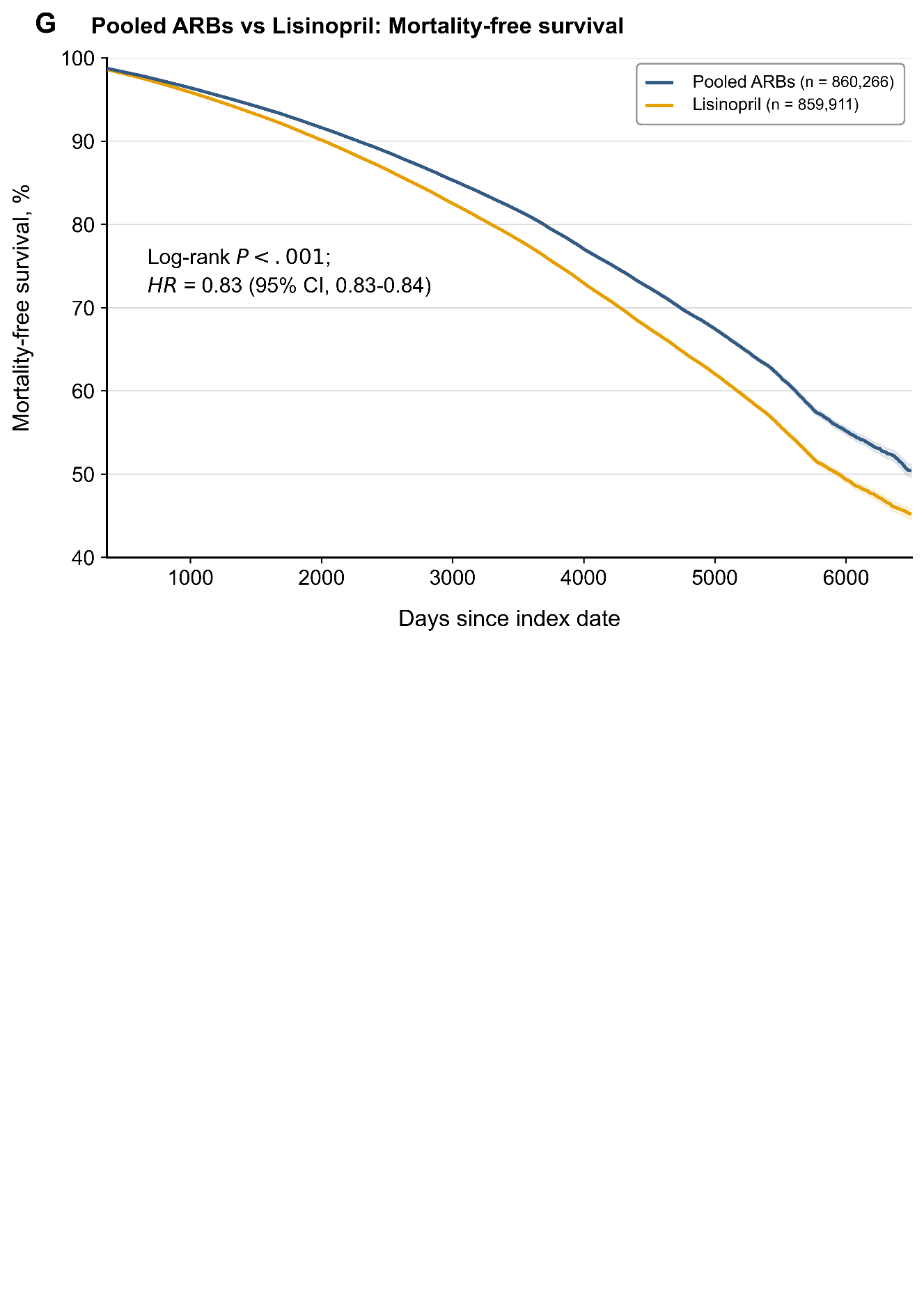

Kaplan–Meier curves show mortality-free survival for propensity-score–matched comparisons of individual and pooled angiotensin receptor blockers (ARBs) with lisinopril (reference). Patients were followed from 1 day after the index date until death, loss to follow-up, or the end of the observation window. Hazard ratios (HRs) with 95% confidence intervals and log-rank P values were computed over the entire follow-up period. For visual clarity, curves are truncated during the sparse terminal follow-up period; this visual truncation did not affect the hazard-ratio estimates. Shaded areas indicate 95% confidence intervals, which widen at later time points as observations become sparse. Numbers at risk at individual follow-up time points are not shown because outcome-specific risk-set counts were not available from the aggregate TriNetX analytic output used for this study. Abbreviation: HR, hazard ratio. Python/Matplotlib code used to render this figure was generated using ChatGPT (OpenAI; GPT-5-series, 2026), Claude (Anthropic; Claude Opus 4.8 and earlier models, 2026), and/or Gemini (Google; 2026); the figure was verified against the source data.

**Figure S5. Hazard ratios for all-cause dementia, mortality and composite outcome in between-class comparisons**

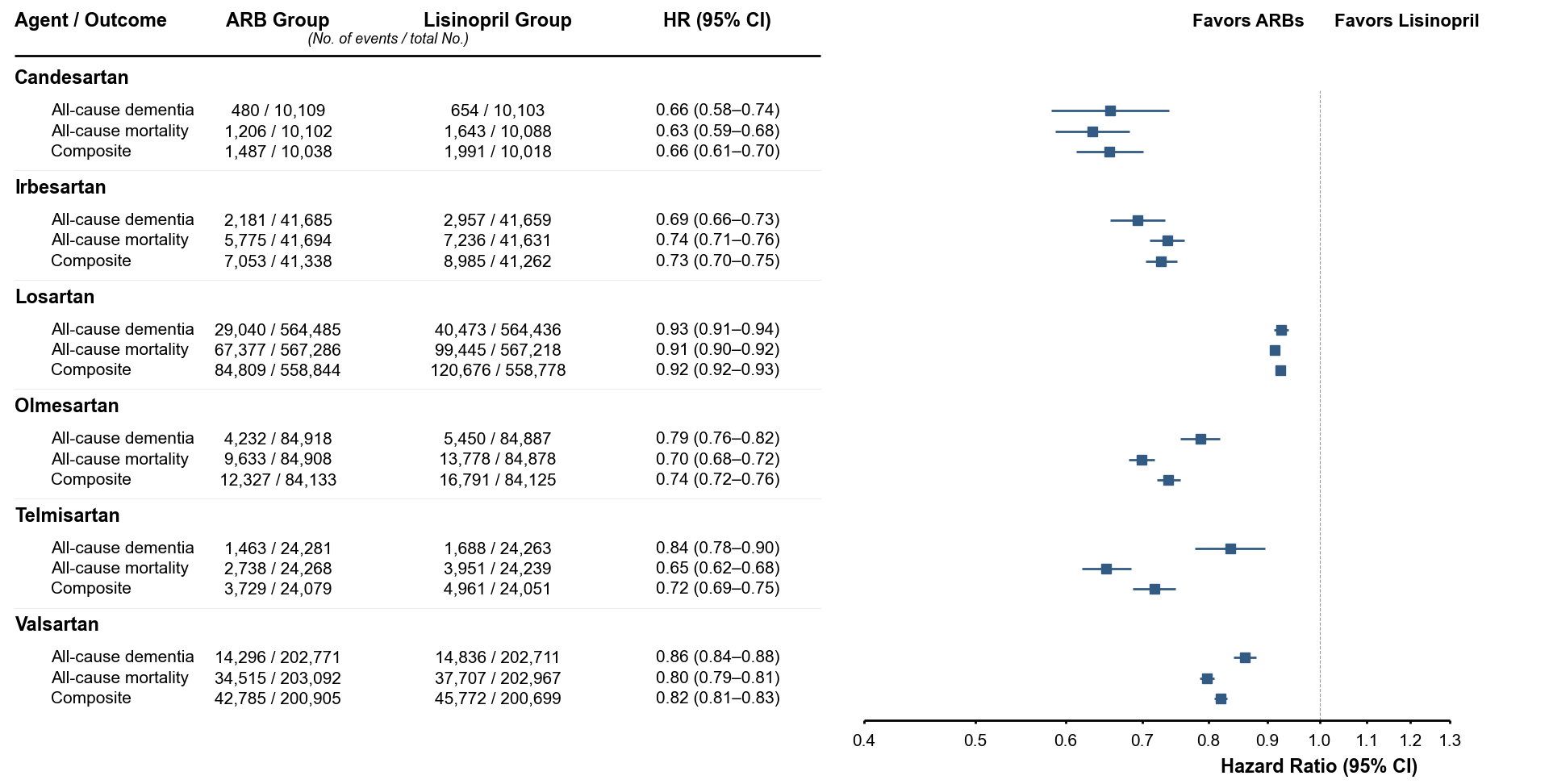

Forest plots show hazard ratios (HRs) and 95% confidence intervals for all-cause dementia (primary), all-cause mortality, and the composite outcome of dementia or death comparing individual angiotensin receptor blockers (ARBs) with lisinopril (reference). Hazard ratios are plotted on a logarithmic scale. Outcomes were defined using International Classification of Diseases, Tenth Revision (ICD-10) codes, except for death, which was identified using mortality records in the TriNetX network. Because secondary outcomes were not adjusted for multiple comparisons, findings for mortality and the composite outcome should be considered exploratory. Python/Matplotlib code used to render this figure was generated using ChatGPT (OpenAI; GPT-5-series, 2026), Claude (Anthropic; Claude Opus 4.8 and earlier models, 2026), and/or Gemini (Google; 2026); the figure was verified against the source data.

**Figure S6. Hazard ratios for secondary outcomes in within-class comparisons**

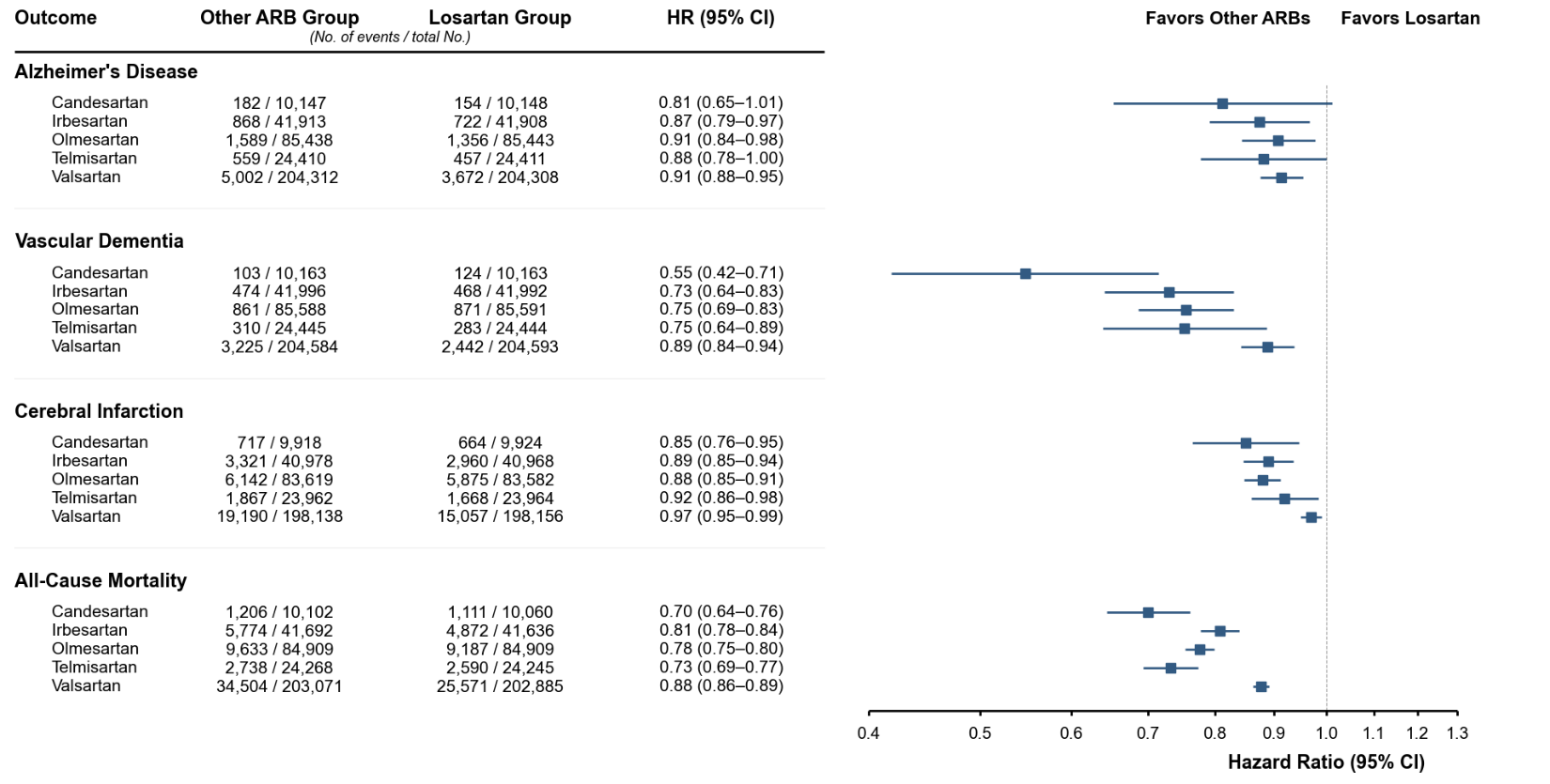

Forest plots show hazard ratios and 95% confidence intervals for selected secondary outcomes comparing other ARBs with losartan (reference), including Alzheimer disease, vascular dementia, cerebral infarction, and death. The composite outcome of dementia or death, also a secondary outcome, is presented separately in eFigure 6. Hazard ratios are plotted on a logarithmic scale. Outcomes were defined using International Classification of Diseases, 10th Revision (ICD-10) diagnostic codes, except for death, which was identified using mortality records in the TriNetX network. Because analyses of secondary outcomes were not adjusted for multiple comparisons, the hazard-ratio estimates should be considered hypothesis-generating. Python/Matplotlib code used to render this figure was generated using ChatGPT (OpenAI; GPT-5-series, 2026), Claude (Anthropic; Claude Opus 4.8 and earlier models, 2026), and/or Gemini (Google; 2026); the figure was verified against the source data.

**Figure S7. Hazard ratios for all-cause dementia, mortality and composite outcome in within-class comparisons**

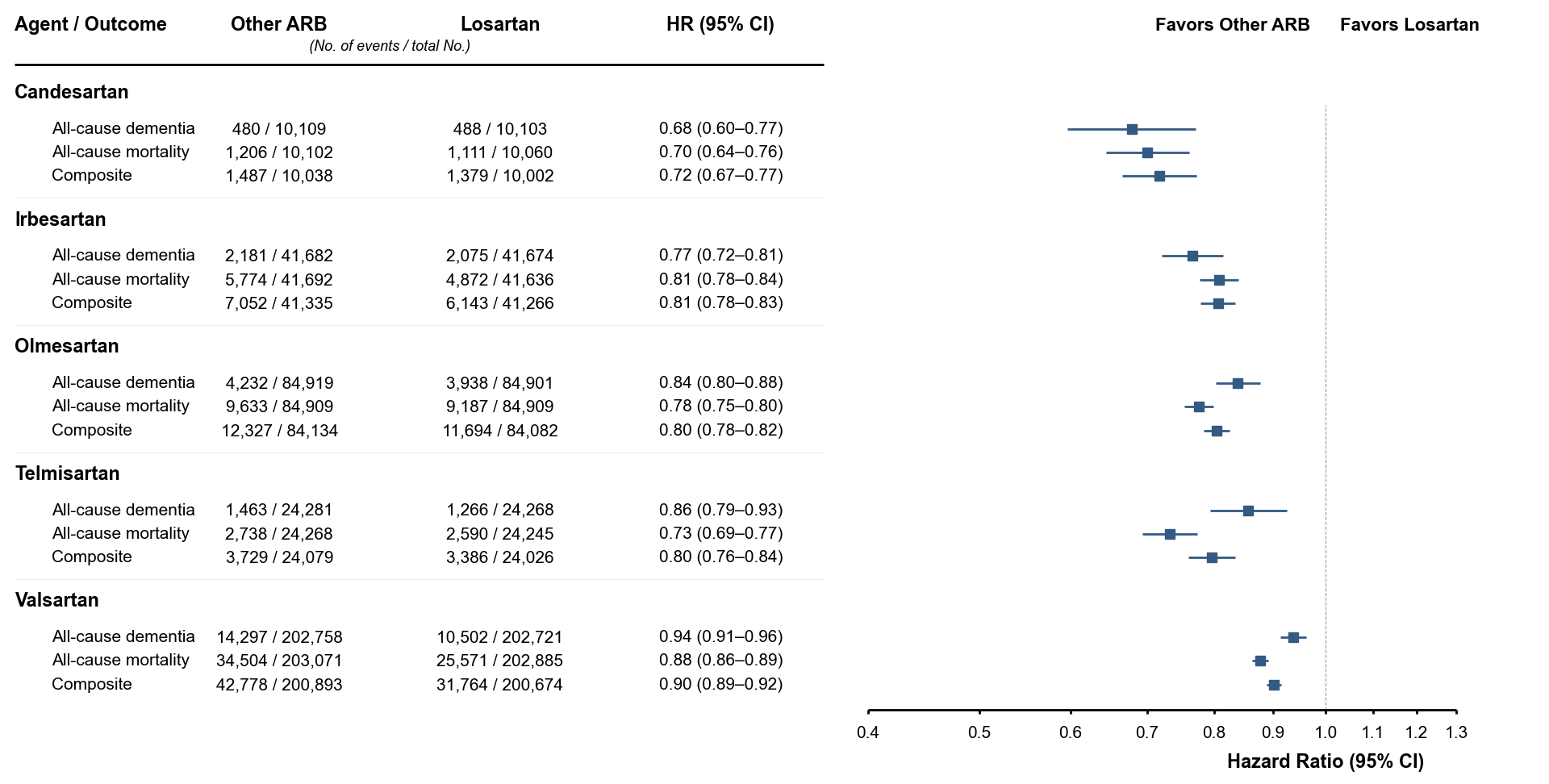

Forest plots show hazard ratios (HRs) and 95% confidence intervals for all-cause dementia (primary), all-cause mortality, and the composite outcome of dementia or death comparing other ARBs with losartan (reference). Hazard ratios are plotted on a logarithmic scale. Outcomes were defined using International Classification of Diseases, Tenth Revision (ICD-10) codes, except for death, which was identified using mortality records in the TriNetX network. Because analyses of secondary outcomes were not adjusted for multiple comparisons, these findings should be considered exploratory. Python/Matplotlib code used to render this figure was generated using ChatGPT (OpenAI; GPT-5-series, 2026), Claude (Anthropic; Claude Opus 4.8 and earlier models, 2026), and/or Gemini (Google; 2026); the figure was verified against the source data.

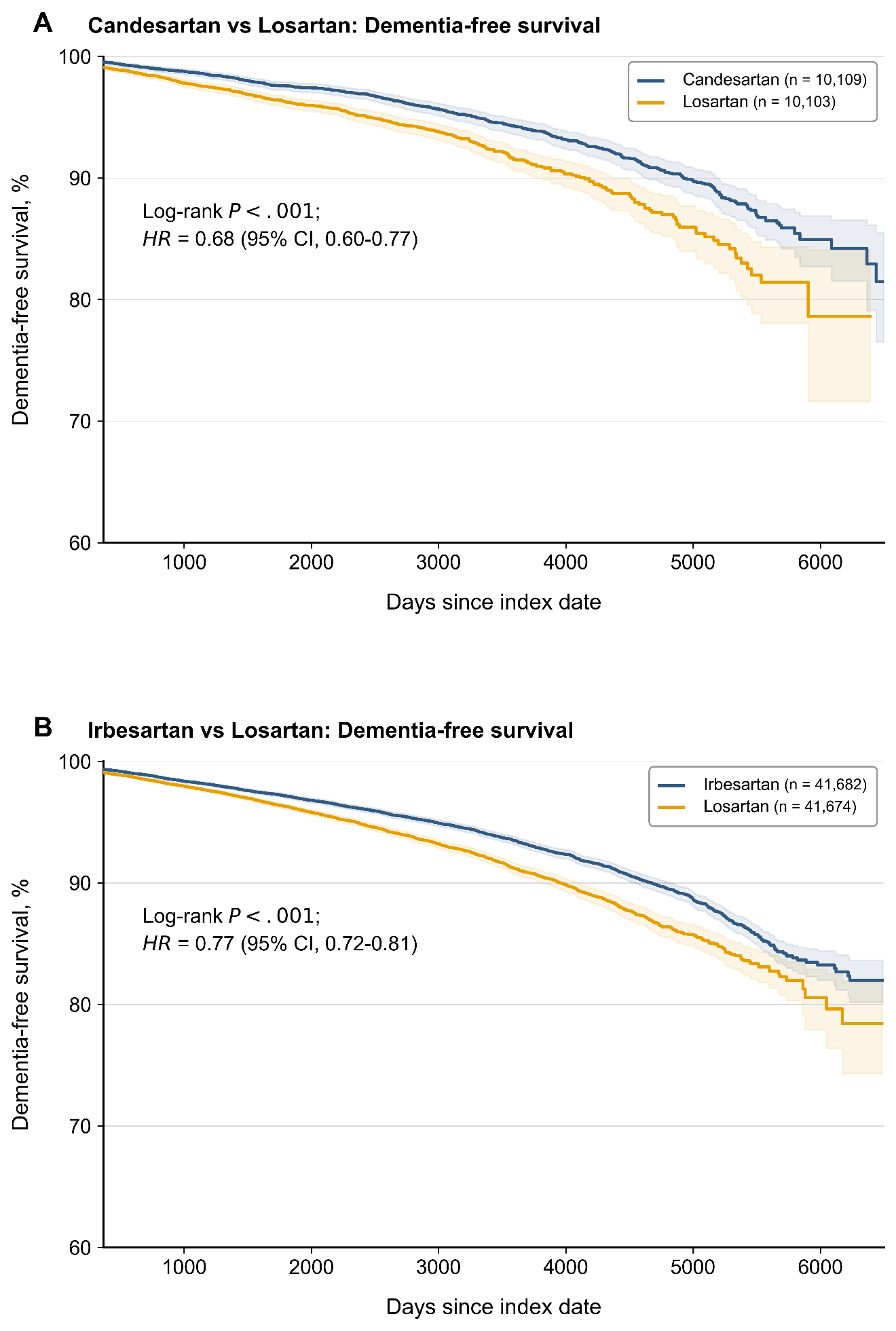
**Figure S8. Kaplan–Meier curves for dementia-free survival in within-class comparisons**

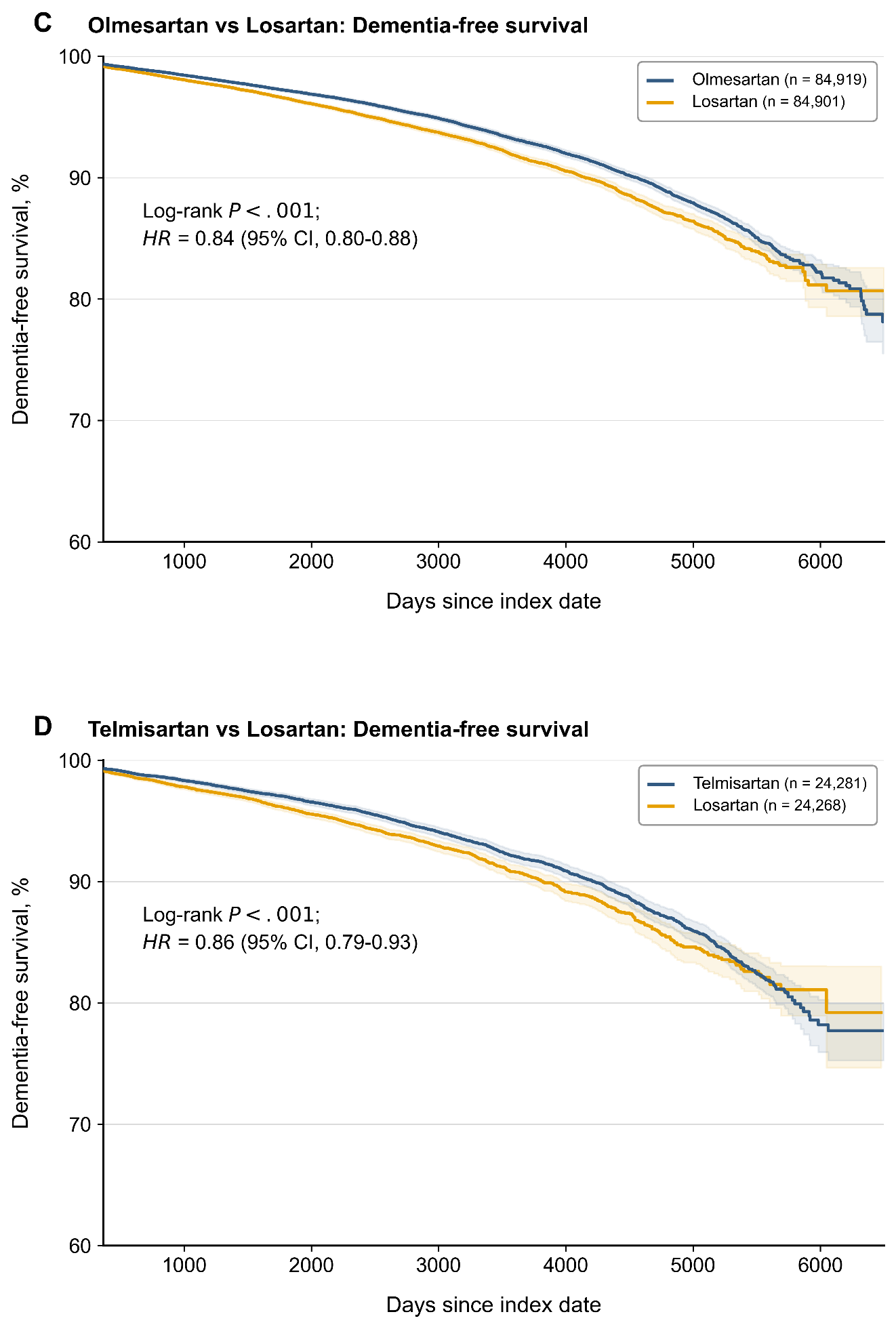

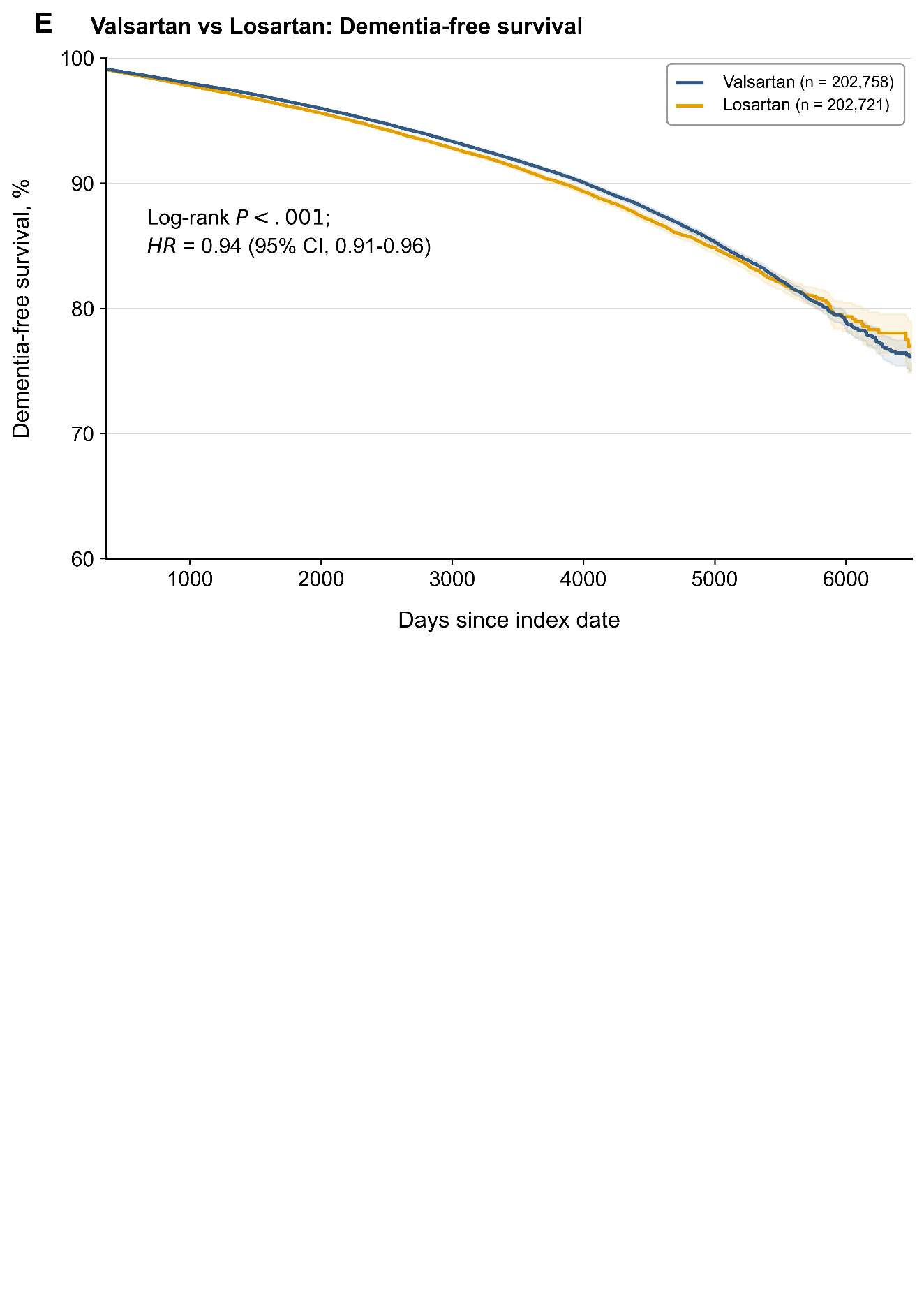

Kaplan–Meier curves show dementia-free survival for propensity-score–matched comparisons of other angiotensin receptor blockers (ARBs) with losartan (reference). Patients were followed from 1 day after the index date until incident dementia; those who died, were lost to follow-up, or reached the end of the observation window were censored. Hazard ratios (HRs) with 95% confidence intervals and log-rank P values were computed over the entire follow-up period. For visual clarity, curves are truncated during the sparse terminal follow-up period; this visual truncation did not affect the hazard-ratio estimates. Shaded areas indicate 95% confidence intervals, which widen at later time points as observations become sparse. Numbers at risk at individual follow-up time points are not shown because outcome-specific risk-set counts were not available from the aggregate TriNetX analytic output used for this study. Abbreviation: HR, hazard ratio. Python/Matplotlib code used to render this figure was generated using ChatGPT (OpenAI; GPT-5-series, 2026), Claude (Anthropic; Claude Opus 4.8 and earlier models, 2026), and/or Gemini (Google; 2026); the figure was verified against the source data.

**Figure S9.** **Kaplan–Meier Curves for** **mortality-free survival in within-class comparisons**

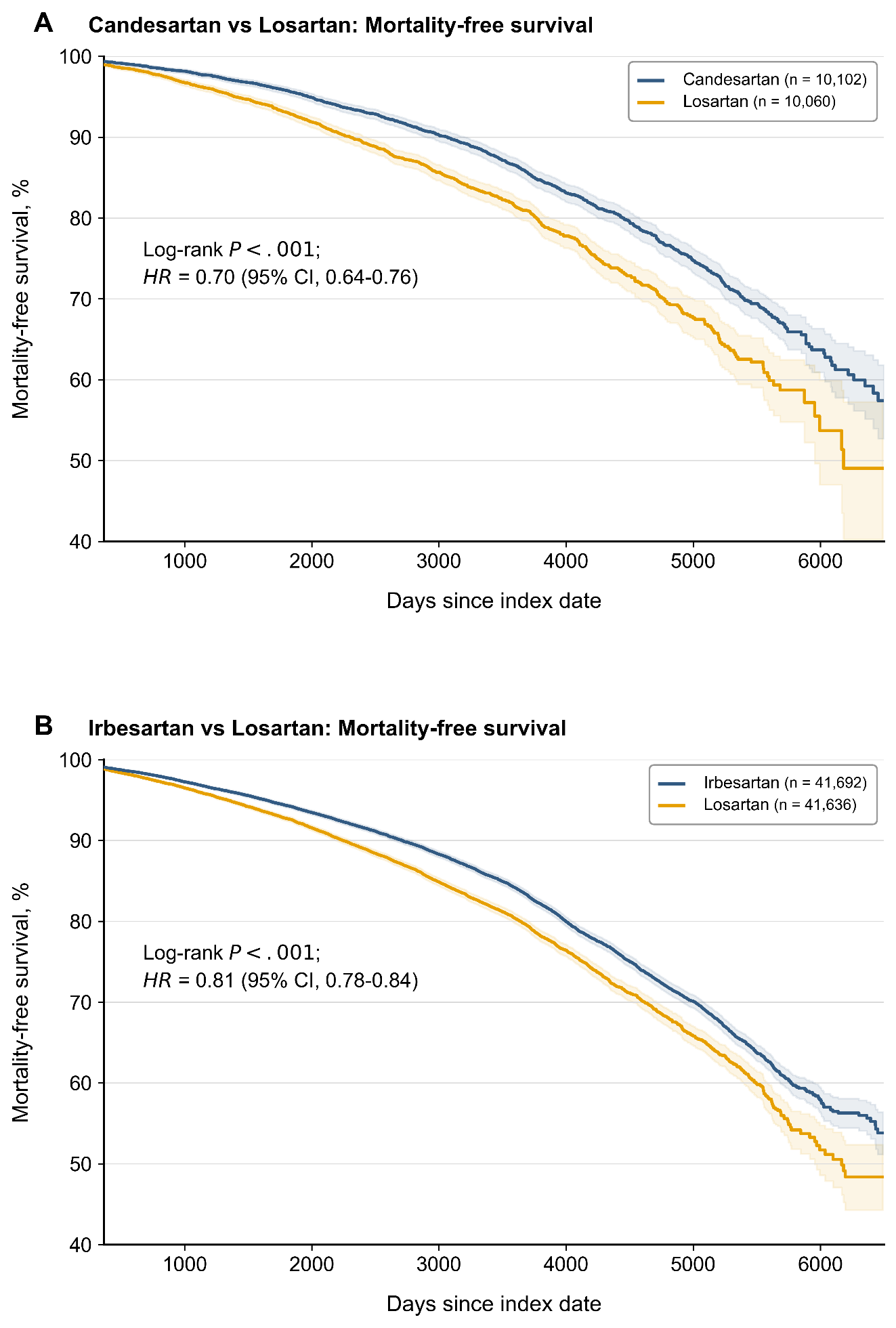

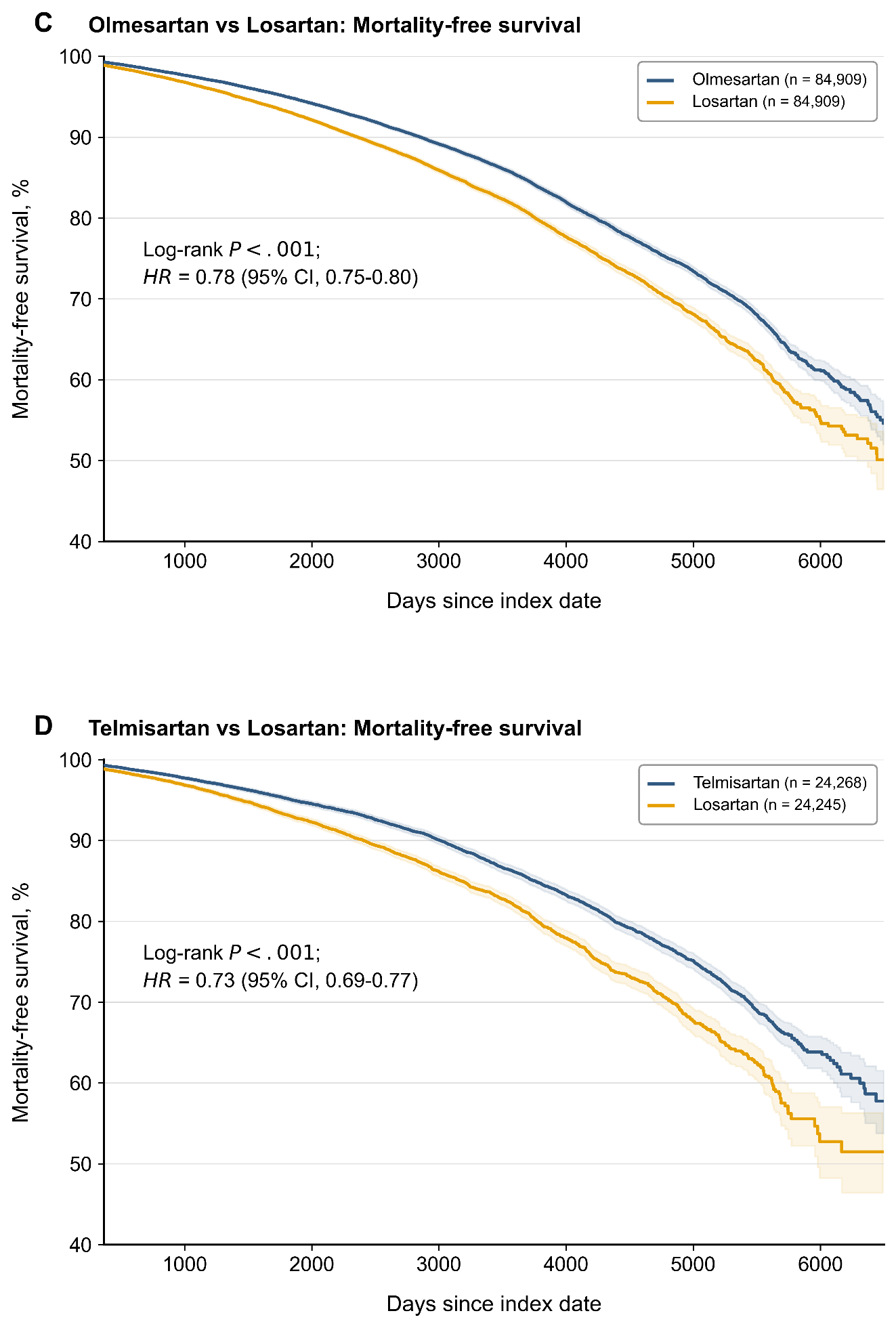

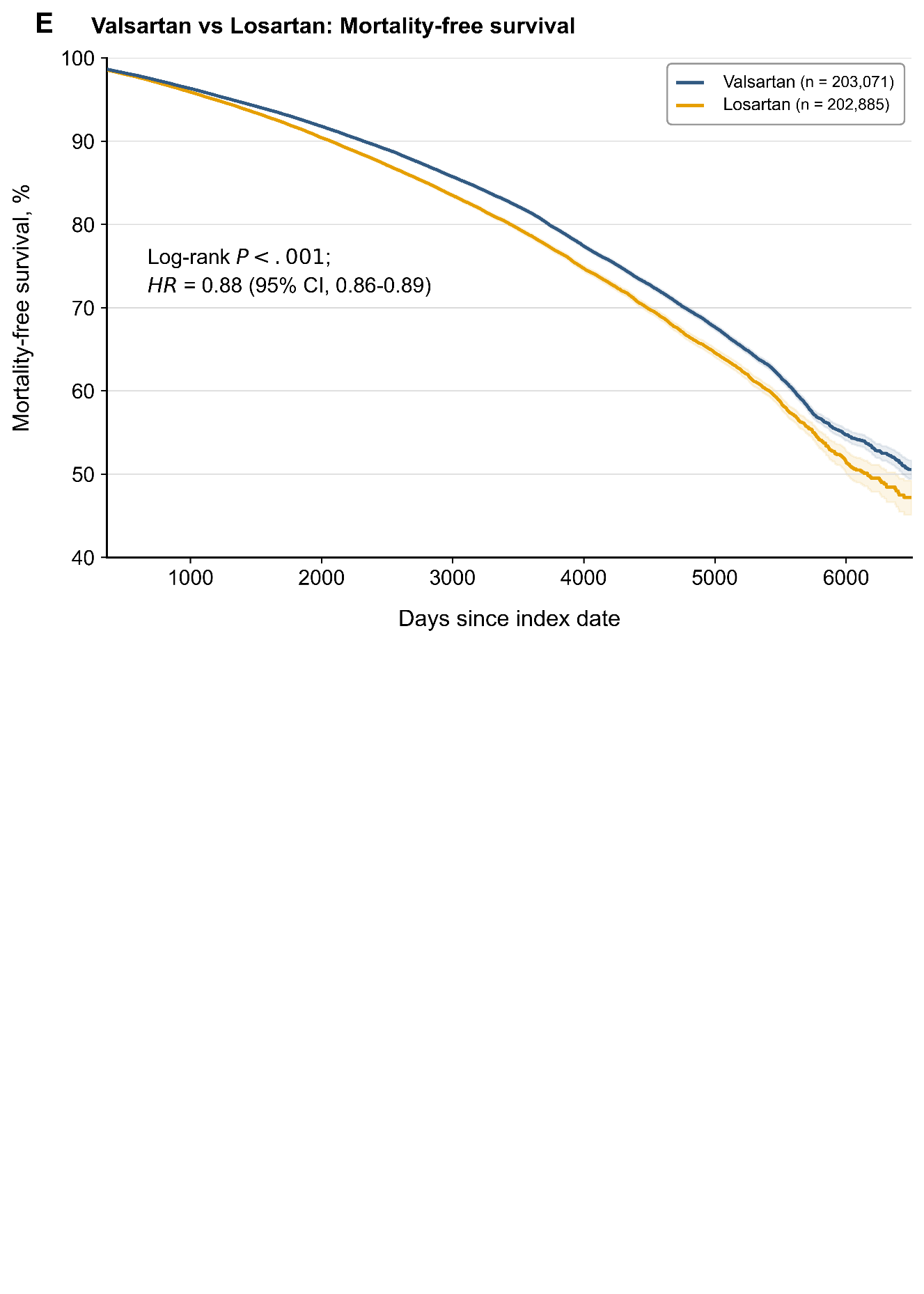

Kaplan–Meier curves show mortality-free survival for propensity-score–matched comparisons of other angiotensin receptor blockers (ARBs) with losartan (reference). Patients were followed from 1 day after the index date until death, loss to follow-up, or the end of the observation window. Hazard ratios (HRs) with 95% confidence intervals and log-rank P values were computed over the entire follow-up period. For visual clarity, curves are truncated during the sparse terminal follow-up period; this visual truncation did not affect the hazard-ratio estimates. Shaded areas indicate 95% confidence intervals, which widen at later time points as observations become sparse. Numbers at risk at individual follow-up time points are not shown because outcome-specific risk-set counts were not available from the aggregate TriNetX analytic output used for this study. Abbreviation: HR, hazard ratio. Python/Matplotlib code used to render this figure was generated using ChatGPT (OpenAI; GPT-5-series, 2026), Claude (Anthropic; Claude Opus 4.8 and earlier models, 2026), and/or Gemini (Google; 2026); the figure was verified against the source data.

**Table S13. Absolute risk reduction (ARR) and number needed to treat (NNT) at 10 years for all-cause dementia and death for within-class comparisons**

| **Outcome** | **Comparison** | **HR** | **ARR (%)** | **NNT** |
| --- | --- | --- | --- | --- |
| **All-Cause Dementia** | Candesartan vs Losartan | 0.72 | 2.17 | 46 |
|  | Irbesartan vs Losartan | 0.76 | 2.115 | 47 |
|  | Olmesartan vs Losartan | 0.83 | 1.253 | 80 |
|  | Telmisartan vs Losartan | 0.87 | 0.746 | 134 |
|  | Valsartan vs Losartan | 0.93 | 0.663 | 151 |
| **All-cause mortality** | Candesartan vs Losartan | 0.70 | 4.562 | 22 |
|  | Telmisartan vs Losartan | 0.74 | 4.324 | 23 |
|  | Irbesartan vs Losartan | 0.77 | 4.207 | 24 |
|  | Olmesartan vs Losartan | 0.75 | 4.066 | 25 |
|  | Valsartan vs Losartan | 0.85 | 2.951 | 34 |

Absolute risk reduction (ARR) and number needed to treat (NNT) were estimated at 10 years using Kaplan–Meier cumulative incidence estimates from the matched cohorts. ARR was calculated as the difference in absolute risk between treatment groups, and NNT as the reciprocal of ARR. ARR values are expressed as percentages, and NNT values were rounded to the nearest integer. Hazard ratios (HRs) correspond to analyses restricted to the same 10-year follow-up horizon, rounded to the 2nd decimal place, and therefore may differ from the primary full-follow-up estimates. Abbreviations: ARR, absolute risk reduction; NNT, number needed to treat; HR, hazard ratio.

**Table S14.** **Per-protocol-like (PP) vs. intention-to-treat (ITT) hazard ratio estimates for all-cause dementia for select comparisons**

| **Comparison** | **ARB (n)** | **ARB (events)** | **Lisinopril (n)** | **Lisinopril (events)** | **HR** | **95% CI** |
| --- | --- | --- | --- | --- | --- | --- |
| Irbesartan vs Lisinopril ITT (main analysis) | 41,685 | 2,181 | 41,659 | 2,957 | 0.69 | (0.66, 0.73) |
| Irbesartan vs Lisinopril PP | 17,863 | 558 | 17,852 | 1,086 | 0.59 | (0.53, 0.65) |
| Losartan vs Lisinopril ITT (main analysis) | 564,485 | 29,040 | 564,436 | 40,473 | 0.93 | (0.91, 0.94) |
| Losartan vs Lisinopril PP | 341,416 | 14,419 | 341,239 | 21,731 | 0.86 | (0.85, 0.88) |

Hazard ratios for all-cause dementia are shown for intention-to-treat (ITT) and per-protocol-like (PP) analyses in propensity-score–matched cohorts. The ITT analysis classified participants according to the index prescription and followed them regardless of subsequent treatment changes. The PP analysis required sustained exposure to the index medication, defined as at least two prescriptions within 12 months and no prior or subsequent exposure to alternative angiotensin receptor blockers or angiotensin-converting–enzyme inhibitors (ACEIs). ACEIs included lisinopril, benazepril, quinapril, perindopril, enalapril, ramipril, trandolapril, captopril, fosinopril, and moexipril. Follow-up began one day after the index event and continued until outcome occurrence, death, or last recorded clinical encounter. Hazard ratios were estimated using Cox proportional-hazards models, with 95% confidence intervals shown. These analyses were performed to assess the robustness of findings to treatment adherence and switching. Abbreviations: ARB, angiotensin receptor blocker; ITT, intention-to-treat; HR, hazard ratio; CI, confidence interval.

**Table S15.** **1- and 3-year lag analyses hazard ratio estimates for all-cause dementia in select comparisons**

| **Comparison** | **ARB (n)** | **ARB (events)** | **Lisinopril (n)** | **Lisinopril (events)** | **HR** | **95% CI** |
| --- | --- | --- | --- | --- | --- | --- |
| Irbesartan vs Lisinopril ITT (main analysis) | 41,685 | 2,181 | 41,659 | 2,957 | 0.69 | (0.66, 0.73) |
| Irbesartan vs Lisinopril ITT (1-year lag) | 41,421 | 1,922 | 41,274 | 2,458 | 0.72 | (0.68, 0.76) |
| Irbesartan vs Lisinopril ITT (3-year lag) | 41,031 | 1,532 | 40,702 | 1,886 | 0.73 | (0.68, 0.78) |
| Losartan vs Lisinopril ITT (main analysis) | 564,485 | 29,040 | 564,436 | 40,473 | 0.93 | (0.91, 0.94) |
| Losartan vs Lisinopril ITT (1-year lag) | 559,357 | 23,901 | 558,468 | 34,495 | 0.94 | (0.92, 0.95) |
| Losartan vs Lisinopril ITT (3-year lag) | 552,857 | 17,401 | 550,487 | 26,514 | 0.95 | (0.93, 0.96) |

Hazard ratios for all-cause dementia are shown for intention-to-treat (ITT) analyses incorporating 1-day (main analysis), 1-year, and 3-year lag periods in propensity-score–matched cohorts. In all analyses, participants were classified according to the index prescription and followed regardless of subsequent treatment changes. The primary ITT analysis applied a 1-day exclusion period after the index event, with follow-up beginning thereafter. Lag analyses extended this exclusion period to 1 or 3 years to assess for and reduce potential bias from reverse causation, with events occurring during the lag period excluded. Follow-up continued until outcome occurrence, death, or last recorded clinical encounter. Hazard ratios were estimated using Cox proportional-hazards models, with 95% confidence intervals shown. Abbreviations: ARB, angiotensin receptor blocker; ITT, intention-to-treat; HR, hazard ratio; CI, confidence interval.

**Table S16.** **10-year truncated follow up for all-cause dementia in select comparisons**

| **Comparison** | **ARB (n)** | **ARB (events)** | **Lisinopril (n)** | **Lisinopril (events)** | **HR** | **95% CI** |
| --- | --- | --- | --- | --- | --- | --- |
| Irbesartan vs Lisinopril ITT (main analysis) | 41,685 | 2,181 | 41,659 | 2,957 | 0.69 | (0.66, 0.73) |
| Irbesartan vs Lisinopril 10 year truncated | 41,685 | 1,672 | 41,665 | 2,397 | 0.69 | (0.65, 0.73) |
| Losartan vs Lisinopril ITT (main analysis) | 564,485 | 29,040 | 564,436 | 40,473 | 0.93 | (0.91, 0.94) |
| Losartan vs Lisinopril 10 year truncated | 564,485 | 25,850 | 564,436 | 34,213 | 0.92 | (0.90, 0.93) |

Hazard ratios for all-cause dementia are shown for intention-to-treat (ITT) analyses using the full available follow-up (main analysis) and follow-up truncated at 10 years in propensity-score–matched cohorts. In all analyses, participants were classified according to the index prescription and followed regardless of subsequent treatment changes. The primary ITT analysis applied a 1-day exclusion period after the index event, with follow-up beginning thereafter. In the 10-year truncated analyses, follow-up was additionally censored at 10 years to assess the potential influence of differences in follow-up duration. Follow-up continued until outcome occurrence, death, or last recorded clinical encounter, whichever occurred first. Hazard ratios were estimated using Cox proportional-hazards models, with 95% confidence intervals shown. Abbreviations: ARB, angiotensin receptor blocker; ITT, intention-to-treat; HR, hazard ratio; CI, confidence interval.

**Table S17.** **Low vs high dose hazard ratio estimates for all-cause dementia in select comparisons**

| **Comparison** | **ARB (n)** | **ARB (events)** | **Lisinopril (n)** | **Lisinopril (events)** | **HR** | **95% CI** |
| --- | --- | --- | --- | --- | --- | --- |
| Irbesartan vs Lisinopril (main analysis) | 41,685 | 2,181 | 41,659 | 2,957 | 0.69 | (0.66, 0.73) |
| Low Dose Irbesartan (75, 150 mg) vs Low Dose Lisinopril (2.5, 5, 10 mg) | 24,543 | 1,035 | 24,532 | 1,696 | 0.68 | (0.63, 0.74) |
| High Dose Irbesartan (300 mg) vs High Dose Lisinopril (20, 30, 40 mg) | 21,318 | 1,041 | 21,327 | 1,592 | 0.73 | (0.68, 0.79) |
| Losartan vs Lisinopril (main analysis) | 564,485 | 29,040 | 564,436 | 40,473 | 0.93 | (0.91, 0.94) |
| Low Dose Losartan (25, 50 mg) vs Low Dose Lisinopril (2.5, 5, 10 mg) | 414,188 | 21,238 | 413,731 | 29,559 | 0.93 | (0.91, 0.95) |
| High Dose Losartan (100 mg) vs High Dose (20, 30, 40 mg) Lisinopril | 303,069 | 17,138 | 303,590 | 23,429 | 0.89 | (0.87, 0.91) |

Hazard ratios for all-cause dementia are shown for intention-to-treat (ITT) analyses comparing low- and high-dose categories of angiotensin receptor blockers (ARBs) compared with lisinopril in propensity-score–matched cohorts. Dose categories were defined a priori based on commonly prescribed doses and were assigned at the index event using RxNorm-coded medication records, which include standardized drug and strength information. Participants were classified according to the index prescription and followed regardless of subsequent treatment changes, with dose categories not updated during follow-up. Follow-up began one day after the index event and continued until outcome occurrence, death, or last recorded clinical encounter. Hazard ratios were estimated using Cox proportional-hazards models, with 95% confidence intervals shown. Abbreviations: ARB, angiotensin receptor blocker; ITT, intention-to-treat; HR, hazard ratio; CI, confidence interval.

**Figure S10**. **Negative control outcomes for between-class comparisons**

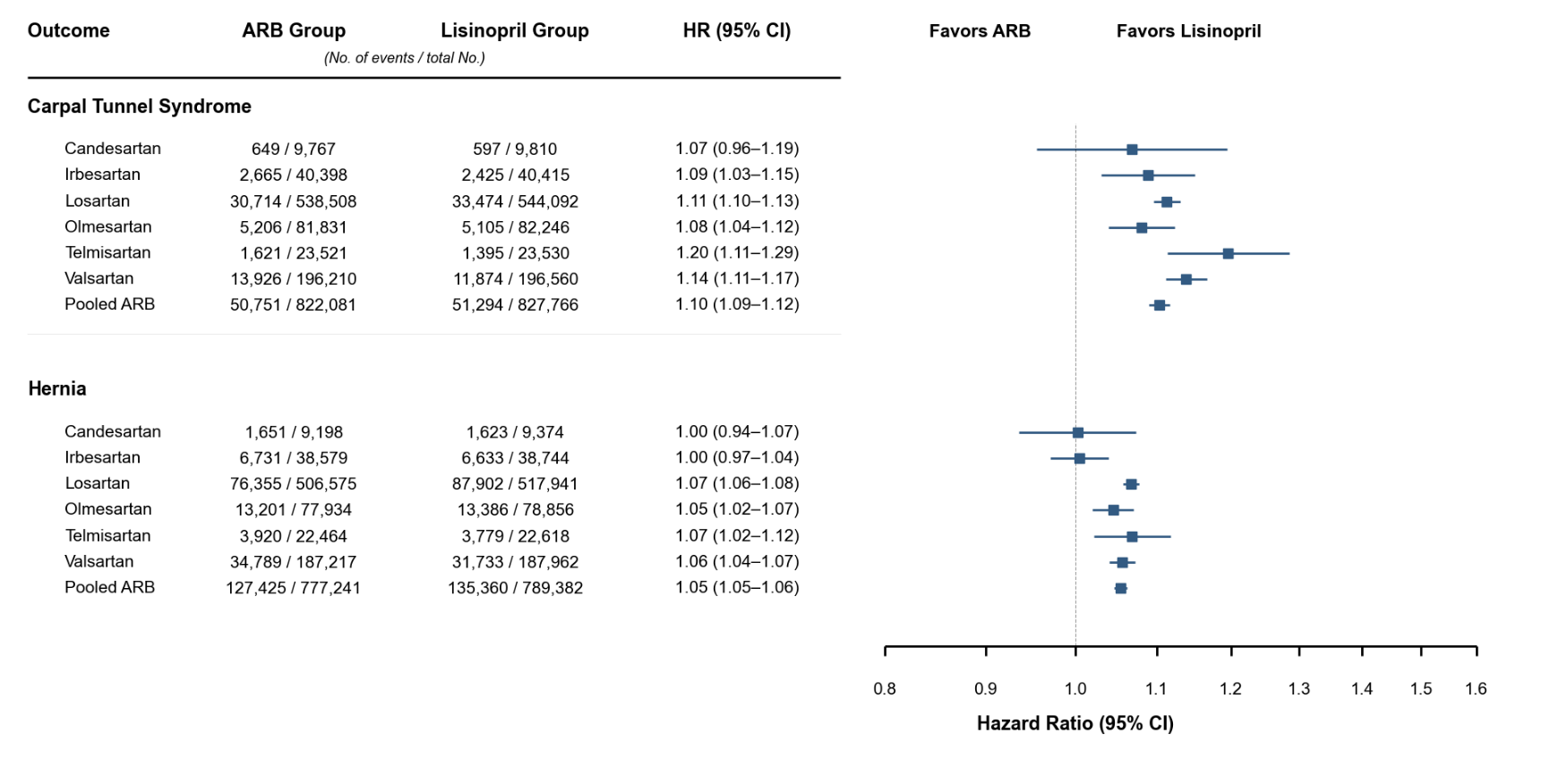

Forest plots show hazard ratios and 95% confidence intervals for negative control outcomes comparing individual and pooled ARBs with lisinopril (reference). Hazard ratios are plotted on a logarithmic scale. Outcomes were defined using International Classification of Diseases, Tenth Revision (ICD-10) codes. Python/Matplotlib code used to render this figure was generated using ChatGPT (OpenAI; GPT-5-series, 2026), Claude (Anthropic; Claude Opus 4.8 and earlier models, 2026), and/or Gemini (Google; 2026); the figure was verified against the source data.

**Figure S11**. **Negative control outcomes for within class comparisons**

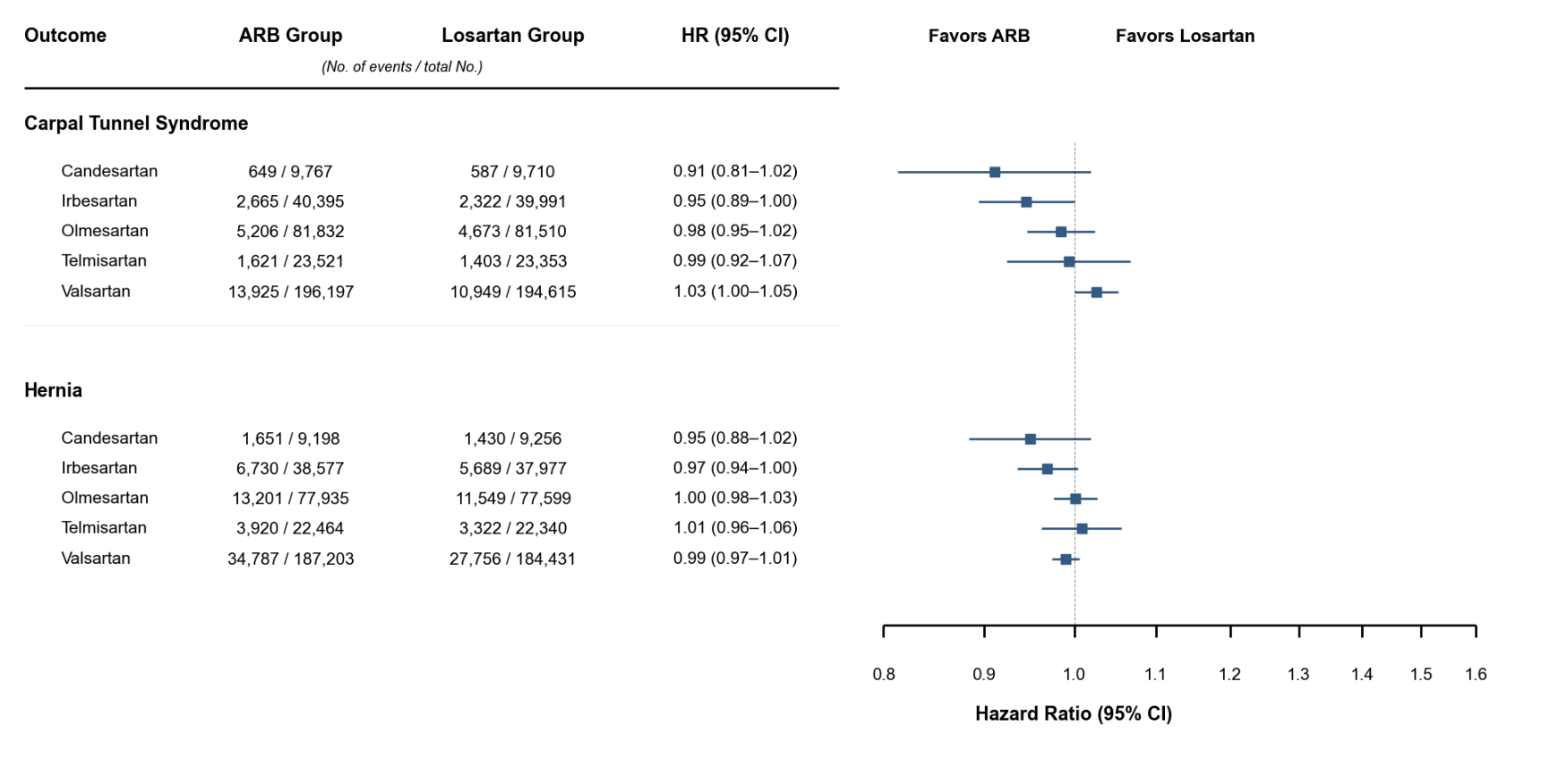

Forest plots show hazard ratios and 95% confidence intervals for negative control outcomes comparing other ARBs with losartan (reference). Hazard ratios are plotted on a logarithmic scale. Outcomes were defined using International Classification of Diseases, Tenth Revision (ICD-10) codes. Python/Matplotlib code used to render this figure was generated using ChatGPT (OpenAI; GPT-5-series, 2026), Claude (Anthropic; Claude Opus 4.8 and earlier models, 2026), and/or Gemini (Google; 2026); the figure was verified against the source data.

**Table S18**. **E-values for all-cause dementia hazard estimates for between-class and within-class comparisons**

| **Between-class** | **Comparison** | **HR (95% CI)** | **E-value (point)** | **E-value (CI limit)** |
| --- | --- | --- | --- | --- |
|  | Candesartan vs Lisinopril | 0.66 (0.58-0.74) | 2.42 | 2.04 |
|  | Irbesartan vs Lisinopril | 0.69 (0.66-0.73) | 2.24 | 2.07 |
|  | Olmesartan vs Lisinopril | 0.79 (0.76-0.82) | 1.86 | 1.74 |
|  | Telmisartan vs Lisinopril | 0.84 (0.78-0.90) | 1.68 | 1.48 |
|  | Valsartan vs Lisinopril | 0.86 (0.84-0.88) | 1.60 | 1.53 |
|  | Losartan vs Lisinopril | 0.93 (0.91-0.94) | 1.37 | 1.32 |
|  | Pooled ARB vs Lisinopril | 0.86 (0.85-0.87) | 1.59 | 1.55 |
| **Within-class** | **Comparison** | **HR (95%CI)** | **E-value (point)** | **E-value (CI limit)** |
|  | Candesartan vs Losartan | 0.68 (0.60-0.77) | 2.31 | 1.92 |
|  | Irbesartan vs Losartan | 0.77 (0.72-0.81) | 1.94 | 1.76 |
|  | Olmesartan vs Losartan | 0.84 (0.80-0.88) | 1.67 | 1.54 |
|  | Telmisartan vs Losartan | 0.86 (0.79-0.93) | 1.61 | 1.38 |
|  | Valsartan vs Losartan | 0.94 (0.91-0.96) | 1.33 | 1.24 |

To assess the potential impact of unmeasured confounding, E-values were calculated using established methods for the primary outcome. The E-value represents the minimum strength of association that an unmeasured confounder would need to have with both the exposure and the outcome, conditional on measured covariates, to explain away the observed association. E-values were computed for both the point estimate and the confidence limit closest to the null. Values were calculated using unrounded HRs and confidence limits; displayed HRs are rounded. Larger E-values indicate that stronger unmeasured confounding would be required to explain away the observed association. Abbreviations: HR, hazard ratio; CI, confidence interval.

**Figure S12. Positive control outcome (angioedema) for between class comparisons**

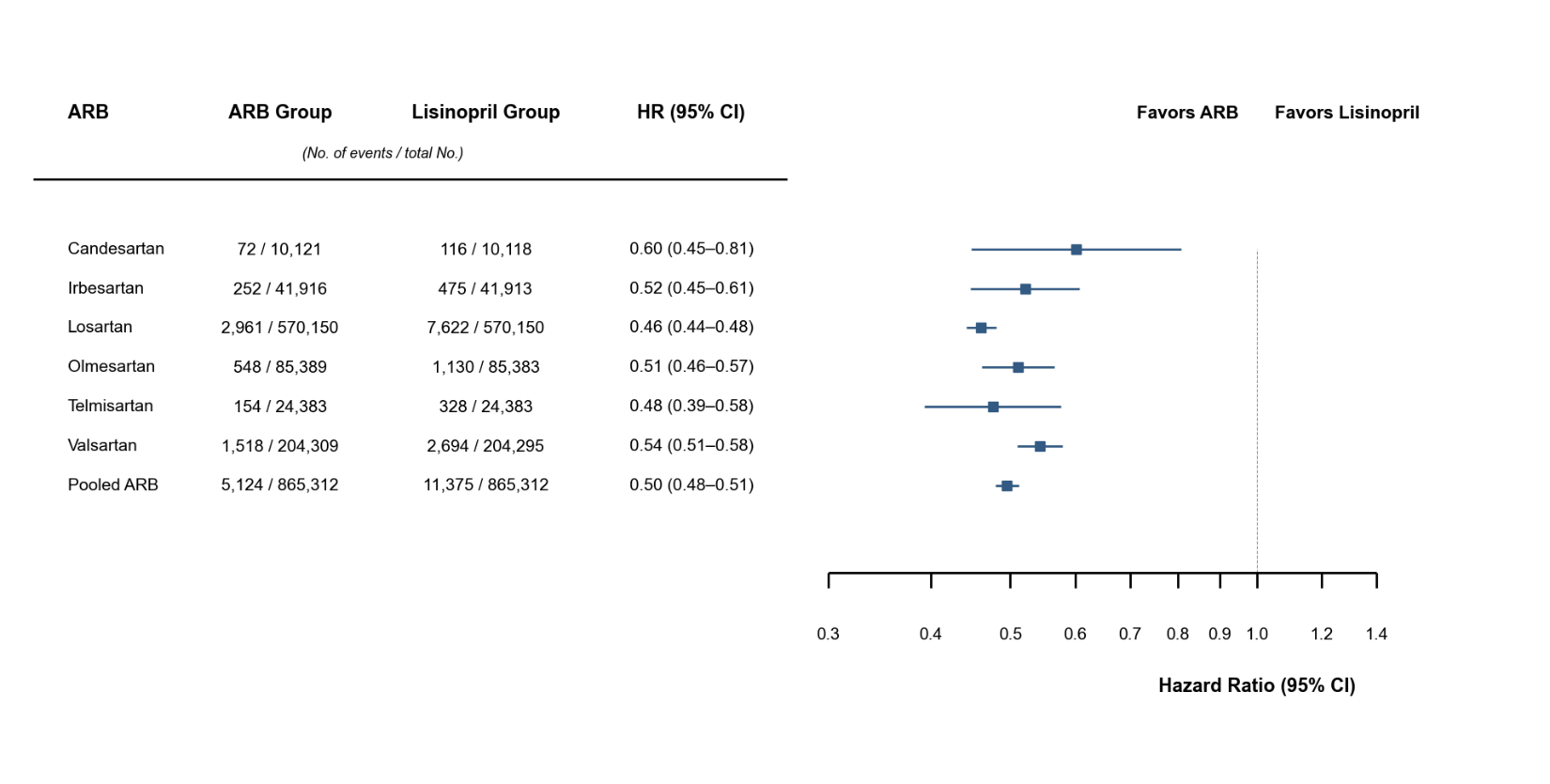

Forest plots show hazard ratios and 95% confidence intervals for positive control outcome (Angioedema) comparing ARBs with lisinopril (reference). Hazard ratios are plotted on a logarithmic scale. Angioedema was pre-specified as a positive control because ARBs are established to carry lower angioedema risk than ACE inhibitors. Outcomes were defined using International Classification of Diseases, Tenth Revision (ICD-10) codes. Python/Matplotlib code used to render this figure was generated using ChatGPT (OpenAI; GPT-5-series, 2026), Claude (Anthropic; Claude Opus 4.8 and earlier models, 2026), and/or Gemini (Google; 2026); the figure was verified against the source data.

**Appendix S1. TriNetX query and index event specifications**

Cohort, index-event, and time-window criteria for the irbesartan-versus-lisinopril intention-to-treat analysis of incident all-cause dementia, generated from the TriNetX platform output. All other between-class comparisons used the identical cohort structure with the index medication changed to the relevant agent; within-class comparisons used losartan as the reference. Outcome codes are listed in eTable 3, and sensitivity analyses (per-protocol, lagged, and dose-stratified) modified only the persistence, index-shift, or dose parameters described in the Methods.

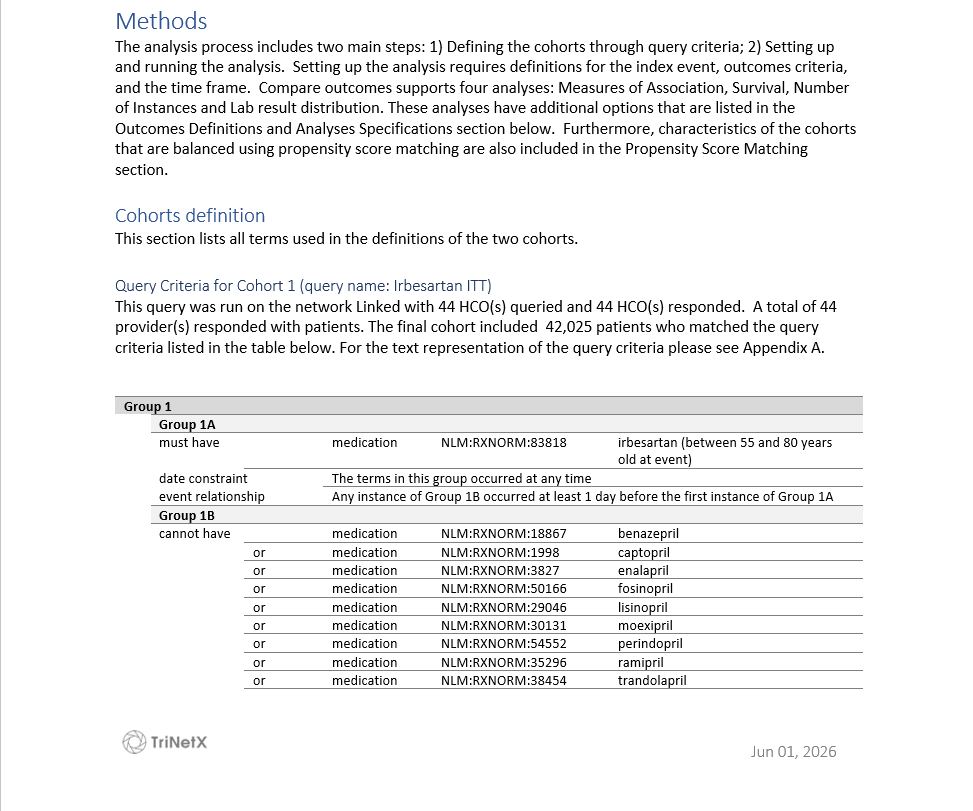

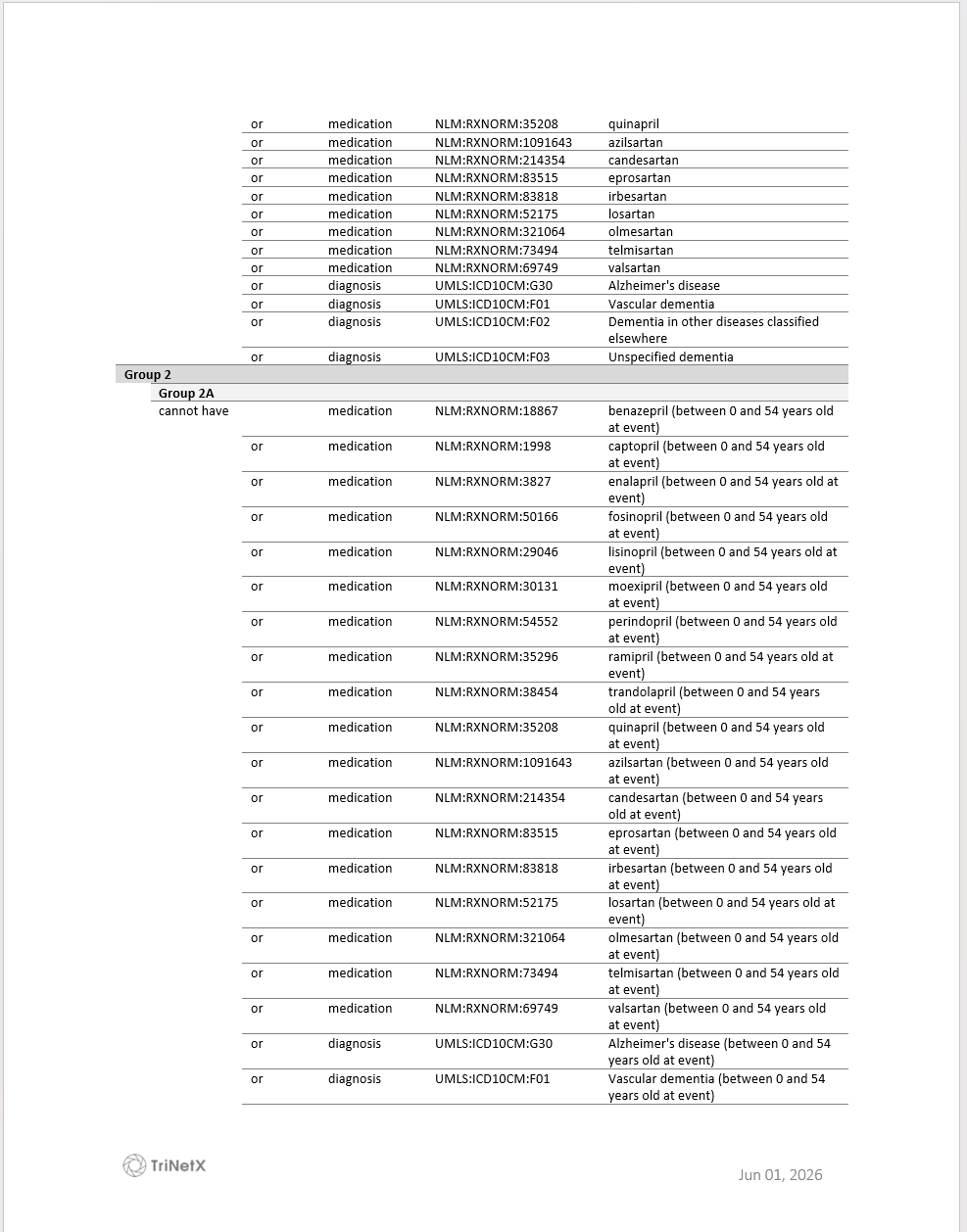

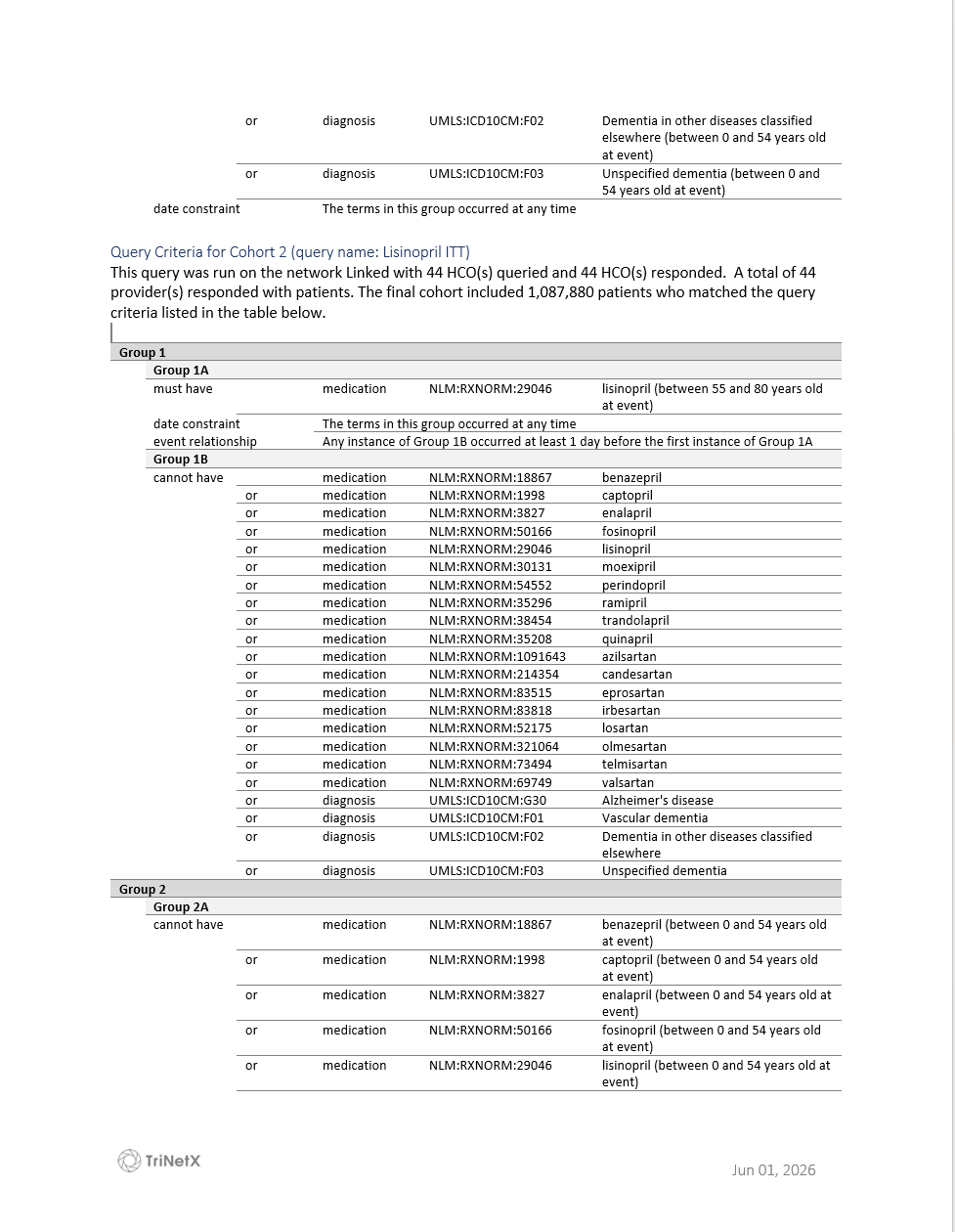

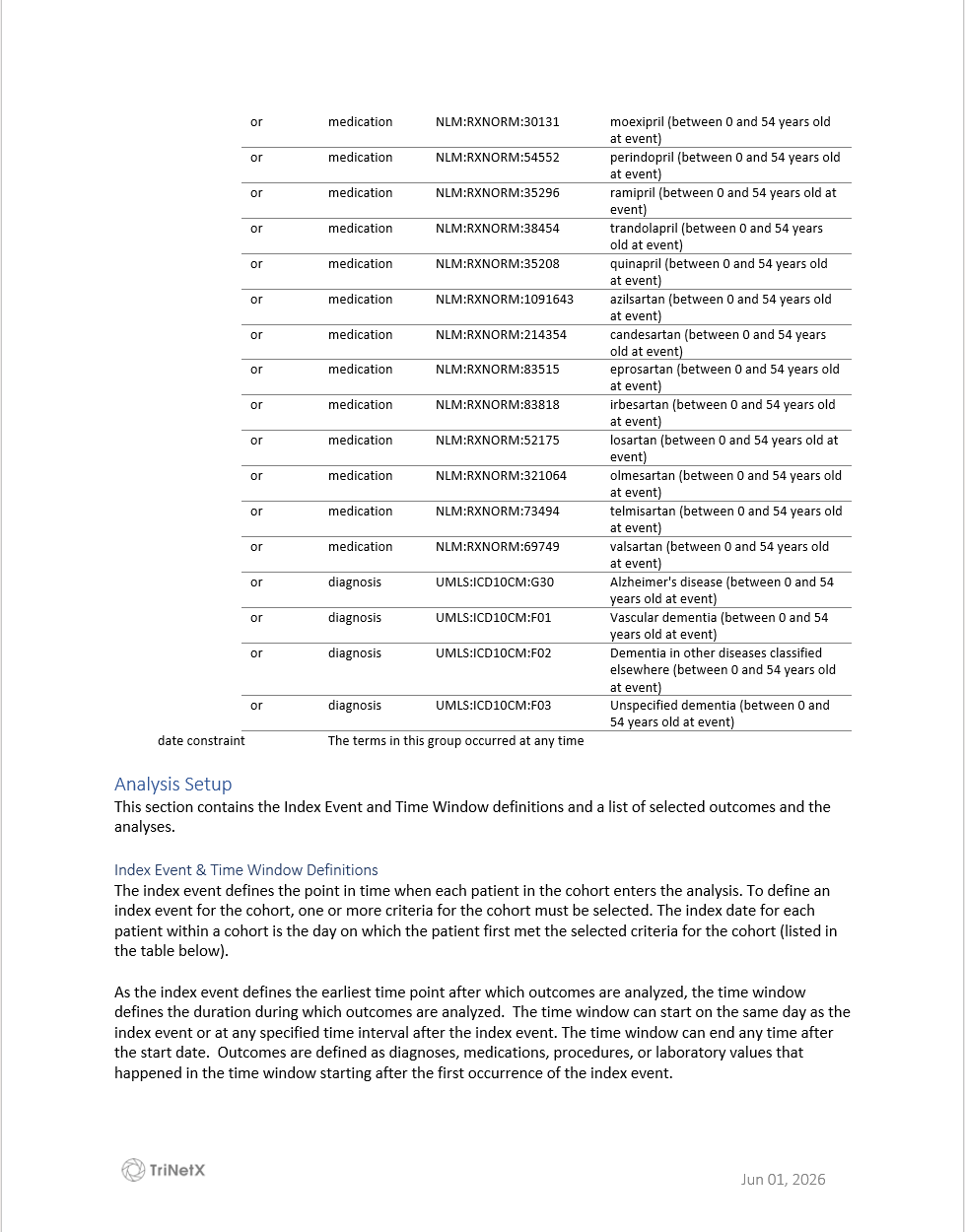

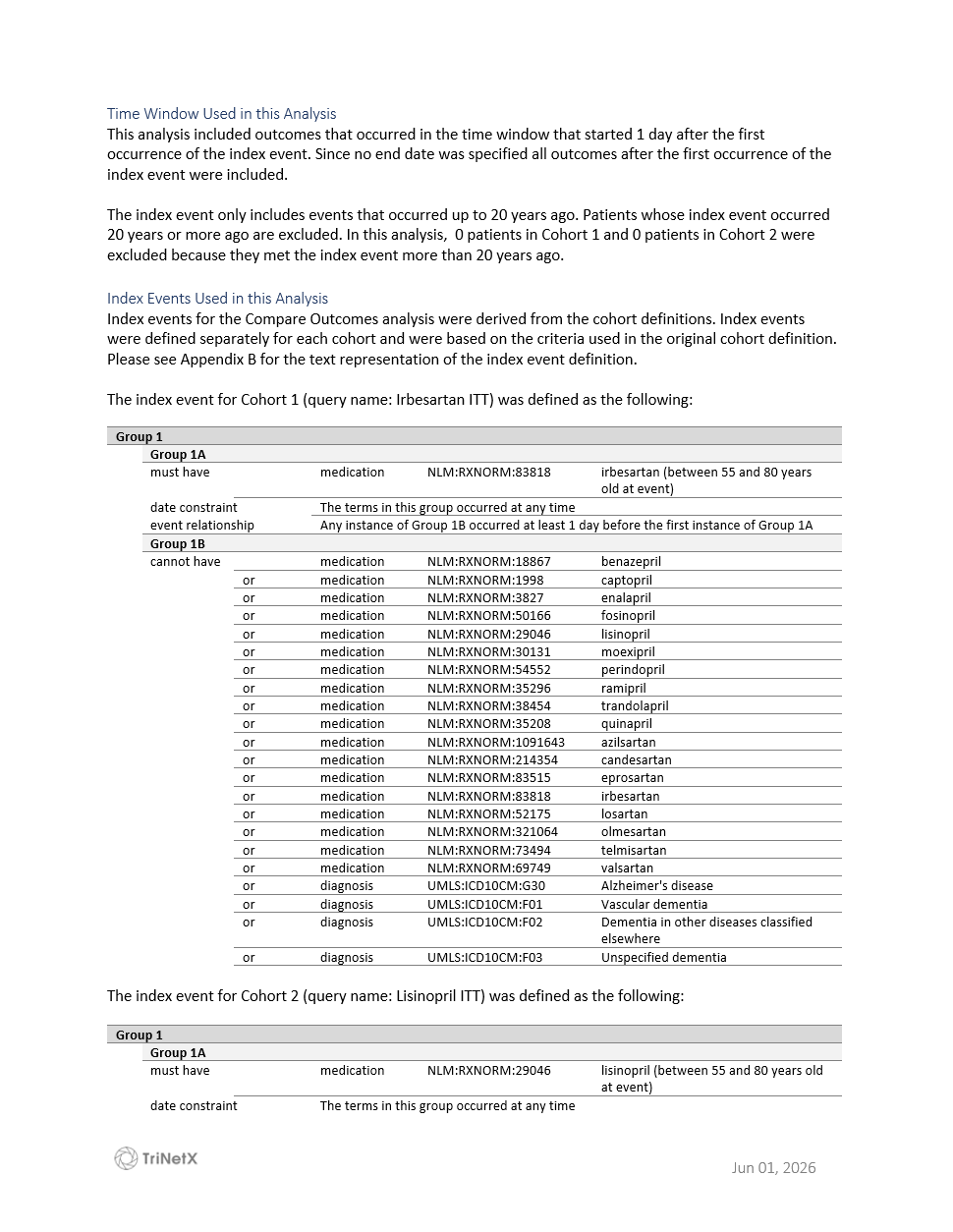

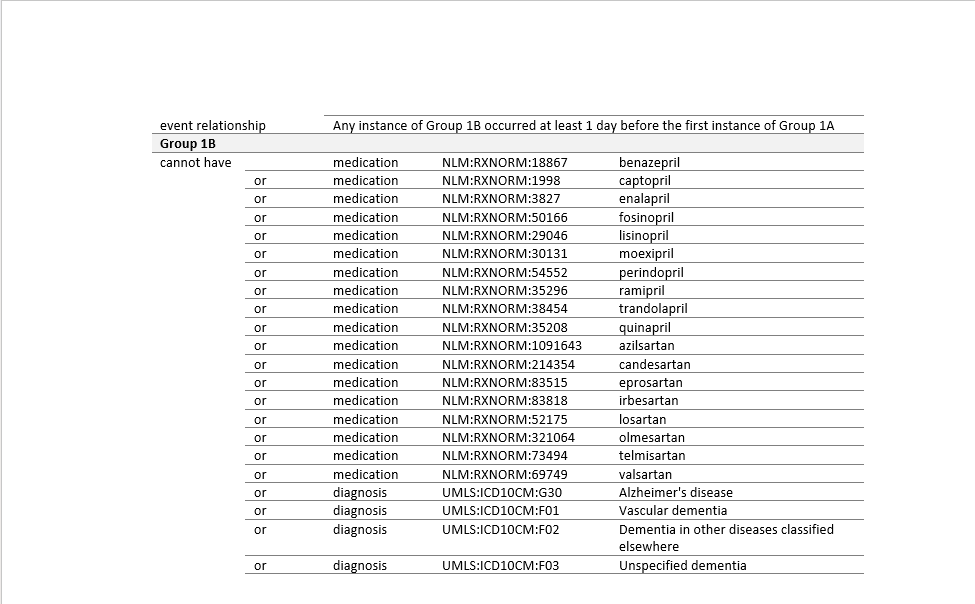

**Appendix S2. Population-level projections of potential dementia reduction**

To illustrate the potential population-level magnitude of the observed absolute risk differences, we scaled the 10-year absolute risk reductions to a hypothetical population of 1 million treated individuals. We estimated the number of potentially fewer dementia cases if patients receiving lisinopril were instead treated with ARBs associated with lower dementia risk, or if patients receiving losartan were instead treated with other ARBs associated with lower dementia risk.

Across between-class comparisons, absolute risk reductions for all-cause dementia ranged from 0.72% to 3.07%, corresponding to NNT values from 33 to 138 over 10 years. The largest absolute risk reduction was observed for candesartan (ARR, 3.07%; NNT, 33), whereas losartan showed the smallest absolute risk reduction (ARR, 0.72%; NNT, 138). Within-class comparisons relative to losartan also demonstrated variation in absolute differences, with ARRs ranging from 0.66% to 2.17% (NNTs, 46–151).

When expressed on a population scale, these differences correspond to approximately **7,230 to 30,670 potentially fewer cases of dementia per 1 million treated individuals over 10 years** if the observed absolute risk differences for ARBs versus lisinopril were applied to a population of this size.

**Relative to losartan**, the corresponding estimates were approximately **6,630 to 21,700 potentially fewer cases of dementia per 1 million treated individuals** **over 10 years** if the observed absolute risk differences for other ARBs versus losartan were applied to a population of this size.

These projections are based on associations observed in a single observational study and should be interpreted as illustrative rather than causal estimates. Confirmation in independent datasets and, ideally, randomized clinical trials is needed.
